# Factors associated with overweight or obesity in children under five years old: a rapid review

**DOI:** 10.1101/2024.10.14.24314839

**Authors:** Jessica Williams, Charlotte Bowles, Antonia Needham-Taylor, David Jarrom, Elizabeth Gillen, Juliet Hounsome, Jacob Davies, Rhiannon Tudor Edwards, Adrian Edwards, Alison Cooper, Ruth Lewis

## Abstract

Over one-quarter of children in Wales aged four-to-five live with overweight or obesity. Children with overweight or obesity may experience health issues during childhood or adolescence, or both. They are also more likely to have overweight or obesity through to adulthood. A wide range of biological (e.g. genetics), psychological, environmental (e.g. barriers to physical activity and access to healthier foods) and societal (e.g. lifestyle and peer influence) factors can potentially be associated with child overweight or obesity.

Evidence reviews of factors associated with child overweight or obesity already exist, but these examine the influence of the above factors across a range of ages. No up-to-date review focuses specifically on factors associated with overweight or obesity when both the factor and its impact on weight status were studied in children under five years of age. We aimed to review existing reviews to identify the factors associated with overweight and obesity, specifically when impact on weight status was reported in children under five years of age.

The review included evidence available up until December 2023, and 30 systematic reviews were identified. A wide range of biological, psychological, environmental and societal factors were consistently found to be associated with an increased risk of child overweight and/or obesity.

Factors consistently found to be associated with a decreased risk of child overweight or obesity were breastfeeding and larger household size. Most of the high-certainty evidence related to child overweight. No high-certainty evidence was identified on the association between factors and child obesity specifically, and further research studies are needed.

The high-certainty evidence supports helping women with overweight (who are thinking about having a baby or trying to conceive) to lose weight, reducing rapid weight gain during the first 12 months of life, providing opportunities for children of working mothers to access healthier foods and be more physically active. The moderate-certainty evidence supports promoting breastfeeding, reducing rapid weight gain during the first 13 months of life, monitoring the child’s growth rate during the first two years of life (particularly for babies with catch-up growth), promoting baby-led weaning, reducing consumption of sugary drinks, and educating and supporting the wider caregivers to provide healthier foods and opportunities for play and physical activity.

The findings from this review may differ from other reviews conducted to inform practice. This review only reports on overweight or obesity outcomes where they are measured before the age of five years, whereas previous reviews have measured these outcomes over a wider age range. This review also focusses on evidence that specifically classifies children as living with overweight or obesity using body mass index (or other well-accepted measures for children under two years).

**Funding statement:** The authors and their Institutions were funded for this work by the Health and Care Research Wales Evidence Centre, itself funded by Health and Care Research Wales on behalf of Welsh Government

## 1. BACKGROUND

### 1.1 Who is this review for?

This work was requested by the Welsh Government Public Health and Inequalities team, as part of the Wellbeing and Future Generations Act work, to help inform the ‘Healthy Weight: Healthy Wales’ strategy and delivery plans (Welsh Government 2023). This rapid review may be helpful for policy makers and others involved in children’s health in Wales.

### 1.2 Background and purpose of this review

Obesity and overweight are terms that refer to an excess of body fat and the resulting weight gain (Public Health Wales 2018). Overweight and obesity are multifactorial and complex, and their causes vary between population groups and across a person’s life course (Public Health Wales 2018).

The most widely used measure of obesity is the body mass index (BMI) (Public Health Wales 2018). Less commonly used are measures of central obesity, such as the waist-to-hip ratio, waist circumference and abdominal fat. BMI in children is classified using thresholds that vary to take into account the child’s age and sex. There are different definitions of obesity and overweight in children, but definitions include those such as the British 1990 growth reference scale (UK90), which defines obesity as a BMI at or above the 95^th^ percentile for children and adolescents of the same age and sex, and overweight as BMIs between the 85 and 94 percentile on age-growth charts (Public Health Wales 2018). BMI is not recommended for use in children under two years (Centers for Disease Control and Prevention 2015). Instead, the World Health Organization developed growth charts based on weight-for-length percentiles, head circumference, length-for-age percentiles and weight-for-age percentiles for infants aged less than two years (Centers for Disease Control and Prevention 2015).

Over one-quarter of children in Wales aged four to five years old are either overweight or obese (Public Health Wales). The proportion of children aged four to five years classified as overweight and obese is higher than in England (Public Health Wales 2018). Early childhood is a critical period for the development of obesity or overweight, because, for example, dietary and physical-activity habits formed during these years can last a lifetime. Children who are overweight or obese are more likely to be overweight or obese through to adulthood, which can cause associated health problems such as diabetes, heart disease, cancer, and premature mortality (NHS Wales). Children who are obese may also experience health issues during childhood or adolescence, such as non-alcoholic fatty liver disease, gall stones, asthma, sleep-disordered breathing and musculoskeletal conditions. There are also emotional and psychological effects of children being obese or overweight, including bullying, low self-esteem, anxiety and depression. They may find physical activity difficult, which can increase the likelihood of a sedentary lifestyle (NHS Wales).

One of the aims of the Healthy Weight Strategy is to reduce and prevent obesity, with a particular focus on childhood obesity in pre-school age children (Welsh Government 2023). The aim of this work is to identify the factors associated with childhood obesity and overweight, specifically where weight status is affected before the age of five years. This will help to inform the start of an ongoing piece of work looking into the root causes of obesity or overweight in children under five years old.

**Review Question:** Which factors are associated with childhood obesity or overweight in children under five years old?

The review question was addressed using a review of existing systematic reviews. The eligibility criteria and methods for conducting the review are detailed in Section 5.

## 2. RESULTS

### 2.1 Overview of the Evidence Base

#### 2.1.1 Quantitative evidence

Twenty-eight systematic reviews reported on the association between factors and obesity and/or overweight in children under five years. Most of these reviews included studies from Organization for Economic Cooperation and Development countries, and/or focused on predominantly Caucasian race or ethnicity, and were therefore judge to be relevant to the UK setting and population. Twelve reviews included studies conducted in the UK. The USA was the most frequently reported country in which studies were conducted in the reviews (19/28 reviews). Only one review was identified as likely to be not relevant to the UK. Five-out-of-28 reviews included children less than two years of age.

Figure 1 is an interactive evidence map of the study design and characteristics of the quantitative evidence. Figure 2 is an interactive map of the outcomes from the quantitative evidence. The results (and the classification system developed to structure these) are presented in Section 2.2.

**Figure 1.** (accessed here: https://healthtechnology.wales/wp-content/uploads/20240418-Figure-1-design-and-characteristics-map.html). An interactive visual display of systematic reviews investigating factors potentially associated with childhood obesity and/or overweight. In addition to the factors reported, the map details the outcomes (obesity, overweight, obesity/overweight) and how this was measured. Reviews are divided by their likely relevance to UK/Wales (Green: likely relevant; Blue: likely not to be relevant; Purple: relevance unclear). Generated using v.2.3.0 of the EPPI-Mapper, powered by EPPI Reviewer.

**Figure 2.** (accessed here: https://healthtechnology.wales/wp-content/uploads/20240418-Figure-2-outcomes-map.html). An interactive visual display of the outcomes of the studies included in the systematic reviews investigating factors potentially associated with childhood obesity and/or overweight. The map describes whether the factors were associated with a statistically significant increased risk, statistically significant decreased risk, or had no statistically significant relationship with childhood obesity and/or overweight. Additionally, the map details the age categories at which the outcomes were measured, and the outcomes (obesity, overweight, obesity or overweight). Reviews are divided by the certainty of the evidence; Green: high certainty; Orange: moderate certainty; Blue: low certainty; Purple: very low certainty. Generated using v.2.3.0 of the EPPI-Mapper, powered by EPPI Reviewer.

The certainty of the evidence for each factor (or outcome) was categorised using the Grading of Recommendations, Assessment, Development, and Evaluations (GRADE) approach into high, moderate, low or very low certainty. These ratings provide an indication of how much confidence to place in the findings. For the purpose of this review, we assessed each individual study for how certain the findings are, rather than assess the overall body of evidence for each factor, which is how GRADE is generally applied. The definitions of the GRADE ratings are provided in Table 1. Further detail on how the evidence was assessed are provided in the methods Section 5.10.

**Table 1:** GRADE ratings of the certainty in the evidence and their definitions (Higgins JPT 2023)

| GRADE rating | Definition |
| --- | --- |
| **High** | We are very confident that the true effect lies close to that of the estimate of the effect |
| **Moderate** | We are moderately confident in the effect estimate: The true effect is likely to be close to the estimate of the effect, but there is a possibility that it is substantially different |
| **Low** | Our confidence in the effect estimate is limited: The true effect may be substantially different from the estimate of the effect. |
| **Very low** | We have very little confidence in the effect estimate: The true effect is likely to be substantially different from the estimate of effect |

A list of the categories of factors, and specific types of factors within each category, which were covered by this review are presented in Table 2.

**Table 2.**
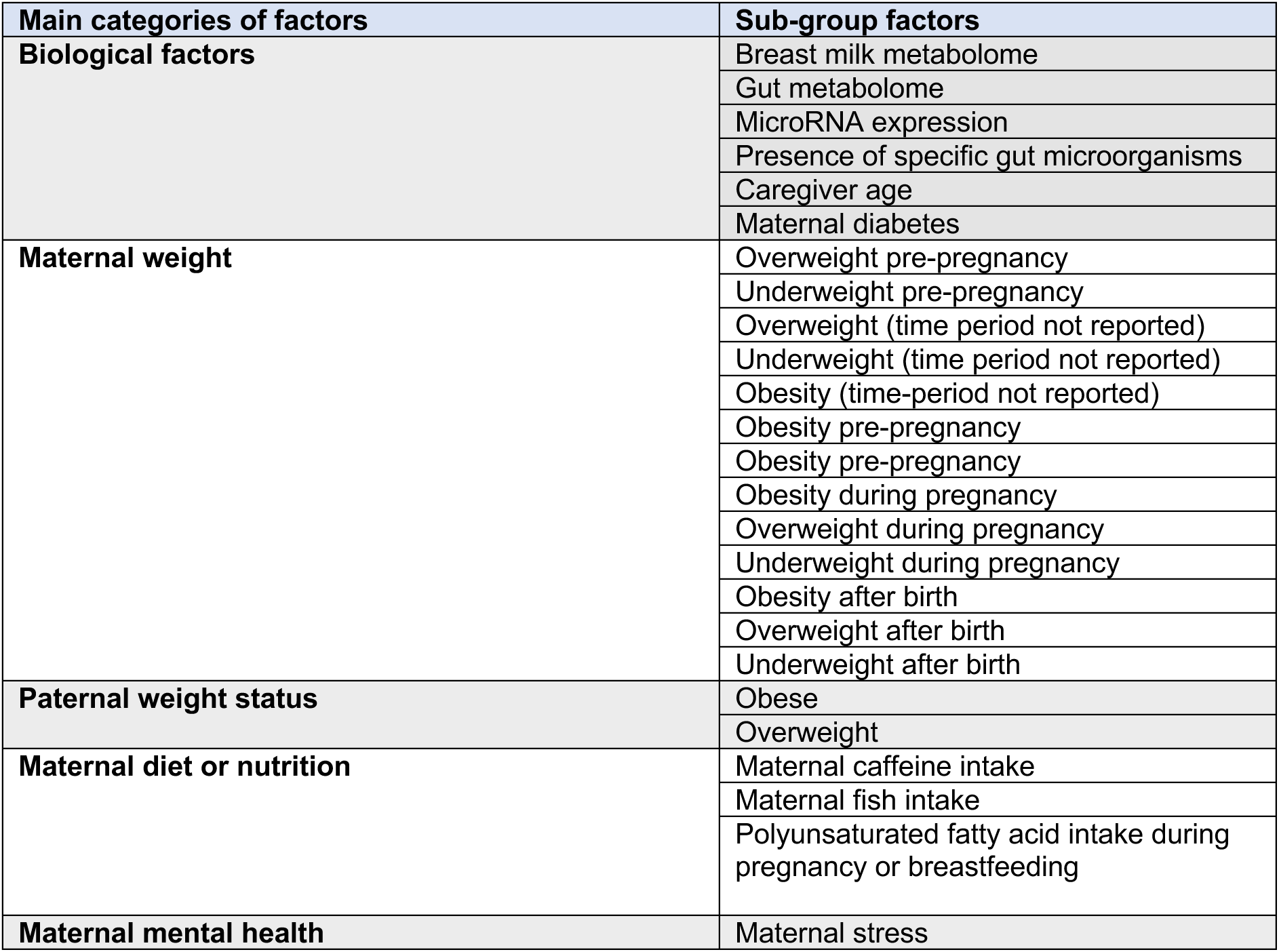
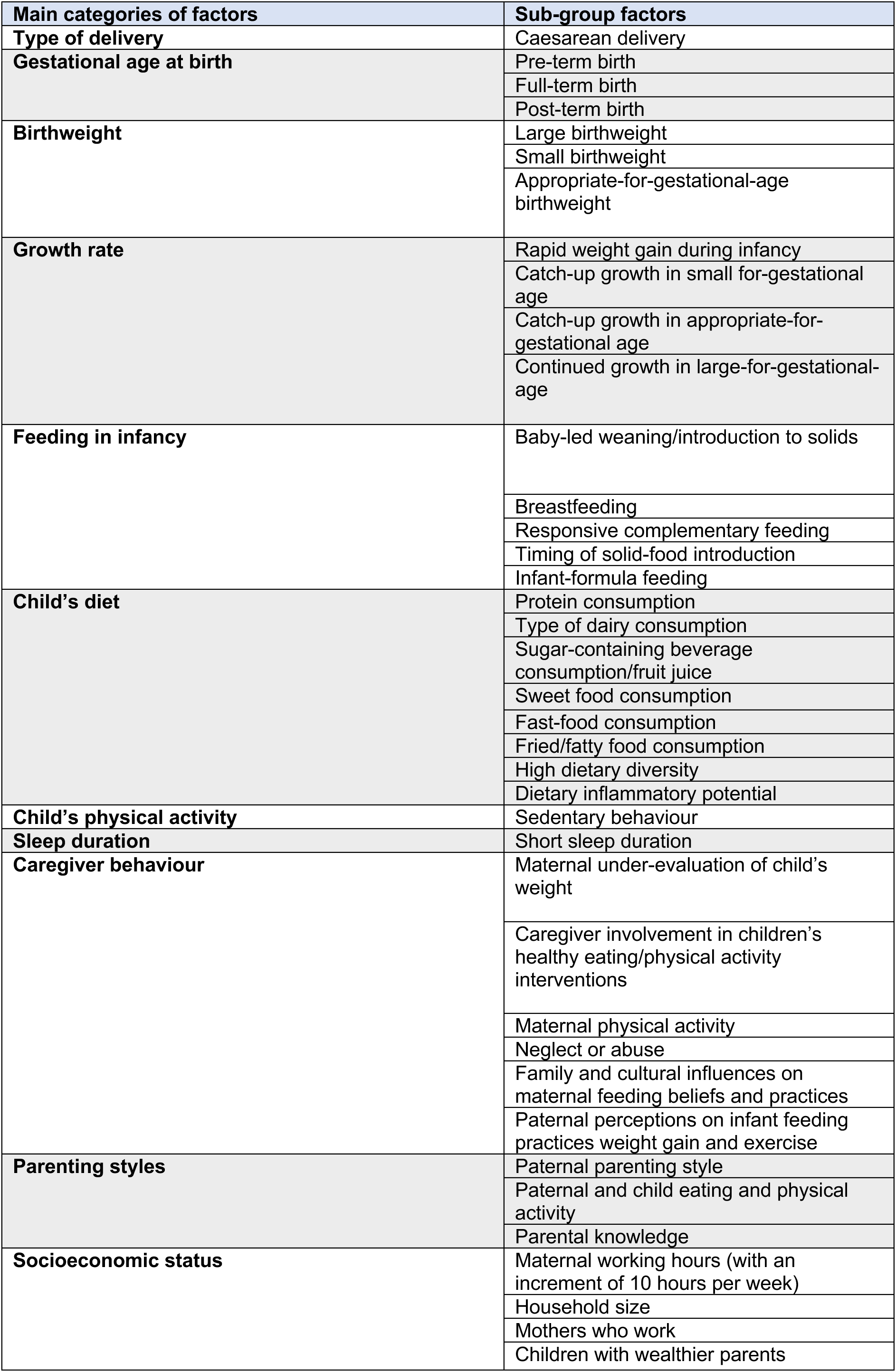
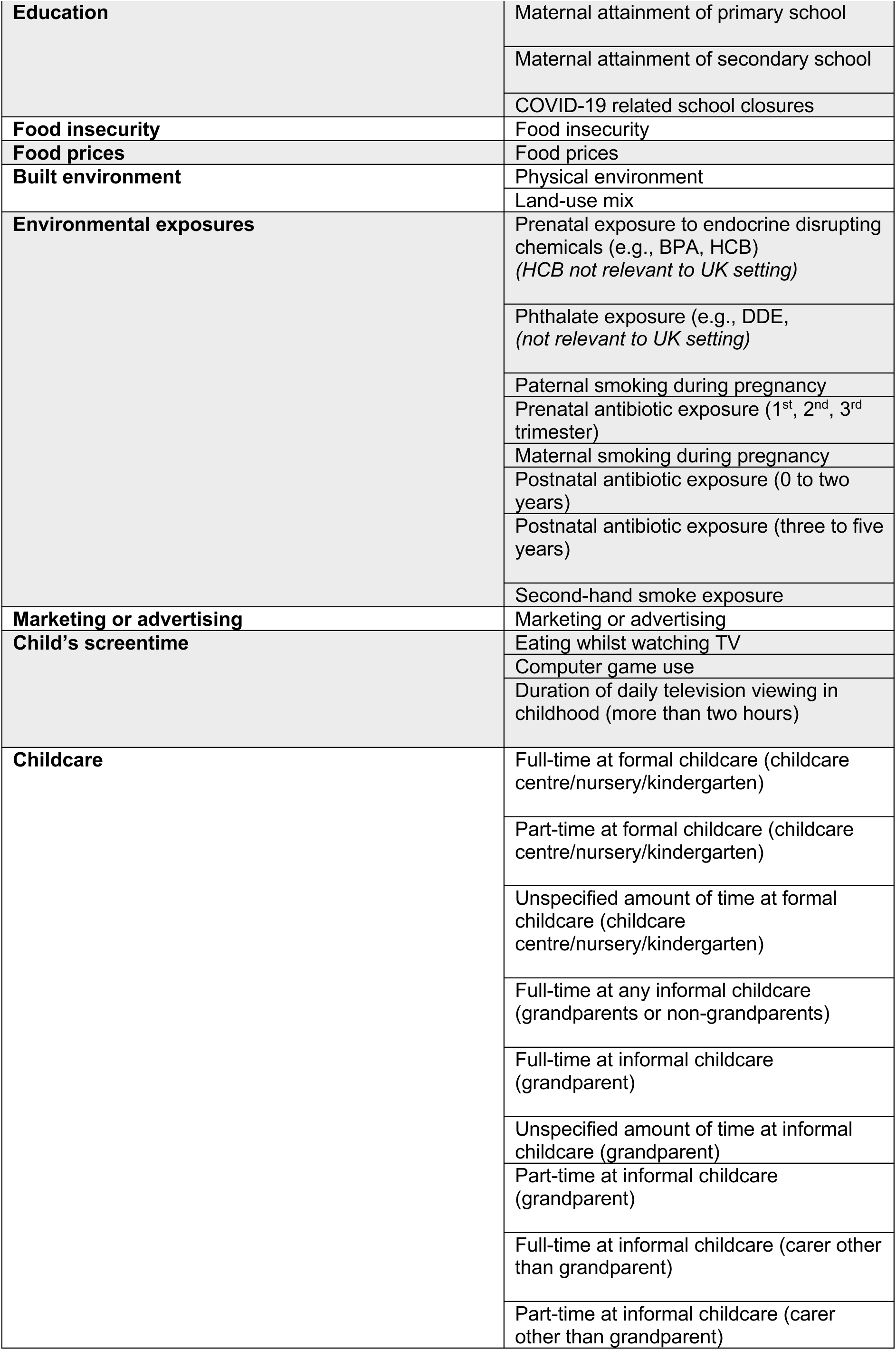
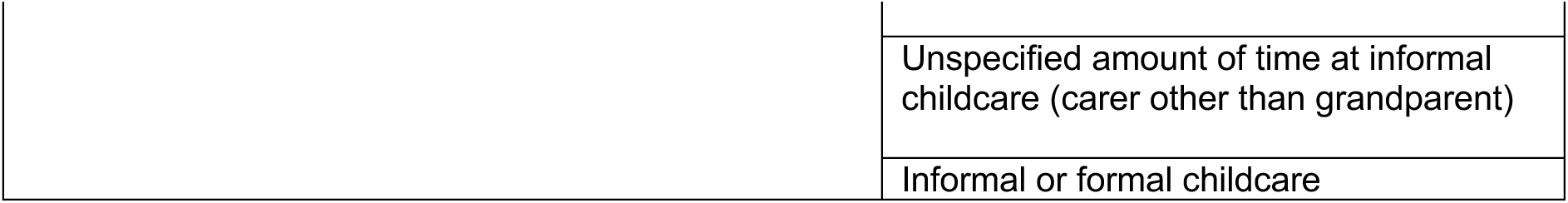
Sub-factors investigated in reviews as being potentially associated with childhood obesity or overweight.

A summary of the findings of the quantitative evidence along with the overall assessment of the certainty in the evidence are provided in Table 3. The findings were mainly judged to be of very low, low and moderate certainty.

**Table 3.**
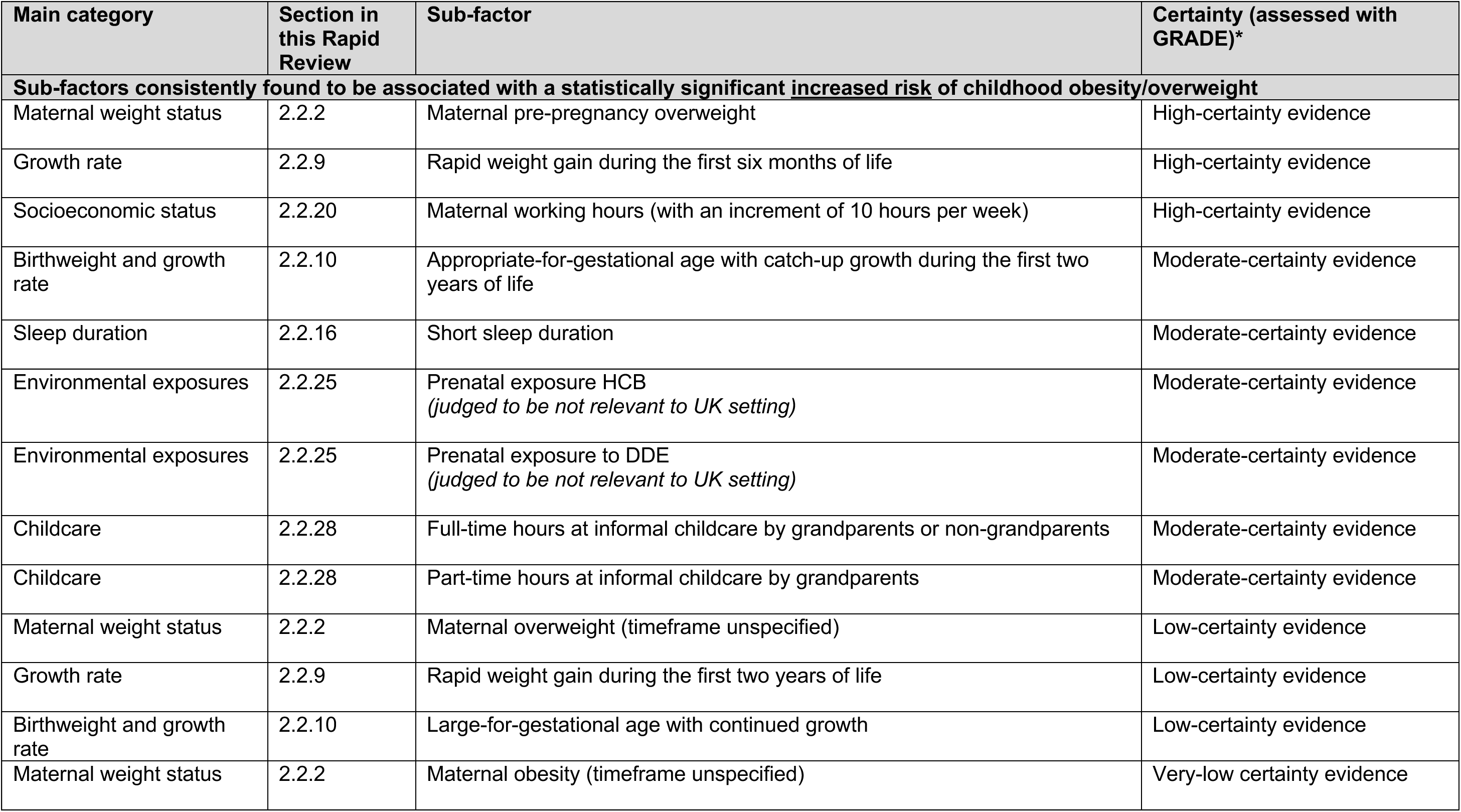
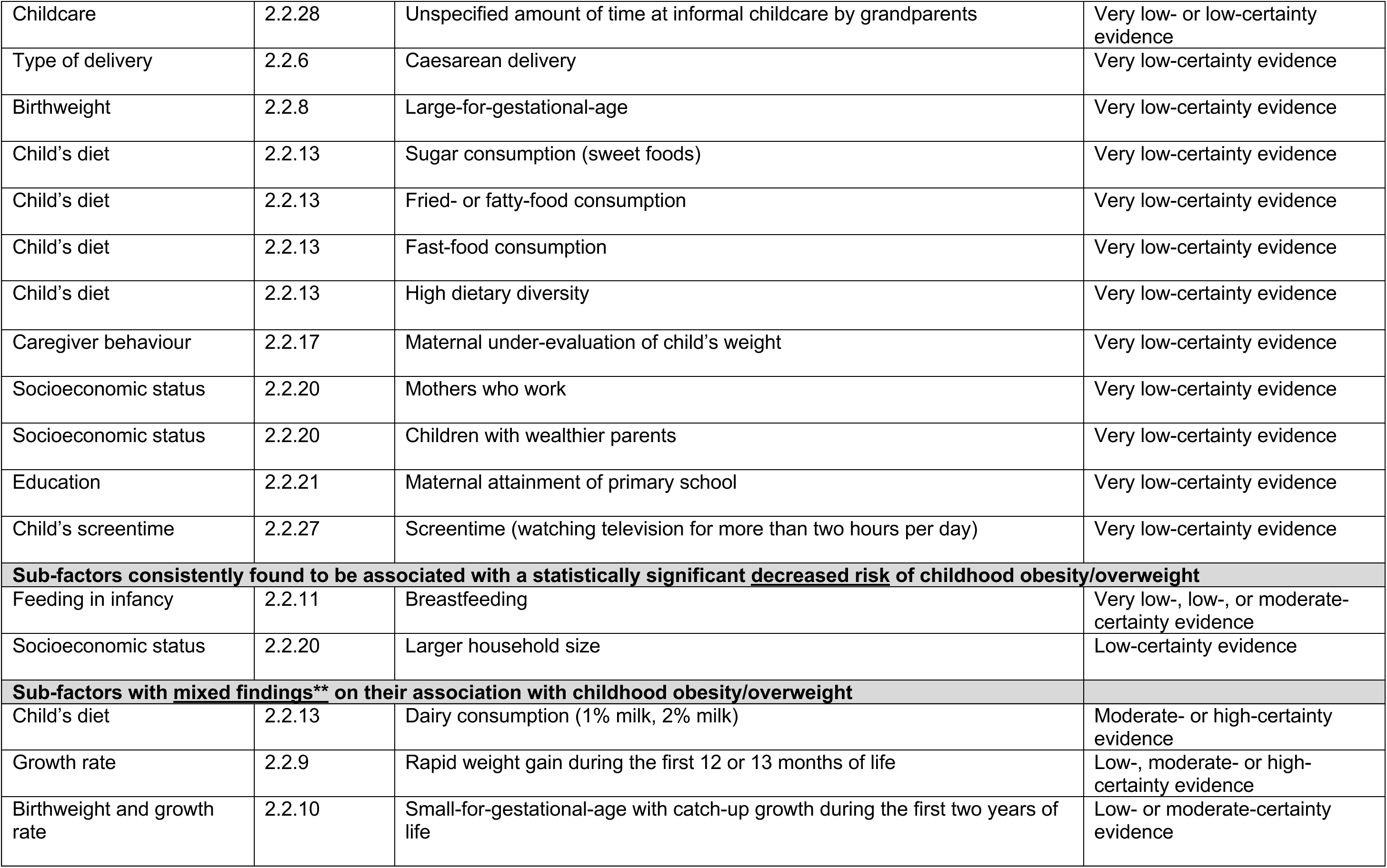
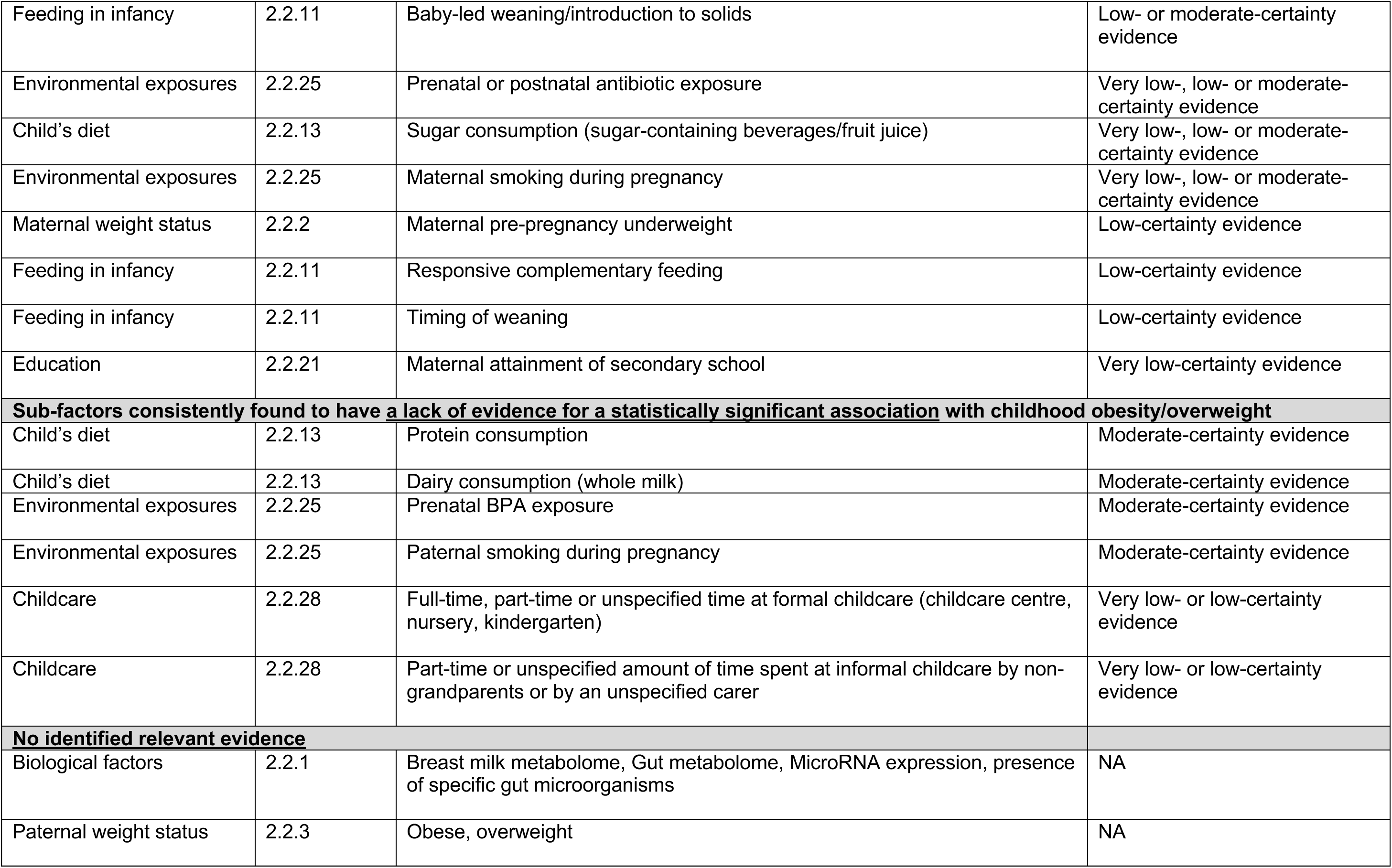
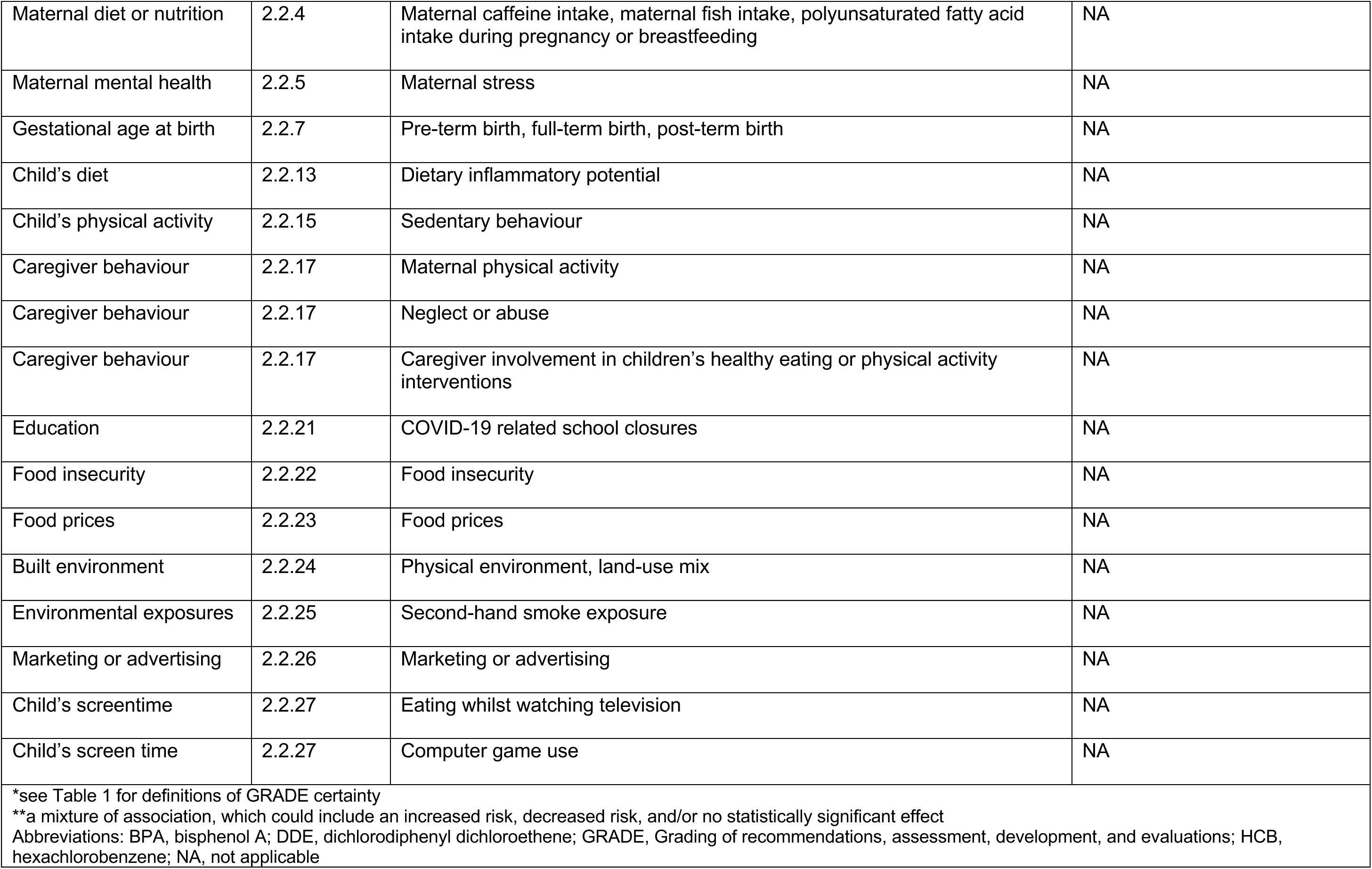
Summary of the findings and certainty of the quantitative evidence. Factors potentially associated with childhood obesity and/or overweight have been ordered under each header according to the level of certainty in the evidence, with factors with the highest certainty level evidence reported first

Details of the results for the quantitative evidence are available in Table 4 to Table 17. A summary of the design and characteristics of the studies in the reviews can be found in Section 6.2.

**Table 4:**
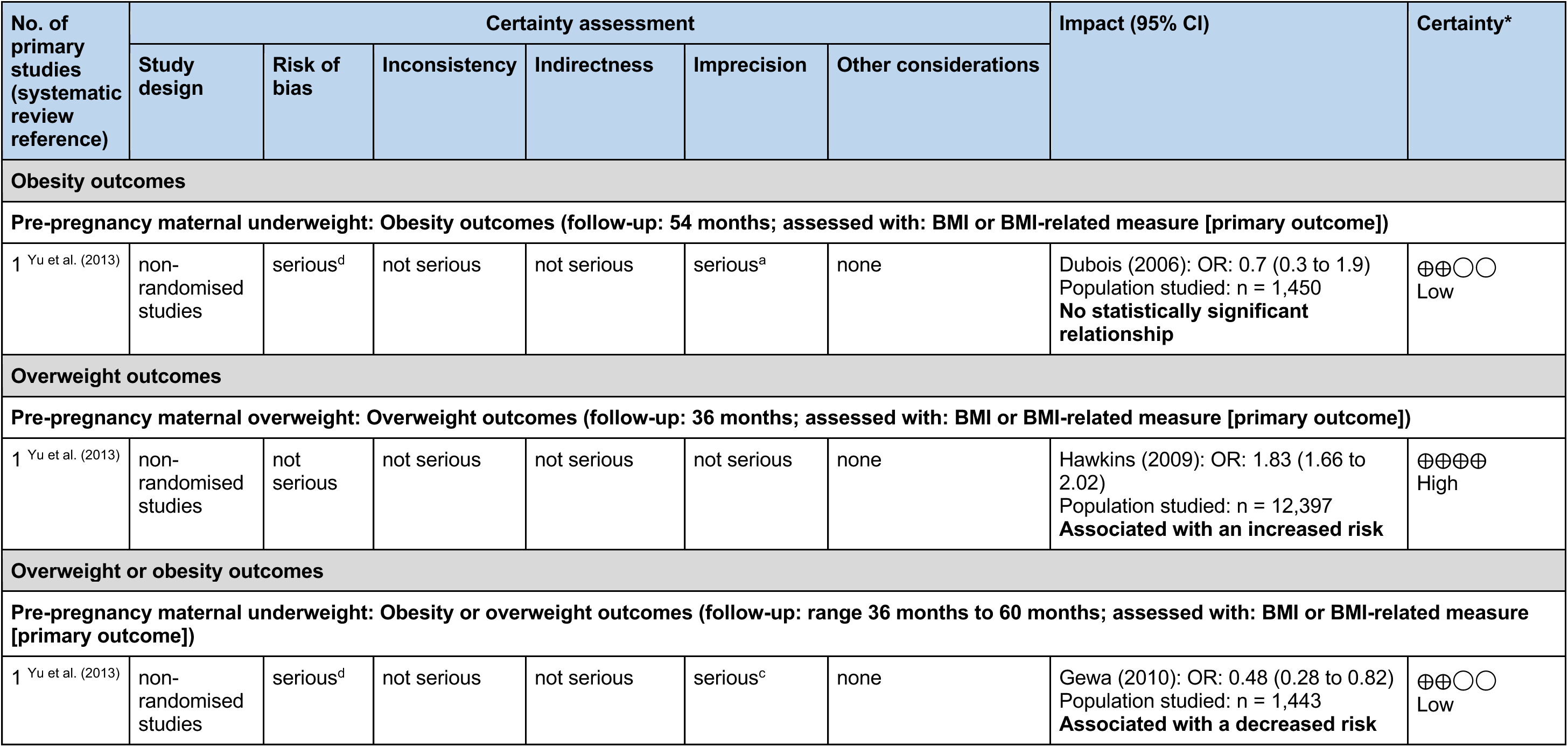
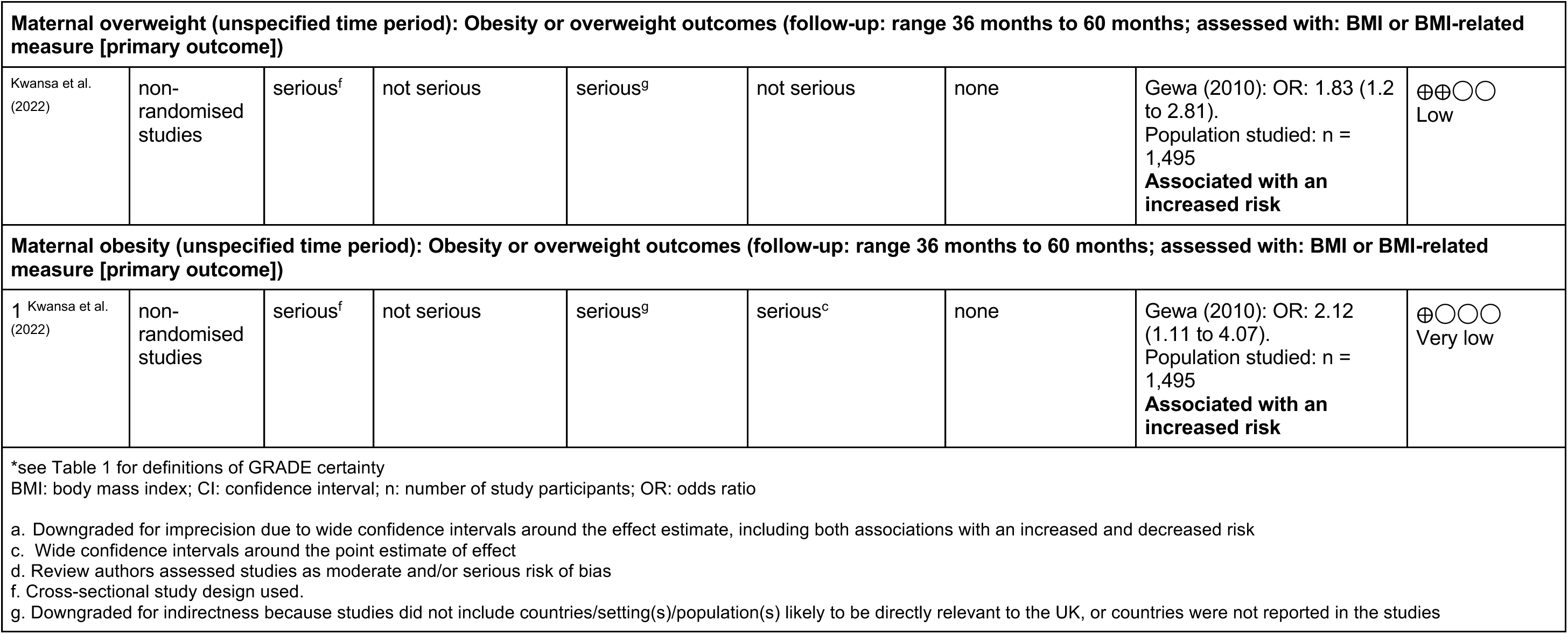
Summary of reviews reporting maternal weight status as a factor associated with childhood overweight or obesity: adapted from GRADE.

#### 2.1.2 Qualitative evidence

Two systematic reviews were identified that reported qualitative evidence syntheses (Cartagena et al. 2014, Fraser et al. 2011). Both systematic reviews explored factors that could contribute to overfeeding or weight gain among preschool children. Themes identified through completing a rapid qualitative evidence synthesis include **infant feeding practices**: breastfeeding and formula-feeding practices, family and cultural influences on maternal feeding beliefs and practices, paternal perceptions on infant-feeding practices, weight gain and exercise; and **parenting style**: paternal-parenting styles, paternal and child eating and physical activity, and parental knowledge.

Both systematic reviews referenced in our rapid qualitative evidence synthesis highlight the impact of cultural and familial influences on infant feeding practices across different demographics. Notably, this primary theme is closely related to the quantitative findings on feeding in infancy (Section 2.2.11), diet in childhood (Section 2.2.13) and caregiver behaviour (Section 2.2.18). A second main theme, parenting style, was less represented or related to any of the quantitative evidence on factors associated with childhood obesity or overweight.

A key limitation of this evidence is uncertainty in its applicability and generalisability to Wales or UK settings. Cartagena et al. (2014) only included studies of people of Hispanic ethnicity, and only three out of ten studies included in Fraser et al. (2011) were UK studies.

### 2.2 The association between factors and childhood obesity or overweight

We conducted a preliminary search of existing reviews to inform the current rapid review, and to develop our own classification system of potentially relevant factors (see Methods, Section 5.1 for details). As part of the initial preliminary search, we included any reviews reporting any factors associated with childhood obesity or overweight, or reporting factors as a possible association. We compiled this information from multiple reviews to determine as many potentially relevant factors to include in our own review as possible. Using this list, and building on the work of a previous review of existing reviews in this area (Monasta 2010), we developed a list of main categories of factors (Figure 3), and sub-groups of specific types of factor within each category (Table 2). Some of the reviews included in the initial preliminary search (and assessing certain factors) were subsequently excluded from the current, more focused, rapid review as they did not meet the final eligibility criteria (outlined in Section 5, Table 18). However, we kept all the factors identified within these reviews as part of the classification system we used within this rapid review. This helped us identify evidence gaps.

**Figure 3.**
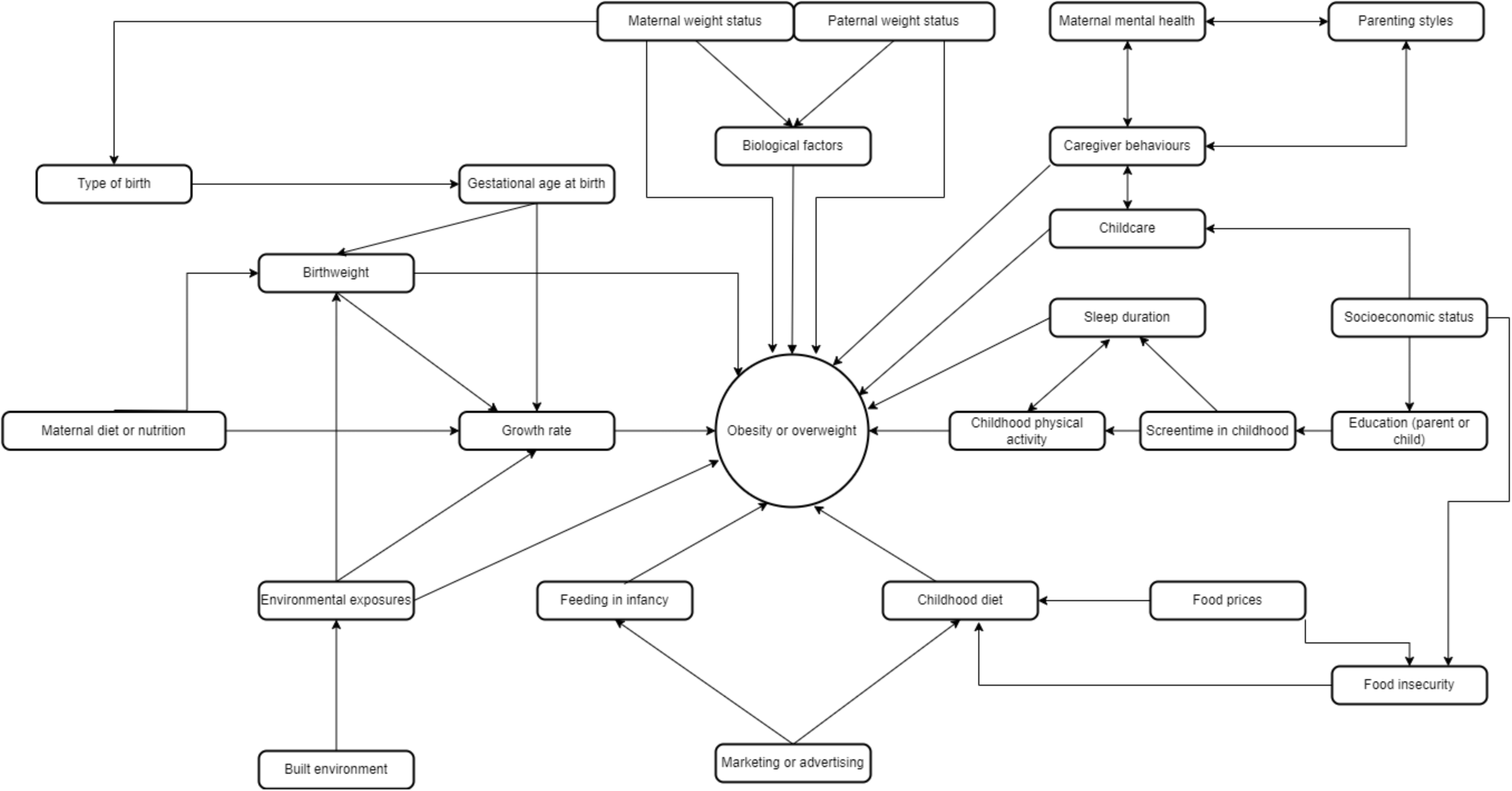
Factors potentially associated with childhood obesity or overweight, and their potential relationships with each other. Adapted from Monasta (2010)

Table 2 lists the main categories of factors, and also specific factors (sub-groups) within each category. Here, we list these in an approximate order of a child’s chronology.

Figure 3 shows the main categories of factors potentially associated with childhood obesity or overweight that we identified, and their potential relationships with each other.

An overall summary of the findings and certainty of the evidence for all factors identified in the reviews of quantitative evidence is provided in Table 3. The sections that follow report the specific findings for each of the main factors we identified.

#### 2.2.1 Biological factors

No relevant reviews were identified that reported biological factors (for example, breast milk metabolome, gut metabolome, microRNA expression, presence of specific gut micro-organisms, maternal age and maternal diabetes) and any associations with obesity or overweight in children less than five years old.

#### 2.2.2 Maternal weight status (quantitative evidence)

Two reviews examined the effect of maternal weight status as a factor for obesity or overweight in children less than five years old (Kwansa et al. 2022, Yu et al. 2013). A summary of the design and characteristics of the studies in the reviews can be found in Section 6.2, Table 19. Comprehensive details of the results for this section are available in Table 4.

##### 2.2.2.1 Maternal underweight

Two studies, in the review by Yu et al. (2013), reported mixed findings on the association between maternal pre-pregnancy underweight and childhood overweight and/or obesity (Dubois 2006, Gewa 2010). Maternal pre-pregnancy underweight was found to be associated with a decreased risk of childhood obesity or overweight in one study (Gewa 2010). In the other study by Dubois (2006), there was no statistically significant relationship between maternal pre-pregnancy underweight and childhood obesity. Both sets of results were judged to be low-certainty evidence.

##### 2.2.2.2 Maternal overweight/obesity

Two studies, in the reviews by Kwansa et al. (2022) and Yu et al. (2013), reported an association between maternal overweight and increased risk of childhood overweight (Hawkins 2009) (high-certainty evidence), and maternal overweight and childhood obesity or overweight (low-certainty evidence) (Gewa 2010). One study reported an association between maternal obesity and increased risk of childhood obesity or overweight (Gewa 2010) (very low-certainty evidence).

#### 2.2.3 Paternal weight status

No relevant reviews were identified that reported paternal weight status and any association with obesity or overweight in children less than five years old.

#### 2.2.4 Maternal diet or nutrition

No relevant reviews were identified that reported maternal diet or nutrition and any association with obesity or overweight in children less than five years old.

#### 2.2.5 Maternal mental health

No relevant reviews were identified that reported maternal mental health and any association with obesity or overweight in children less than five years old.

#### 2.2.6 Type of delivery (quantitative evidence)

Two reviews examined the effect of caesarean delivery compared to vaginal delivery as a factor for obesity in children less than five years old (Keag et al. 2018, Li et al. 2013). A summary of the design and characteristics of the studies in the reviews can be found in Section 6.2, Table 19. Comprehensive details of the results for this section are available in Table 5.

**Table 5:**
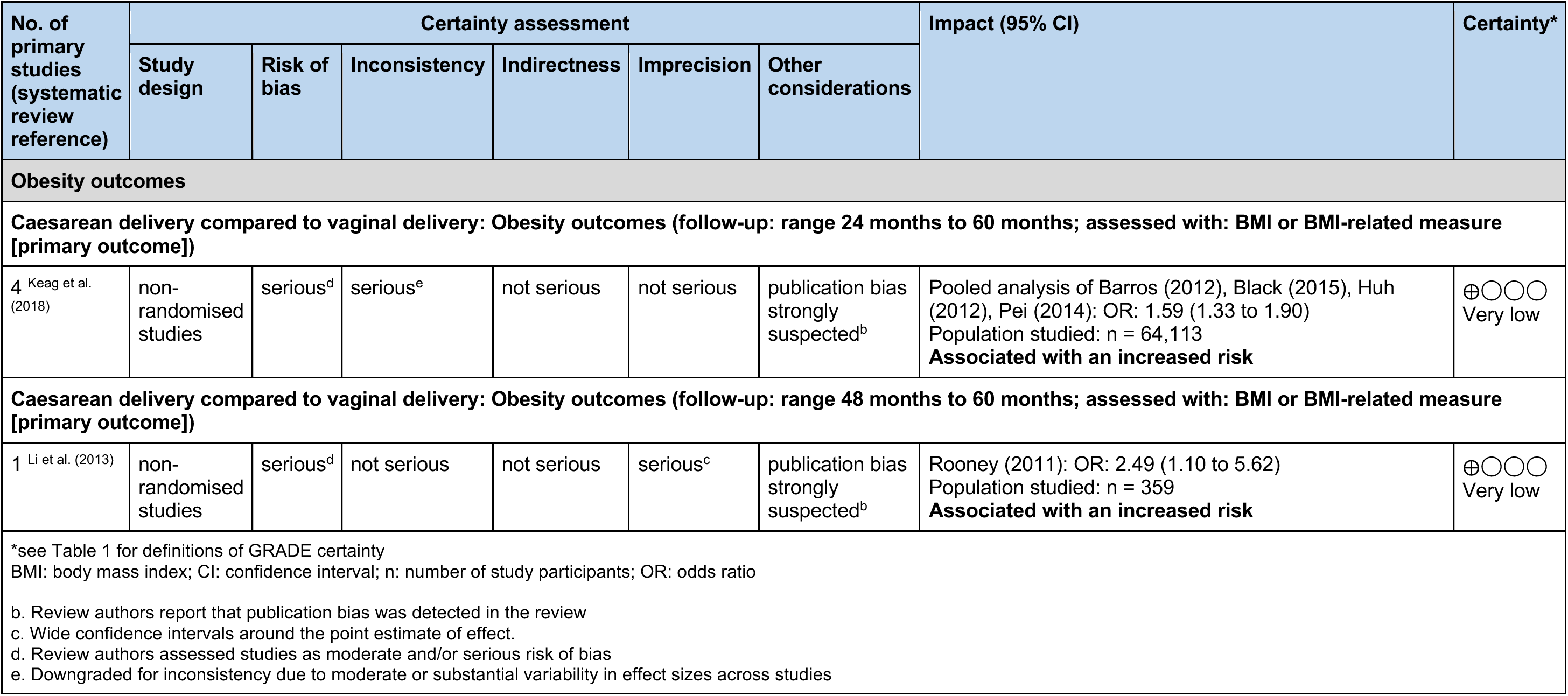
Summary of reviews reporting type of delivery as a factor associated with childhood overweight or obesity: adapted from GRADE.

##### 2.2.6.1 Caesarean section

Pooled analysis of four studies in the review by Keag et al. (2018) reported a positive association between caesarean section delivery and childhood obesity. An additional study by Rooney (2011), in the review by Li et al. (2013), also found an association between caesarean delivery and increased risk of childhood obesity. Both sets of results were judged to be very low-certainty evidence.

#### 2.2.7 Gestational age at birth

No relevant reviews were identified that reported gestational age at birth and any association with obesity or overweight in children less than five years old.

#### 2.2.8 Birthweight (quantitative evidence)

One review examined the effect of infants born large-for-gestational-age compared to those born appropriate-for-gestational-age as a factor for obesity or overweight in children less than five years old (Zhang et al. 2023). A summary of the design and characteristics of the studies in the reviews can be found in Section 6.2, Table 19. Comprehensive details of the results for this section are available in Table 6.

**Table 6:**
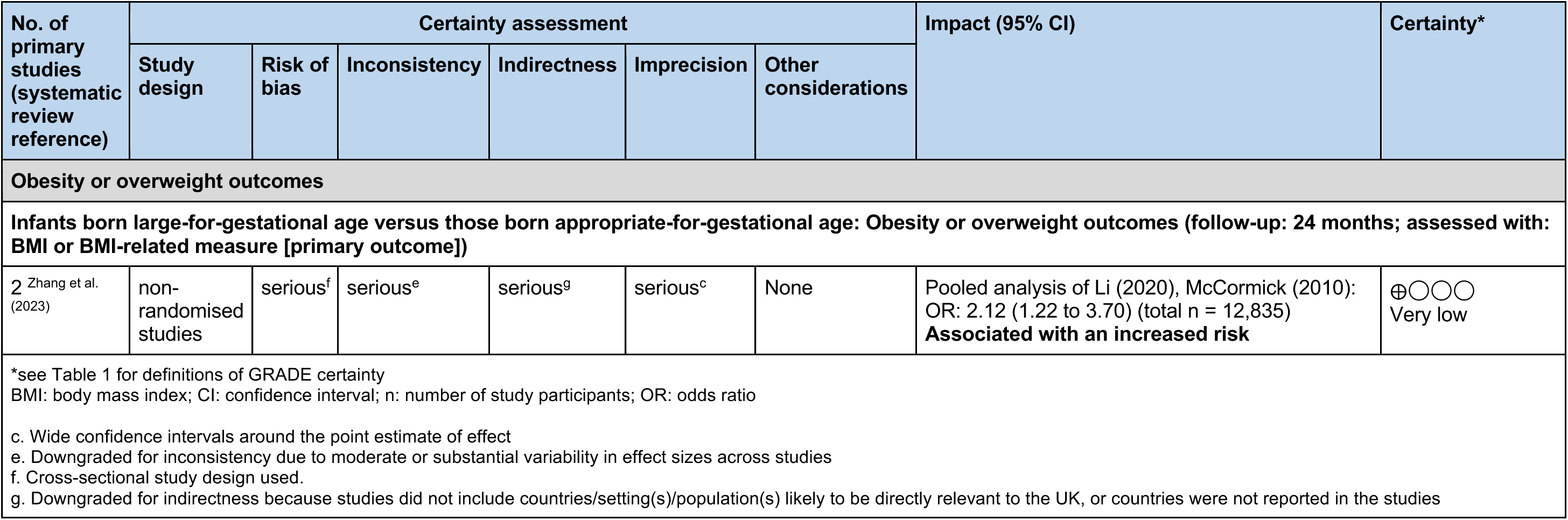
Summary of reviews reporting birthweight as a factor associated with childhood overweight or obesity: adapted from GRADE.

##### 2.2.8.1 Large-for-gestational-age

Pooled analysis of two studies in the review by Zhang et al. (2023) reported an association between infants born large-for-gestational-age and increased risk of childhood obesity or overweight outcomes (Zhang et al. 2023) (very low-certainty evidence)

##### 2.2.8.2 Small-for-gestational age

No relevant reviews were identified that reported small-for-gestational-age and its association with obesity or overweight in children less than five years old.

##### 2.2.8.3 Appropriate-for-gestational age

No relevant reviews were identified that reported appropriate-for-gestational-age and its association with obesity or overweight in children less than five years old.

#### 2.2.9 Growth rate (quantitative evidence)

Two reviews examined the effect of rapid weight gain during the first two years of life as a factor for obesity or overweight in children less than five years old (Andrea et al. 2017, Zheng et al. 2018). A summary of the design and characteristics of the studies in the reviews can be found in Section 6.2, Table 19. Comprehensive details of the results for this section are available in Table 7.

**Table 7:** Summary of reviews reporting growth rate as a factor associated with childhood overweight or obesity: adapted from GRADE.

| No. of primary studies<br>(systematic review reference) | Certainty assessment |  |  |  |  |  | Impact (95% CI) | Certainty* |
| --- | --- | --- | --- | --- | --- | --- | --- | --- |
|  | Study design | Risk of bias | Inconsistency | Indirectness | Imprecision | Other considerations |  |  |
| Obesity outcomes |  |  |  |  |  |  |  |  |
| Rapid weight gain at 0 to 12 months: Obesity outcomes (follow-up: range 24 to 38; assessed with: BMI or BMI-related measure [primary outcome]) |  |  |  |  |  |  |  |  |
| 1 <sup>Andrea et al. (2017)</sup> | non-randomised studies | not serious | not serious | not serious | serious<sup>c</sup> | none | Goodell (2009): OR: 9.24 (3.73 to 22.91)<br>Population studied: n = 203<br>**Associated with an increased risk** | ⊕⊕⊕○<br>Moderate |
| Rapid weight gain at 0 to 13 months: Obesity outcomes (follow-up: 48 months; assessed with: BMI or BMI-related measure [primary outcome]) |  |  |  |  |  |  |  |  |
| 1 <sup>Andrea et al. (2017)</sup> | non-randomised studies | not serious | not serious | not serious | serious<sup>a</sup> | none | Roy (2015): OR: 0.85 (0.10 to 7.20)<br>Population studied: n = 635<br>**No statistically significant relationship** | ⊕⊕⊕○<br>Moderate |
| Overweight outcomes |  |  |  |  |  |  |  |  |
| Rapid weight gain at 0 to 6 months: Overweight outcomes (follow-up: range 48 months to 60 months; assessed with: BMI or BMI-related measure [primary outcome]) |  |  |  |  |  |  |  |  |
| 1 <sup>Andrea et al. (2017)</sup> | non-randomised studies | not serious | not serious | not serious | not serious | none | Stettler (2002): OR: 1.17 (1.11 to 1.24)<br>Population studied: n = 12,277<br>**Associated with an increased risk** | ⊕⊕⊕⊕<br>High |
| Rapid weight gain at 0 to 12 months: Overweight outcomes (follow-up: range 24 months to 36 months; assessed with: BMI or BMI-related measure [primary outcome]) |  |  |  |  |  |  |  |  |
| 1 <sup>Andrea et al. (2017)</sup> | non-randomised studies | not serious | not serious | not serious | serious<sup>a</sup> | none | Goodell (2009): OR: 2.06 (0.86 to 4.92)<br>Population studied: n = 203<br>**No statistically significant relationship** | ⊕⊕⊕○<br>Moderate |
| Rapid weight gain at 0 to 12 months: Overweight outcomes (follow-up: 36 months; assessed with: BMI or BMI-related measure [primary outcome]) |  |  |  |  |  |  |  |  |
| 1 <sup>Zheng et al. (2018)</sup> | non-randomised studies | not serious | not serious | not serious | not serious | none | Weng (2013): OR: 4.15 (3.64 to 4.73)<br>Population studied: n = 18,296<br>**Associated with an increased risk** | ⊕⊕⊕⊕<br>High |
|  | Study design | Risk of bias | Inconsistency | Indirectness | Imprecision | Other considerations |  |  |
| Obesity or overweight outcomes |  |  |  |  |  |  |  |  |
| Rapid weight gain at 0 to 12 months: Obesity or overweight outcomes (follow-up: 24 months; assessed with: BMI or BMI-related measure [primary outcome]) |  |  |  |  |  |  |  |  |
| 1 Zheng et al. (2018) | non-randomised studies | serious<sup>d</sup> | not serious | not serious | serious<sup>c</sup> | none | Webster (2013): OR: 5.22 (2.33 to 11.71)<br>Population studied: n = 157<br>**Associated with an increased risk** | ⊕⊕○○<br>Low |
| Rapid weight gain at 0 to 24 months: Obesity or overweight outcomes (follow-up: range 36 months to 60 months; assessed with: BMI or BMI-related measure [primary outcome]) |  |  |  |  |  |  |  |  |
| 3 Zheng et al. (2018) | non-randomised studies | not serious | not serious | serious<sup>g</sup> | serious<sup>c</sup> | none | Heppe (2013): OR: 6.39 (4.54 to 8.99);<br>Rathnayake KM (2013): OR: 6.29 (2.04 to 19.49);<br>Taal (2013): OR: 3.11 (2.37 to 4.08)<br>Population studied: n = 7,693<br>**Associated with an increased risk** | ⊕⊕○○<br>Low |
| *see Table 1 for definitions of GRADE certainty<br>BMI: body mass index; CI: confidence interval; n: number of study participants; OR: odds ratio |  |  |  |  |  |  |  |  |
| a. Downgraded for imprecision due to wide confidence intervals around the effect estimate, including both being associated with an increased and decreased risk<br>c. Wide confidence intervals around the point estimate of effect<br>d. Review authors assessed studies as moderate and/or serious risk of bias<br>g. Downgraded for indirectness because studies did not include countries/setting(s)/population(s) likely to be directly relevant to the UK, or countries were not reported in the studies |  |  |  |  |  |  |  |  |

##### 2.2.9.1 Rapid weight gain during first six months of life

One study in the review by Andrea et al. (2017) reported that rapid weight gain from birth to six months was associated with an increased risk of childhood overweight (Stettler 2002) (high-certainty evidence).

##### 2.2.9.2 Rapid weight gain during first 12 or 13 months of life

Four studies in the reviews by Andrea et al. (2017) and Zheng et al. (2018) reported mixed findings for the association between rapid weight gain from birth to 12 or 13 months and obesity and/or overweight (Goodell 2009, Roy 2015, Webster 2013, Weng 2013). Two of these studies reported an association between rapid weight gain from birth to 12 months and increased risk of overweight (Weng 2013) (high-certainty evidence), or obesity or overweight (Webster 2013) (low-certainty evidence). Two studies reported no statistically significant relationship between rapid weight gain at birth to 13 months and childhood obesity (Roy 2015) (moderate-certainty evidence) or childhood overweight (Goodell 2009) (moderate-certainty evidence).

##### 2.2.9.3 Rapid weight gain during first 24 months of life

Three studies in the review by Zheng et al. (2018) all reported an association between rapid weight gain from birth to 24 months and increased risk of childhood obesity or overweight (Heppe 2013, Rathnayake KM 2013, Taal 2013). The results from these studies were judged as low-certainty evidence.

#### 2.2.10 Birthweight and growth rate (quantitative evidence)

Two reviews examined the effect of birthweight and growth rate during the first two years of life as a factor for obesity or overweight in children less than five years old (Halilagic & Moschonis 2021, Matthews et al. 2017). A summary of the design and characteristics of the studies in the reviews can be found in Section 6.2, Table 19. Comprehensive details of the results for this section are available in Table 8.

**Table 8:** Summary of reviews reporting birthweight and growth rate as a factor associated with childhood overweight or obesity: adapted from GRADE.

| No. of primary studies (systematic review reference) | Certainty assessment |  |  |  |  |  | Impact (95% CI) | Certainty* |
| --- | --- | --- | --- | --- | --- | --- | --- | --- |
|  | Study design | Risk of bias | Inconsistency | Indirectness | Imprecision | Other considerations |  |  |
| Obesity or overweight |  |  |  |  |  |  |  |  |
| Small-for-gestational age with catch-up growth (birth to 2 years) compared to those appropriate-for-gestational-age with normal growth: Obesity or overweight outcomes (follow-up: range 24 months to 48 months; assessed with: BMI or BMI-related measure [primary outcome]) |  |  |  |  |  |  |  |  |
| 1 Matthews et al. (2017) | non-randomised studies | serious<sup>d</sup> | not serious | not serious | serious<sup>a</sup> | none | Taal (2013): OR: 0.92 (0.43 to 1.96)<br>Population studied: n = 3,941<br>**No statistically significant relationship** | ⊕⊕○○<br>Low |
| Small-for-gestational age with catch-up growth: Obesity or overweight outcomes (follow-up: range 24 months to 60 months; assessed with: BMI or BMI-related measure [primary outcome]) |  |  |  |  |  |  |  |  |
| 1 Halilagic & Moschonis (2021) | non-randomised studies | not serious | not serious | serious<sup>g</sup> | not serious | none | Shi HuiQing (2018): OR: 2.3 (1.8 to 3.0)<br>Population studied: n = 3,004<br>**Associated with an increased risk** | ⊕⊕⊕○<br>Moderate |
| Small-for-gestational age with excessive-rapid catch-up growth: Obesity or overweight outcomes (follow-up: range 24 months to 60 months; assessed with: BMI or BMI-related measure [primary outcome]) |  |  |  |  |  |  |  |  |
| 1 Halilagic & Moschonis (2021) | non-randomised studies | not serious | not serious | serious<sup>g</sup> | not serious | none | Shi HuiQing (2018): OR: 11.6 (8.8 to 15.3)<br>Population studied: n = 3,004<br>**Associated with an increased risk** | ⊕⊕⊕○<br>Moderate |
| Appropriate-for-gestational-age with catch-up-growth [birth to 2 years] compared to those appropriate-for-gestational-age with normal growth: Obesity or overweight outcomes (follow-up: range 24 months to 48 months; assessed with: BMI or BMI-related measure [primary outcome]) |  |  |  |  |  |  |  |  |
| 1 Halilagic & Moschonis (2021),<br>Matthews et al. (2017) | non-randomised studies | serious<sup>d</sup> | not serious | not serious | not serious | none | Taal (2013): OR: 3.11 (2.37 to 4.08)<br>Population studied: n = 3,941<br>**Associated with an increased risk** | ⊕⊕⊕○<br>Moderate |
|  | Study design | Risk of bias | Inconsistency | Indirectness | Imprecision | Other considerations |  |  |
| Large-for-gestational age with continued growth (birth to 2 years) compared to those appropriate-for-gestational-age with normal growth (follow-up: range 24 months to 48 months; assessed with: BMI or BMI-related measure [primary outcome]) |  |  |  |  |  |  |  |  |
| 1 Matthews et al. (2017) | non-randomised studies | serious<sup>d</sup> | not serious | not serious | serious<sup>c</sup> | none | Taal (2013): OR: 12.46 (6.07 to 25.58)<br>Population studied: n = 3,941<br>**Associated with an increased risk** | ⊕⊕○○<br>Low |
| *see Table 1 for definitions of GRADE certainty<br>BMI: body mass index; CI: confidence interval; n: number of study participants; OR: odds ratio<br>a. Downgraded for imprecision due to wide confidence intervals around the effect estimate, including both being associated with an increased or decreased risk<br>c. Wide confidence intervals around the point estimate of effect.<br>d. Review authors assessed studies as moderate and/or serious risk of bias<br>g. Downgraded for indirectness because studies did not include countries/setting(s)/population(s) likely to be directly relevant to the UK, or countries were not reported in the studies |  |  |  |  |  |  |  |  |

##### 2.2.10.1 Small-for-gestational-age with catch-up growth

Two studies in the reviews by Halilagic & Moschonis (2021) and Matthews et al. (2017) reported mixed findings on the association between small-for-gestational age with catch-up growth during the first two years of life and childhood obesity or overweight (Shi HuiQing 2018, Taal 2013). Catch-up growth was defined as an increase in weight-for-age z-score greater than 0.67 standard deviation units in the first two years of life. One of the studies reported no statistically significant relationship between small-for-gestational-age with catch-up growth and childhood obesity or overweight (Taal 2013) (low-certainty evidence). The other study reported an association between small-for-gestational-age with catch-up growth or excessive-rapid catch-up growth and increased risk childhood obesity or overweight (Shi HuiQing 2018) (moderate-certainty evidence).

##### 2.2.10.2 Appropriate-for-gestational-age with catch-up growth

In the reviews by Halilagic & Moschonis (2021) and Matthews et al. (2017), the study by Taal (2013) reported an association between being born appropriate-for-gestational-age with catch-up growth during the first two years of life and increased risk of childhood obesity or overweight, compared to those born appropriate-for-gestational-age with normal growth (moderate-certainty evidence).

##### 2.2.10.3 Large-for-gestational-age with continued growth

In the review by Matthews et al. (2017), the study by Taal (2013) reported an association between being born large-for-gestational-age with continued growth during the first two years of life and increased risk of childhood obesity or overweight, compared to those born appropriate-for-gestational-age with normal growth (low-certainty evidence).

#### 2.2.11 Feeding in infancy (quantitative evidence)

Five reviews examined the effect of feeding in infancy as a factor for obesity or overweight in children less than five years old (Bergamini et al. 2022, Martinon-Torres et al. 2021, Moorcroft et al. 2011, Verga et al. 2022, Weng et al. 2012). A summary of the design and characteristics of the studies in the reviews can be found in Section 6.2, Table 19.

Comprehensive details of the results for this section are available in Table 9.

**Table 9:** Summary of reviews reporting feeding in infancy as a factor associated with childhood overweight or obesity: adapted from GRADE.

| No. of primary studies (systematic review reference) | Certainty assessment |  |  |  |  |  | Impact (95% CI) | Certainty |
| --- | --- | --- | --- | --- | --- | --- | --- | --- |
|  | Study design | Risk of bias | Inconsistency | Indirectness | Imprecision | Other considerations |  |  |
| Overweight outcomes |  |  |  |  |  |  |  |  |
| Baby-led weaning (≤10% spoon-feeding and purees) versus traditional spoon-feeding: Overweight outcomes (follow-up: range 18 months to 24 months; assessed with: Weight and/or length measures [primary outcome]) |  |  |  |  |  |  |  |  |
| 1 Martinon-Torres et al. (2021) | non-randomised studies | serious<sup>d</sup> | not serious | not serious | not serious | none | Brown (2015): prevalence: 8.1% (baby-led weaning); 19.2% (traditional spoon feeding)<br>Population studied: n = 298<br>p-value not reported but review states traditional spoon-feeding is associated with significantly higher prevalence<br>**Associated with a decreased risk** | ⊕⊕⊕○<br>Moderate |
| Introduction of solid foods before 4 months versus introduction of solid food after 4 months: Overweight outcomes (follow-up: 36 months; assessed with: BMI or BMI-related measure [primary outcome]) |  |  |  |  |  |  |  |  |
| 1 Moorcroft et al. (2011), Weng et al. (2012) | non-randomised studies | serious<sup>d</sup> | not serious | not serious | not serious | publication bias strongly suspected<sup>b</sup> | Hawkins (2009): OR: 1.12 (1.02 to 1.23)<br>Population studied: n = 13,188<br>**Associated with an increased risk** | ⊕⊕○○<br>Low |
| Whether infant was ever breastfed versus never breastfed: Overweight outcomes (follow-up: 24 months; assessed with: BMI or BMI-related measure [primary outcome]) |  |  |  |  |  |  |  |  |
| 1 Weng et al. (2012) | non-randomised studies | serious<sup>d</sup> | not serious | not serious | serious<sup>a</sup> | publication bias strongly suspected<sup>b</sup> | Weyermann (2006): OR: 2.20 (0.70 to 7.20)<br>Population studied: n = 855<br>**No statistically significant relationship** | ⊕○○○<br>Very low |
| Whether infant was ever breastfed versus never breastfed: Overweight outcomes (follow-up: 36 months; assessed with: BMI or BMI-related measure [primary outcome]) |  |  |  |  |  |  |  |  |
| 2 Weng et al. (2012) | non-randomised studies | serious<sup>d</sup> | not serious | not serious | not serious | publication bias strongly suspected<sup>b</sup> | Hawkins (2009): OR: 0.86 (0.76 to 0.97); Taveras (2006): OR: 0.34 (0.14 to 0.87)<br>Population studied: n = 14,160 | ⊕⊕○○<br>Low |

| No. of primary studies (systematic review reference) | Certainty assessment |  |  |  |  |  | Impact (95% CI) | Certainty* |
| --- | --- | --- | --- | --- | --- | --- | --- | --- |
|  | Study design | Risk of bias | Inconsistency | Indirectness | Imprecision | Other considerations |  |  |
|  |  |  |  |  |  |  | Associated with a decreased risk |  |
| **Whether infant was ever breastfed versus never breastfed: Overweight outcomes (follow-up: 42 months; assessed with: BMI or BMI-related measure [primary outcome])** |  |  |  |  |  |  |  |  |
| 1 Weng et al. (2012) | non-randomised studies | not serious | not serious | not serious | not serious | publication bias strongly suspected<sup>b</sup> | Armstrong (2002): OR: 0.72 (0.65 to 0.79)<br>Population studied: n = 32,200<br>**Associated with a decreased risk** | ⊕⊕⊕○<br>Moderate |
| **Whether infant was ever breastfed versus never breastfed: Overweight outcomes (follow-up: 48 months; assessed with: BMI or BMI-related measure [primary outcome])** |  |  |  |  |  |  |  |  |
| 1 Weng et al. (2012) | non-randomised studies | serious<sup>d</sup> | not serious | not serious | not serious | publication bias strongly suspected<sup>b</sup> | Grummer-Strawn (2004): OR: 0.72 (0.65 to 0.80)<br>Population studied: n = 12,587<br>**Associated with a decreased risk** | ⊕⊕○○<br>Low |
| **Obesity or overweight outcomes** |  |  |  |  |  |  |  |  |
| **Baby-led weaning/baby-led introduction to solids compared to other models of complementary feeding: Obesity or overweight outcomes (follow-up: range 12 months to 24 months; assessed with: Multiple primary outcomes measures)** |  |  |  |  |  |  |  |  |
| 2 Bergamini et al. (2022) | randomised trials | serious<sup>d</sup> | not serious | not serious | serious<sup>a</sup> | none | Pooled analysis of 2 RCTs (Dogan 2018, Taylor 2017) (n = 457): RR: 0.15 (0.00 to 17.79)<br>**No statistically significant relationship** | ⊕⊕○○<br>Low |
| **Responsive complementary feeding compared to other models of complementary feeding: Obesity or overweight outcomes (follow-up: 12 months; assessed with: Weight and/or length measures [primary outcome])** |  |  |  |  |  |  |  |  |
| 1 Bergamini et al. (2022) | randomised trials | serious<sup>d</sup> | not serious | not serious | serious<sup>a</sup> | none | Savage (2016): RR: 0.44 (0.19 to 1.03, p = 0.06).<br>Population studied: n = 250<br>**No statistically significant relationship** | ⊕⊕○○<br>Low |
| **Responsive complementary feeding compared to other models of complementary feeding: Obesity or overweight outcomes (follow-up: range 24 months to 36 months; assessed with: BMI or BMI-related measure [primary outcome])** |  |  |  |  |  |  |  |  |
| 2 Bergamini et al. (2022) | Randomised and non- | serious<sup>d</sup> | not serious | not serious | serious<sup>c</sup> | none | Pooled analysis of 1 RCT (Paul 2018) and 1 observational study (Machuca 2016) (n = 466): RR: 0.37 (0.21 to 0.64) | ⊕⊕○○<br>Low |
|  | Study design | Risk of bias | Inconsistency | Indirectness | Imprecision | Other considerations |  |  |
|  | randomised studies |  |  |  |  |  | Associated with a decreased risk |  |
| **Introduction of complementary feeding at 4 to 6 months versus introduction of complementary feeding at 6 months: Obesity or overweight outcomes (follow-up: 18 months; assessed with: Weight and/or length measures [primary outcome])** |  |  |  |  |  |  |  |  |
| 1 Verga et al. (2022) | randomised trials | serious<sup>d</sup> | not serious | not serious | serious<sup>a</sup> | none | Jonsdottir (2014): RR: 1.30 (0.37 to 4.56)<br>Population studied: n = 94<br>**No statistically significant relationship** | ⊕⊕○○<br>Low |
| **Exclusively breastfed infants who received complementary feeding between 4 and 6 months versus complementary feeding at 6 months: Obesity or overweight outcomes (follow-up: 36 months; assessed with: BMI or BMI-related measure [primary outcome])** |  |  |  |  |  |  |  |  |
| 1 Verga et al. (2022) | non-randomised studies | serious<sup>d</sup> | not serious | not serious | serious<sup>a</sup> | none | Huh (2011): RR: 0.80 (0.51 to 1.23)<br>Population studied: n = 525<br>**No statistically significant relationship** | ⊕⊕○○<br>Low |
| **Formula-fed infants who received complementary feeding at 4 to 5 months versus complementary feeding at ≥ 6 months: Obesity or overweight outcomes (follow-up: 36 months; assessed with: BMI or BMI-related measure [primary outcome])** |  |  |  |  |  |  |  |  |
| 1 Verga et al. (2022) | non-randomised studies | serious<sup>d</sup> | not serious | not serious | serious<sup>a</sup> | none | Huh (2011): RR: 1.24 (0.66 to 2.33)<br>Population studied: n = 525<br>**No statistically significant relationship** | ⊕⊕○○<br>Low |
| *see Table 1 for definitions of GRADE certainty<br>BMI: body mass index; CI: confidence interval; n: number of study participants; OR: odds ratio; RCT: randomised controlled trial; RR: risk ratio<br>a. Downgraded for imprecision due to wide confidence intervals around the effect estimate, including both being associated with an increased or decreased risk<br>b. Review authors report that publication bias was detected in the review<br>c. Wide confidence intervals around the point estimate of effect.<br>d. Review authors assessed studies as moderate and/or serious risk of bias |  |  |  |  |  |  |  |  |

##### 2.2.11.1 Type of weaning

Six studies in the reviews by Martinon-Torres et al. (2021) and Bergamini et al. (2022) reported mixed findings for the type of weaning and childhood obesity and/or overweight outcomes. One study reported that baby-led weaning was associated with decreased risk of childhood overweight, compared to traditional spoon-feeding (Brown 2015) (moderate-certainty evidence). A pooled analysis of two studies reported no statistically significant relationship between baby-led weaning or baby-led introduction to solids compared to other models of complementary feeding and childhood obesity or overweight (Dogan 2018, Taylor 2017) (low-certainty evidence). One pooled analysis of two studies reported a decreased risk of childhood obesity or overweight using responsive complementary feeding compared to other models of complementary feeding (Machuca 2016, Paul 2018) (low-certainty evidence). Another study by Savage (2016) also compared responsive complementary feeding to other models of complementary feeding, but reported uncertainty or no association with childhood obesity or overweight (low certainty-evidence).

##### 2.2.11.2 Timing of weaning

Three studies in the reviews by Moorcroft et al. (2011), Weng et al. (2012) and Verga et al. (2022) reported mixed findings on the timing of the introduction of solid foods and the association with childhood obesity or overweight. One study reported an association between early introduction of solid foods (before four months of age) and risk of childhood overweight (Hawkins 2009). Another study found no effect or uncertainty of early introduction of complementary feeding (at four to six months of age) on childhood obesity or overweight status, compared to complementary feeding at older ages (Jonsdottir 2014). One study reported no statistically significant relationship between infants who were exclusively breastfed and received complementary feeding between four and six months, and those who received complementary feeding at six months, and childhood obesity or overweight (Huh 2011). This study also found no statistically significant relationship between formula-fed infants who received complementary feeding at four to five months compared to those who received complementary feeding at more than six months, and childhood obesity or overweight (Huh 2011). The results of each of these studies were judged to be low-certainty evidence.

##### 2.2.11.3 Breastfeeding

Five studies in the review by Weng et al. (2012) reported mixed findings for the association between breastfeeding (compared to infants who were never breastfed) and childhood overweight. Three studies reported a decreased risk of the association between breastfeeding and childhood overweight (moderate- to low-certainty evidence) (Armstrong 2002, Grummer-Strawn 2004, Hawkins 2009); the fourth study reported no effect or uncertainty (very low-certainty evidence) (Weyermann 2006).

#### 2.2.12 Feeding in infancy (qualitative evidence)

A summary of the design and characteristics of the studies in the reviews can be found in Section 6.2, Table 20.

One systematic review was identified which aimed to evaluate evidence on factors associated with infant overfeeding among Hispanic mothers that may explain the high infant overweight rates often seen in this ethnic group (Cartagena et al. 2014). The main theme in this paper was breastfeeding and formula-feeding beliefs, attitudes, and practices in the USA. The paper included 35 studies related to infant feeding with Hispanic-only mothers or studies with multiethnic participants in which findings were categorised by racial and ethnic groups (Cartagena et al. 2014).

##### 2.2.12.1 Infant Feeding Practices

The primary theme was the influence of infant feeding practices on childhood obesity. Cartagena et al. (2014) also reported on breastfeeding and formula-feeding practices along with family and cultural influences on infant feeding practices. Sub-themes include family and cultural influences of maternal feeding beliefs and practices; and maternal and paternal perceptions of infant feeding practices, weight gain and exercise.

##### 2.2.12.2 Breastfeeding and Formula Feeding Practices

Cartagena et al (2014) found that Hispanic mothers in the USA are more likely to initiate breastfeeding but are less likely to practice exclusive breastfeeding compared to White or Black mothers, with immigrant mothers showing higher rates of breastfeeding initiation but a decrease in exclusive breastfeeding practices with increased years of USA residence. For example, Harley (2007), cited in Cartagena et al. (2014), reported that the median duration of exclusive breastfeeding was one to two months for women living in the USA for 10 or fewer years, and less than one week for women living in the United States for 11 years or their entire lives. Several papers in Cartagena et al. (2014) examined the influence of acculturation on breastfeeding practices, with less acculturated mothers showing higher breastfeeding intention and initiation. Cartagena et al. (2014) found that concerns about breast milk supply led many Hispanic mothers to introduce formula feeding, with a preference for a combination of breast and bottle feeding (CBFF), citing benefits such as providing the infant with breast milk and additional nutrients from formula.

Holmes (2011), cited in Cartagena et al. (2014), found ethnicity to be independently associated with CBFF, with a higher percentage of Hispanic infants taking breast milk and formula milk daily during the first week of their life when compared with White or Black infants (24.4% of Hispanic infants, compared with 17.9% of Black infants and 7.2% of White infants). However, several studies found CBFF often leads to shorter breastfeeding durations, and Hispanic infants receiving combination feeding or formula exclusively tend to have higher caloric intakes, potentially contributing to infants become overweight or obese. Holmes (2011), cited in Cartagena et al. (2014), found that infants exclusively breastfed for four months showed a decreased risk of becoming overweight or obese between age two and six years.

This section has highlighted the complex interplay between ethnicity, acculturation, breastfeeding practices, and infant health outcomes among Hispanic mothers in the United States. The research found that whilst Hispanic mothers demonstrate higher rates of breastfeeding initiation, there is a trend of decreased exclusive breastfeeding and an increasing trend in CBFF. While CBFF is associated with higher caloric intake and potential risks of infant obesity, exclusive breastfeeding for extended periods shows protective effects against these outcomes.

#### 2.2.13 Child’s diet (quantitative evidence)

Five reviews examined the effect of childhood diet as a factor for obesity or overweight in children less than five years old (Dror 2014, Ferre et al. 2021, Frantsve-Hawley et al. 2017, Kwansa et al. 2022, Lu et al. 2016). A summary of the design and characteristics of the studies in the reviews can be found in Section 6.2, Table 19. Comprehensive details of the results for this section are available in Table 10.

**Table 10:** Summary of reviews reporting child’s diet as a factor associated with childhood overweight or obesity: adapted from GRADE.

| No. of primary studies (systematic review reference) | Certainty assessment |  |  |  |  |  | Impact (95% CI) | Certainty |
| --- | --- | --- | --- | --- | --- | --- | --- | --- |
|  | Study design | Risk of bias | Inconsistency | Indirectness | Imprecision | Other considerations |  |  |
| Obesity outcomes |  |  |  |  |  |  |  |  |
| One to less-than-2 servings of sugar-containing beverages per day versus less-than-1 serving of sugar-containing beverage per day: Obesity outcomes (follow-up: range 36 months to 48 months; assessed with: BMI or BMI-related measure [primary outcome]) |  |  |  |  |  |  |  |  |
| 1 Frantsve-Hawley et al. (2017) | non-randomised studies | serious<sup>d</sup> | not serious | not serious | serious<sup>a</sup> | None | Welsh (2011): OR: 1.5 (0.9 to 2.4)<br>Population studied: n = 10,904<br>No statistically significant relationship | ⊕⊕○○<br>Low |
| Two to less-than-3 servings of sugar-containing beverages per day versus less-than-1 serving of sugar-containing beverage per day: Obesity outcomes (follow-up: range 36 months to 48 months; assessed with: BMI or BMI-related measure [primary outcome]) |  |  |  |  |  |  |  |  |
| 1 Frantsve-Hawley et al. (2017) | non-randomised studies | serious<sup>d</sup> | not serious | not serious | serious<sup>a</sup> | none | Welsh (2011): OR: 1.4 (0.8 to 2.3)<br>Population studied: n = 10,904<br>No statistically significant relationship | ⊕⊕○○<br>Low |
| More-than-3 servings of sugar-containing beverages per day versus less-than-1 serving of sugar-containing beverage per day: Obesity outcomes (follow-up: range 36 months to 48 months; assessed with: BMI or BMI-related measure [primary outcome]) |  |  |  |  |  |  |  |  |
| 1 Frantsve-Hawley et al. (2017) | non-randomised studies | serious<sup>d</sup> | not serious | not serious | serious<sup>a</sup> | none | Welsh (2011): OR: 1.3 (0.8 to 2.1)<br>Population studied: n = 10,904<br>No statistically significant relationship | ⊕⊕○○<br>Low |
| One to less-than-2 servings of fruit juice per day versus less-than-1 serving of fruit juice per day: Obesity outcomes (follow-up: range 36 months to 48 months; assessed with: BMI or BMI-related measure [primary outcome]) |  |  |  |  |  |  |  |  |
| 1 Frantsve-Hawley et al. (2017) | non-randomised studies | serious<sup>d</sup> | not serious | not serious | serious<sup>a</sup> | none | Welsh (2011): OR: 1.1 (0.8 to 1.5)<br>Population studied: n = 10,904<br>No statistically significant relationship | ⊕⊕○○<br>Low |
| Two to less-than-3 servings of fruit juice per day versus less-than-1 serving of fruit juice per day: Obesity outcomes (follow-up: range 36 months to 48 months; assessed with: BMI or BMI-related measure [primary outcome]) |  |  |  |  |  |  |  |  |

| No. of primary studies (systematic review reference) | Certainty assessment |  |  |  |  |  | Impact (95% CI) | Certainty* |
| --- | --- | --- | --- | --- | --- | --- | --- | --- |
|  | Study design | Risk of bias | Inconsistency | Indirectness | Imprecision | Other considerations |  |  |
| 1 Frantsve-Hawley et al. (2017) | non-randomised studies | serious<sup>d</sup> | not serious | not serious | serious<sup>a</sup> | none | Welsh (2011): OR: 1.0 (0.7 to 1.4)<br>Population studied: n = 10,904<br>**No statistically significant relationship** | ⊕⊕○○<br>Low |
| **More-than-3 servings of fruit juice per day versus less-than-1 serving of fruit juice per day: Obesity outcomes (follow-up: range 36 months to 48 months; assessed with: BMI or BMI-related measure [primary outcome])** |  |  |  |  |  |  |  |  |
| 1 Frantsve-Hawley et al. (2017) | non-randomised studies | serious<sup>d</sup> | not serious | not serious | serious<sup>a</sup> | none | Welsh (2011): OR:1.2 (0.8 to 1.7)<br>Population studied: n = 10,904<br>**No statistically significant relationship** | ⊕⊕○○<br>Low |
| **Overweight outcomes** |  |  |  |  |  |  |  |  |
| **Dairy intake: consumption of 1% milk: Overweight outcomes (follow-up: 24 months; assessed with: BMI or BMI-related measure [primary outcome])** |  |  |  |  |  |  |  |  |
| 1 Lu et al. (2016) | non-randomised studies | not serious | not serious | not serious | serious<sup>a</sup> | none | Huh (2010): OR: 0.95 (0.58 to 1.55)<br>Population studied: n = 645<br>**No statistically significant relationship** | ⊕⊕⊕○<br>Moderate |
| **Dairy intake: consumption of 2% milk: Overweight outcomes (follow-up: 24 months; assessed with: BMI or BMI-related measure [primary outcome])** |  |  |  |  |  |  |  |  |
| 1 Lu et al. (2016) | non-randomised studies | not serious | not serious | not serious | serious<sup>a</sup> | none | Huh (2010): OR: 0.91 (0.62 to 1.34)<br>Population studied: n = 645<br>**No statistically significant relationship** | ⊕⊕⊕○<br>Moderate |
| **Dairy intake: consumption of whole milk: Overweight outcomes (follow-up: 24 months; assessed with: BMI or BMI-related measure [primary outcome])** |  |  |  |  |  |  |  |  |
| 1 Lu et al. (2016) | non-randomised studies | not serious | not serious | not serious | serious<sup>a</sup> | none | Huh (2010): OR: 1.04 (0.74 to 1.44)<br>Population studied: n = 645<br>**No statistically significant relationship** | ⊕⊕⊕○<br>Moderate |
| **Regular consumption of sugar-containing beverages between meals: Overweight outcomes (follow-up: 54 months; assessed with: BMI or BMI-related measure [primary outcome])** |  |  |  |  |  |  |  |  |
| 1 Frantsve-Hawley et al. (2017) | non-randomised studies | not serious | not serious | not serious | serious<sup>c</sup> | none | Dubois (2007): OR: 2.36 (1.03 to 5.39)<br>Population studied: n = 1,499<br>**Associated with an increased risk** | ⊕⊕⊕○<br>Moderate |

| Obesity or overweight outcomes |  |  |  |  |  |  |  |  |
| --- | --- | --- | --- | --- | --- | --- | --- | --- |
| Consumption of sugar-sweetened beverages (> 1 bottle in 2 days): Obesity or overweight outcomes (follow-up: range 24 months to 60 months; assessed with: BMI or BMI-related measure [primary outcome]) |  |  |  |  |  |  |  |  |
| 1 Kwansa et al. (2022) | non-randomised studies | serious<sup>f</sup> | not serious | serious<sup>g</sup> | serious<sup>c</sup> | none | Mezie-Okoye (2015): OR: 18.98 (7.6 to 47.40)<br>Population studied: n = 220<br>**Associated with an increased risk** | ⊕○○○<br>Very low |
| Consumption of fried/fatty foods: Obesity or overweight outcomes (follow-up: range 24 months to 60 months; assessed with: BMI or BMI-related measure [primary outcome]) |  |  |  |  |  |  |  |  |
| 1 Kwansa et al. (2022) | non-randomised studies | serious<sup>f</sup> | not serious | serious<sup>g</sup> | serious<sup>c</sup> | none | Mezie-Okoye (2015): OR: 2.16 (1.01 to 4.61)<br>Population studied: n = 220<br>**Associated with an increased risk** | ⊕○○○<br>Very low |
| Consumption of fast food: Obesity or overweight outcomes (follow-up: range 36 months to 60 months; assessed with: BMI or BMI-related measure [primary outcome]) |  |  |  |  |  |  |  |  |
| Kwansa et al. (2022) | non-randomised studies | serious<sup>f</sup> | not serious | serious<sup>g</sup> | serious<sup>c</sup> | none | Wolde (2014): OR: 8.69 (1.11 to 13.50)<br>Population studied: n = 358<br>**Associated with an increased risk** | ⊕○○○<br>Very low |
| Consumption of sweet foods: Obesity or overweight outcomes (follow-up: range 36 months to 60 months; assessed with: BMI or BMI-related measure [primary outcome]) |  |  |  |  |  |  |  |  |
| 2 Kwansa et al. (2022) | non-randomised studies | serious<sup>f</sup> | not serious | serious<sup>g</sup> | serious<sup>c</sup> | none | Sorrie (2017): OR: 2.69 (1.21 to 5.98); Wolde (2014): OR: 6.36 (1.88 to 12.33)<br>Population studied: n = 858<br>**Associated with an increased risk** | ⊕○○○<br>Very low |
| Consumption of protein: Obesity and overweight outcomes (follow-up: 36 months; assessed with: BMI or BMI-related measure [primary outcome]) |  |  |  |  |  |  |  |  |
| 1 Ferre et al. (2021) | non-randomised studies | not serious | not serious | not serious | serious<sup>a</sup> | none | Pimpin (2016): OR: 1.10 (0.99 to 1.22)<br>Population studied: n = 2,435<br>**No statistically significant relationship** | ⊕⊕⊕○<br>Moderate |
| High dietary diversity: Obesity or overweight outcomes (follow-up: range 36 months to 60 months; assessed with: BMI or BMI-related measure [primary outcome]) |  |  |  |  |  |  |  |  |
| 2 Kwansa et al. (2022) | non-randomised studies | serious<sup>f</sup> | not serious | serious<sup>g</sup> | serious<sup>c</sup> | none | Sorrie (2017): OR: 3.73 (1.15 to 12.54); Wolde (2014): OR: 3.48 (1.50 to 8.10)<br>Population studied: n = 858<br>**Associated with an increased risk** | ⊕○○○<br>Very low |

| **Dairy intake: consumption of 1% milk compared to 2% milk: Overweight outcomes (follow-up: 48 months; assessed with: BMI or BMI-related measure [primary outcome])** |  |  |  |  |  |  |  |  |
| --- | --- | --- | --- | --- | --- | --- | --- | --- |
| 1 Dror (2014), Lu et al. (2016) | non-randomised studies | not serious | not serious | not serious | not serious | none | Scharf (2013): OR: 1.57 (1.03 to 2.42)<br>Population studied: n = 8,300<br>**Associated with an increased risk** | ⊕⊕⊕⊕<br>High |
| *see Table 1 for definitions of GRADE certainty<br>BMI: body mass index; CI: confidence interval; n: number of study participants; OR: odds ratio<br>a. Downgraded for imprecision due to wide confidence intervals around the effect estimate, including both being associated with an increased or decreased risk<br>c. Wide confidence intervals around the point estimate of effect.<br>d. Review authors assessed studies as moderate and/or serious risk of bias<br>f. Cross-sectional study design used.<br>g. Downgraded for indirectness because studies did not include countries/setting(s)/population(s) likely to be directly relevant to the UK, or countries were not reported in the studies |  |  |  |  |  |  |  |  |

##### 2.2.13.1 Dairy consumption

Two studies in the reviews by Lu et al. (2016) and Dror (2014) reported mixed findings for the association between consumption of milk and childhood obesity and overweight outcomes in children under five years. One study identified in the reviews by Lu et al. (2016) and Dror (2014) reported an association between consumption of 1% milk and risk of childhood obesity, compared to consumption of 2% milk (Scharf 2013) (high-certainty evidence). Another study included in the review by Dror (2014) reported no statistically significant relationship between consumption of 1% milk, 2% milk or whole milk and childhood overweight (Huh 2010) (moderate-certainty evidence).

##### 2.2.13.2 Sugar consumption

Five studies in the reviews by Kwansa et al. (2022) and Frantsve-Hawley et al. (2017) reported mixed findings for the association between sugar-containing beverages, fruit juice or sweet foods and childhood obesity and/or overweight. One study investigated several different levels of sugar-containing drink consumption (see Table 10 for details) and reported no statistically significant relationship between consuming at least one sugar-containing beverage or fruit-juice serving per day and childhood obesity (Welsh 2011) (low-certainty evidence). One study found an association between consumption of more than one bottle of sugar-sweetened beverages every two days and increased risk of childhood obesity or overweight (Mezie-Okoye 2015) (very low-certainty evidence). Another study found an association between regular consumption of sugar-containing beverages between meals and risk of childhood overweight (Dubois 2007) (moderate-certainty evidence). Two studies reported an association between consumption of sweet foods and childhood obesity or overweight outcomes (Sorrie 2017, Wolde 2014) (very low-certainty evidence).

##### 2.2.13.3 Fried/fatty food consumption

One study by Mezie-Okoye (2015), in the review by Kwansa et al. (2022), reported an association between consumption of fried/fatty foods and increased risk of childhood obesity or overweight. The results were judged to be very low-certainty evidence.

##### 2.2.13.4 Fast-food consumption

One study by Wolde (2014), in the review by Kwansa et al. (2022), reported an association between consumption of fast food and childhood obesity or overweight outcomes. The results were judged to be very low-certainty evidence.

##### 2.2.13.5 Protein consumption

One study by Pimpin (2016), in the study by Ferre et al. (2021), reported uncertainty or no association between consumption of protein and childhood obesity or overweight. The results were judged to be moderate-certainty evidence.

##### 2.2.13.6 High dietary diversity

Two studies reported an association between high dietary diversity and childhood obesity or overweight outcomes (Sorrie 2017, Wolde 2014) (very low-certainty evidence).

##### 2.2.13.7 Dietary inflammatory potential

No relevant reviews were identified that reported dietary inflammatory potential (the effect of diet on the levels of inflammatory factors in the body) in children’s healthy eating/physical activity interventions and its association with obesity or overweight in children less than five years old.

#### 2.2.14 Child’s diet (qualitative evidence)

A summary of the design and characteristics of the studies in the reviews can be found in Section 6.2, Table 20.

One systematic review was identified which aimed to evaluate evidence on factors associated with infant overfeeding among Hispanic mothers that may explain the high infant overweight rates often seen in this ethnic group (Cartagena et al. 2014).

Reifsnider (2008), cited in Cartagena et al. (2014), found that overweight infants tended to have higher daily intakes of specific foods, such as Mexican rice and sugary beverages.

Davis (2012), referenced in Cartagena et al. (2014), examined the independent and additive effects of breastfeeding and sugar-sweetened beverage intake in infancy on the prevalence of childhood overweight and obesity. Overall, children who were breastfed for at least 12 months and did not consume sugar-sweetened beverages had a lower chance of becoming obese (60% decrease in the odds of obesity [95% CI: 0.19 to 0.80]) compared to children who were not breastfed and reported a high sugar-sweetened beverage intake.

#### 2.2.15 Child’s physical activity

No relevant reviews were identified that reported child’s physical activity and any association with obesity and/or overweight in children less than five years old.

#### 2.2.16 Sleep duration (quantitative evidence)

One review by Miller et al. (2018a) examined the effect of sleep duration as a factor for obesity or overweight in children less than five years old. A summary of the design and characteristics of the studies in the reviews can be found in Section 6.2, Table 19.

Comprehensive details of the results for this section are available in Table 11.

**Table 11:** Summary of reviews reporting sleep duration as a factor associated with childhood overweight or obesity: adapted from GRADE.

| No. of primary studies (systematic review reference) | Certainty assessment |  |  |  |  |  | Impact (95% CI) | Certainty |
| --- | --- | --- | --- | --- | --- | --- | --- | --- |
|  | Study design | Risk of bias | Inconsistency | Indirectness | Imprecision | Other considerations |  |  |
| Obesity or overweight outcomes |  |  |  |  |  |  |  |  |
| Short sleep duration: Obesity or overweight outcomes (follow-up: range 0 months to 36 months; assessed with: BMI or BMI-related measure [primary outcome]) |  |  |  |  |  |  |  |  |
| 7 Miller et al. (2018a) | non-randomised studies | not serious | not serious | not serious | not serious | publication bias strongly suspected<sup>b</sup> | Pooled analysis of Bell (2010), Bolijn (2016), Diethelm (2011), Halal (2016), Reilly (2005), Taveras (2008), Touchette (2008) (total n = 14,738)<br>RR: 1.40 (1.19 to 1.65)<br>**Associated with an increased risk** | ⊕⊕⊕○<br>Moderate |
| *see Table 1 for definitions of GRADE certainty<br>BMI: body mass index; CI: confidence interval; n: number of study participants; RR: risk ratio |  |  |  |  |  |  |  |  |
| b. Review authors report that publication bias was detected in the review |  |  |  |  |  |  |  |  |

Pooled analysis of seven studies in the review by Miller et al. (2018a) reported an association between short sleep duration and increased risk of childhood obesity or overweight (moderate-certainty evidence).

#### 2.2.17 Caregiver behaviour (quantitative evidence)

One review by Kwansa et al. (2022) examined the effect of caregiver behaviour as a factor for obesity or overweight in children less than five years old. A summary of the design and characteristics of the studies in the reviews can be found in Section 6.2, Table 19.

Comprehensive details of the results for this section are available in Table 12.

**Table 12:** Summary of reviews reporting caregiver behaviour as a factor associated with childhood overweight or obesity: adapted from GRADE.

| No. of primary studies (systematic review reference) | Certainty assessment |  |  |  |  |  | Impact (95% CI) | Certainty* |
| --- | --- | --- | --- | --- | --- | --- | --- | --- |
|  | Study design | Risk of bias | Inconsistency | Indirectness | Imprecision | Other considerations |  |  |
| Overweight outcomes |  |  |  |  |  |  |  |  |
| Maternal under-evaluation of child's weight: Overweight outcomes (follow-up: range 24 months to 60 months; assessed with: BMI or BMI-related measure [primary outcome]) |  |  |  |  |  |  |  |  |
| 1 Kwansa et al. (2022) | non-randomised studies | very serious<sup>d,f</sup> | not serious | serious<sup>g</sup> | serious<sup>c</sup> | none | Said-Mohamed (2009): OR: 6.52 (2.34 to 18.09)<br>Population studied: n = 165<br>**Associated with an increased risk** | ⊕○○○<br>Very low |
| *see Table 1 for definitions of GRADE certainty<br>BMI: body mass index; CI: confidence interval; n: number of study participants; OR: odds ratio<br>c. Large reported effect, but also very wide confidence intervals around the point estimate of effect.<br>d. Review authors assessed studies as moderate and/or serious risk of bias<br>f. Cross-sectional study design used.<br>g. Downgraded for indirectness because studies did not include countries/setting(s)/population(s) likely to be directly relevant to the UK, or countries were not reported in the studies |  |  |  |  |  |  |  |  |

##### 2.2.17.1 Maternal under-evaluation of child’s weight

One study by Said-Mohamed (2009), in the review by Kwansa et al. (2022), reported an association between maternal under-evaluation of a child’s weight and overweight outcomes (very low-certainty evidence).

##### 2.2.17.2 Neglect or abuse

No relevant reviews were identified that reported neglect or abuse and its association with obesity or overweight in children less than five years old.

##### 2.2.17.3 Caregiver involvement in children’s healthy eating/physical activity interventions

No relevant reviews were identified that reported caregiver involvement in children’s healthy eating/physical activity interventions and its association with obesity or overweight in children less than five years old.

##### 2.2.17.4 Maternal physical activity

No relevant reviews were identified that reported maternal physical activity and its association with obesity or overweight in children less than five years old.

#### 2.2.18 Caregiver behaviour (qualitative evidence)

A summary of the design and characteristics of the studies in the reviews can be found in Section 6.2, Table 20.

A qualitative review by Cartagena et al. (2014) aimed to evaluate evidence on factors associated with infant overfeeding among Hispanic mothers that may explain the high infant overweight rates often seen in this ethnic group. Fraser et al. (2011) conducted a review of the literature on paternal influences on children’s weight gain and obesity. Emerging themes included paternal perceptions of infant feeding practices and weight gain. The review included 10 studies relating to paternal influences on children’s weight gain. Countries within the individual studies included UK, USA, France, and Australia.

##### 2.2.18.1 Family and Cultural Influences on Maternal Feeding Beliefs and Practices

Higgins (2000) and Worobey (2005), cited in Cartagena et al. (2014), found that Hispanic mothers often hold rooted cultural beliefs about what constitutes a healthy weight for infants. They perceive increased body weight as a sign of good health and may have difficulty recognising obesity in their children. Furthermore, there is a preference for heavier infants, with the belief that “a chubby baby is a happy and healthy baby.” Within the Hispanic community, parents and grandparents perceive feeding as a way of nurturing and caring for their child. In Hispanic households, meeting children’s food cravings is seen as a crucial aspect of effective parenting, although it often involves permitting unhealthy food choices and overfeeding. Additionally, family members, for example grandmothers, typically influence Hispanic mothers’ beliefs and behaviours regarding feeding practices. Hispanic mothers frequently opt not to adhere to clinicians’ recommendations regarding infant feeding, particularly if such advice contradicts the guidance and/or beliefs of family members and friends. These mothers often receive encouragement from family members and fathers to introduce formula supplementation if infants are fussy or not ‘gordito’ (chubby).

Maternal perceptions of infant weight status significantly influence daily feeding decisions. The perceived weight status of a mother’s child guides many of these decisions, with heavier infants often receiving more food. Additionally, maternal perception of feeding satiety plays a role, with mothers interpreting crying and fussiness as signs of hunger, leading to increased feeding and an increased risk of the child becoming overweight. Worobey (2005), cited in Cartagena et al. (2014), found that the number of feeds per day at six-months and the maternal lack of sensitivity to the infant’s satiety cues, were significantly related to the infant’s weight gain from age six to 12 months. Hispanic mothers are also more likely to introduce solid foods (weaning) much earlier than average. Cartagena et al. (2014) found Hispanic mothers often introduced solid foods and cereal before the infants were age 4 months.

In summary, this section highlights the profound influence of cultural beliefs and family dynamics on infant feeding practices among Hispanic mothers in the USA. The focus on this ethnic group means the generalisability of the findings to Wales is uncertain. Rooted cultural perceptions, such as associating increased body weight with good health and happiness, shape maternal attitudes towards infant weight status. This perception often leads to overfeeding and early introduction of solid foods, contributing to an increased risk of childhood overweight and obesity. Additionally, familial influences, particularly from grandparents, play a significant role in shaping feeding behaviours, often conflicting with clinical recommendations. Maternal perceptions of infant satiety is also important, with crying and fussiness often interpreted as signs of hunger, leading to excessive feeding.

##### 2.2.18.2 Paternal Perceptions on Infant Feeding Practices, Weight Gain and Exercise

Blissett (2006), a UK study, cited in Fraser et al. (2011), compared maternal and paternal eating-related attitudes and feeding practices and found that mothers and fathers did not differ in their use of restrictive or pressurising practices. However, fathers with greater body dissatisfaction tend to monitor their sons’ food intake more closely, especially if the sons have a higher BMI. Haycraft (2008), a UK study, (2008) in Fraser et al (2011), found that fathers of boys with a high BMI saw their sons as more overweight and were more concerned about their weight compared to fathers of boys with an average BMI. Yet it was fathers of boys with an average BMI that used more controlling child-feeding practices, such as monitoring of food intake and pressure to eat. Additionally, Musher-Eizenman (2009), citied in Fraser et al. (2011), found that there are differences in feeding practices between fathers from different cultural backgrounds, with USA fathers tending to allow more child control over food intake and use food for non-nutritive purposes, while French fathers engage more in monitoring and restriction of food intake for weight control.

Snethen (2008), in Fraser et al. (2011), investigated fathers’ perceptions of dietary and exercise habits and their potential impact on childhood obesity. The study found that most fathers misclassified themselves as having a normal weight, despite 68% being overweight or obese. Similarly, they underestimated their children’s weight, with 20% classified as normal or underweight when they were overweight or obese. Overweight fathers were more prone to underestimating their children’s weight compared to normal weight fathers. Fraser et al. (2011) concluded that if fathers do not perceive overweight or obesity in their children, it is less likely that they will implement or support effective weight management strategies. Both fathers and children in the study consumed less than the recommended daily servings of fruits and vegetables. Additionally, 97% of children had unrestricted access to snacks and sweets at home and often indulged without parental supervision. Fathers reported that overweight children were more likely to exhibit behaviours such as consuming fast food, eating quickly, eating out of boredom, and less likely to have family dinners together. It is unclear whether these behaviours stem from children mimicking their fathers’ habits, but the study found a correlation between fathers’ weight and the amount of time children spent in sedentary activities, like watching TV or using the computer.

These studies emphasise the important role of both mothers and fathers in shaping eating-related attitudes and feeding practices within the family dynamic. While mothers and fathers generally do not differ significantly in their use of restrictive or pressurising feeding practices, paternal attitudes and behaviours can vary based on factors such as body dissatisfaction and the child’s BMI. Fathers, particularly those with greater body dissatisfaction, may exhibit closer monitoring of their sons’ food intake, especially if the sons have a higher BMI. Paternal perceptions of their child’s weight status may not always align with their feeding practices, as fathers of boys with higher BMIs may perceive their sons as more overweight but may not necessarily use more controlling feeding practices. Cultural background also plays a role, with differences observed between fathers from the USA and France in terms of allowing child control over food intake and using food for non-nutritive purposes versus monitoring and restricting food intake for weight control. The study by Snethen (2008), within Fraser et al. (2011), highlights significant misperceptions among fathers regarding both their own and their children’s weight status, with implications for effective weight management strategies

#### 2.2.19 Parenting styles (qualitative evidence)

No relevant quantitative evidence for this factor was identified. Fraser et al. (2011) conducted a review of the literature on paternal influences on children’s weight gain and obesity. Emerging themes were parenting styles and parental knowledge. The paper included 10 studies relating to paternal influences on children’s weight gain. Countries within the individual studies included UK, USA, France, and Australia. A summary of the study design and characteristics are available in Section 6.2, Table 20.

##### 2.2.19.1 Paternal parenting styles

Several studies in Fraser et al. (2011) explored the impact of paternal parenting styles on child feeding practices and weight outcomes. The following themes were identified: permissive parenting styles, authoritative parenting style, low-paternal parenting control and a change in paternal parenting style. Results relating to parenting style varied across studies in Fraser et al. (2011). Whilst one study (Brann 2005) found paternal parenting styles (for example, authoritarian, authoritative, or permissive) do not directly relate to child BMI or weight status, Wake (2007) cited in Fraser et al. (2011) found that fathers’ parenting styles were linked to an increased risk of children becoming overweight and/or obese. Additionally, Stein (2005), cited in Fraser et al. (2011), explored parenting style as a predictor of weight loss maintenance in a paediatric obesity treatment program. They found that changes in paternal-parenting style related to child weight outcomes and treatment success. Greater paternal acceptance correlated with reduced child overweight percentage. These findings suggest that supportive and firm paternal parenting may mitigate child overweight and obesity.

##### 2.2.19.2 Paternal and child eating and physical activity

Johannsen (2006) in Fraser et al. (2011) found no significant relationship between parental eating behaviours and children’s BMI, although there were associations between parental behaviour and body fat. The study found that fathers who displayed controlling behaviours had daughters with a higher percentage of body fat, and these same fathers also reported more concern for their children’s future health. However, the relationship between the controlling behaviour during feeding such as restriction and children’s weight was still unclear. For example, whether a parent’s feeding practice or parenting style affects the child’s weight status, or if the child’s weight status affects parenting style or how the parent feeds.

##### 2.2.19.3 Paternal Knowledge

Horodynski (2005) cited in Fraser et al. (2011) aimed at understanding the mealtime behaviours of African American fathers to gain insights into feeding practices that contribute to childhood obesity in this demographic. Five key themes emerged from the study: mealtime rituals and routines, division of responsibility, family structure, knowledge about healthy eating behaviours, and tension during meals.

The findings reported that fathers possessed a certain level of knowledge regarding mealtime interactions and nutrition for their toddlers, as well as a basic understanding of child growth and development. However, some knowledge gaps were evident, leading to actions such as yielding to their child’s preferences and demands rather than addressing them, and spoon-feeding instead of promoting self-feeding. Previous research cited in Fraser et al. (2011) has shown that these behaviours can lead to negative weight outcomes for children.

Findings suggest that similar perspectives and practices may be prevalent among fathers from various socioeconomic and cultural backgrounds. They emphasise the importance of employing a range of strategies to foster healthy eating habits and overall well-being in children, including setting limits, establishing consistent routines, anticipating the child’s needs, interpreting nonverbal cues, maintaining physical and emotional closeness, and modelling desirable behaviours. These factors align with findings from cross-sectional research on child weight outcomes. Thus, mealtime behaviours among African American fathers is important in shaping childhood obesity trends. The identified themes highlight both the knowledge and gaps in fathers’ feeding practices.

#### 2.2.20 Socioeconomic status (quantitative evidence)

Two reviews by Kwansa et al. (2022) and Lou et al. (2022) examined the effect of socioeconomic status as a factor for obesity or overweight in children less than five years old. A summary of the design and characteristics of the studies in the reviews can be found Section 6.2, Table 19. Comprehensive details of the results for this section are available in Table 13.

**Table 13:**
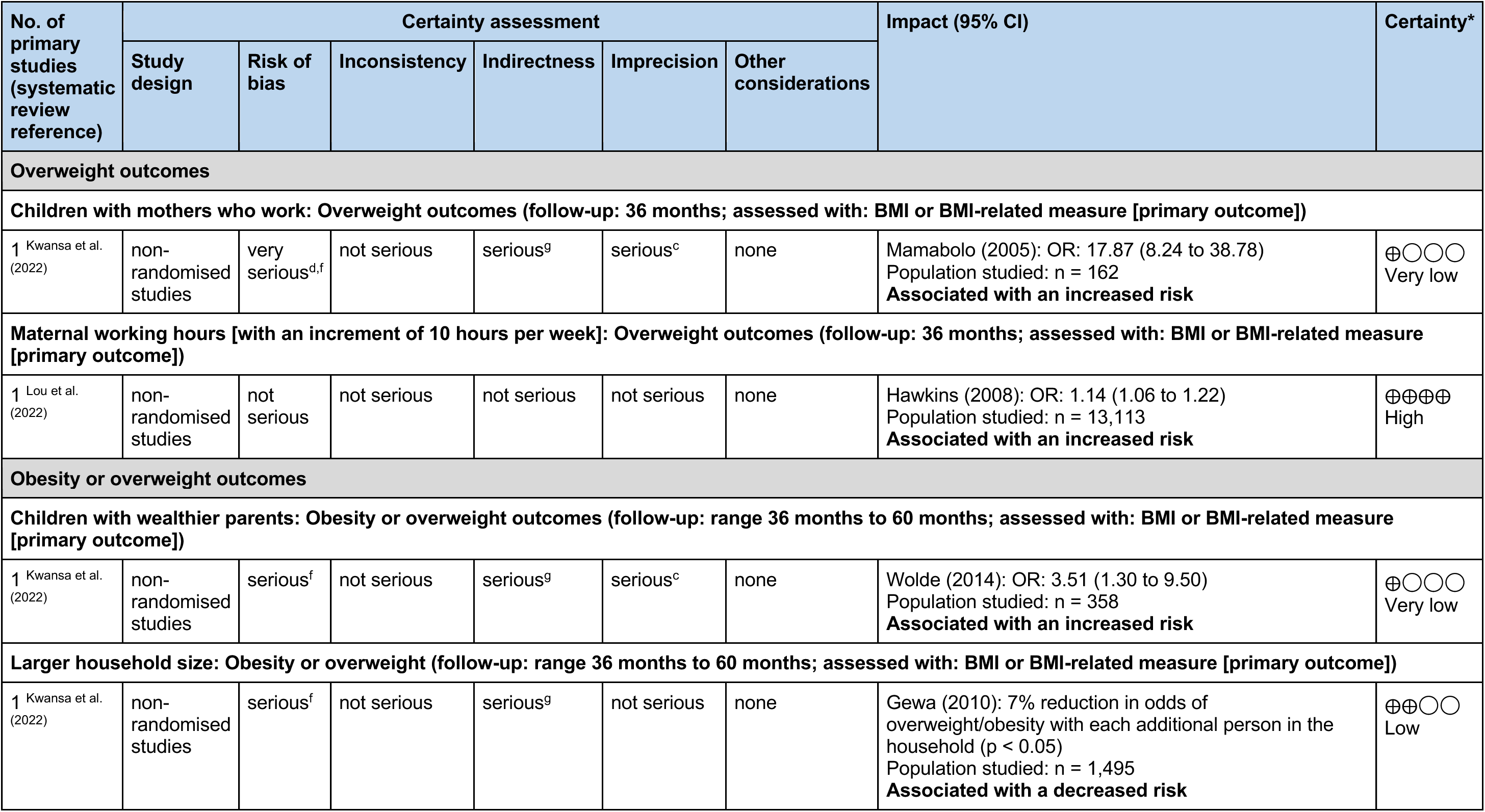
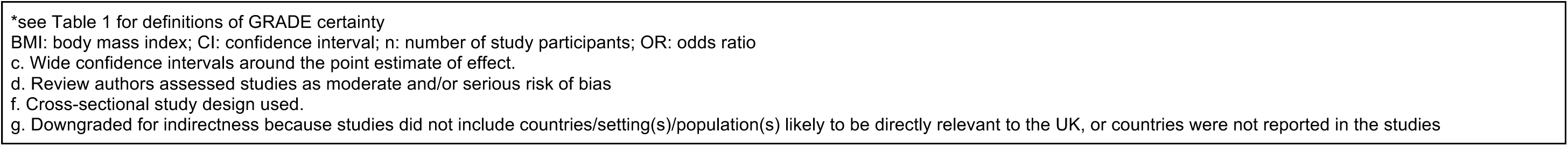
Summary of reviews reporting socioeconomic status as a factor associated with childhood overweight or obesity: adapted from GRADE.

| No. of primary studies (systematic review reference) | Certainty assessment |  |  |  |  |  | Impact (95% CI) | Certainty |
| --- | --- | --- | --- | --- | --- | --- | --- | --- |
|  | Study design | Risk of bias | Inconsistency | Indirectness | Imprecision | Other considerations |  |  |
| Overweight outcomes |  |  |  |  |  |  |  |  |
| Children with mothers who work: Overweight outcomes (follow-up: 36 months; assessed with: BMI or BMI-related measure [primary outcome]) |  |  |  |  |  |  |  |  |
| 1 Kwansa et al. (2022) | non-randomised studies | very serious<sup>d,f</sup> | not serious | serious<sup>g</sup> | serious<sup>c</sup> | none | Mamabolo (2005): OR: 17.87 (8.24 to 38.78)<br>Population studied: n = 162<br>Associated with an increased risk | ⊕○○○<br>Very low |
| Maternal working hours [with an increment of 10 hours per week]: Overweight outcomes (follow-up: 36 months; assessed with: BMI or BMI-related measure [primary outcome]) |  |  |  |  |  |  |  |  |
| 1 Lou et al. (2022) | non-randomised studies | not serious | not serious | not serious | not serious | none | Hawkins (2008): OR: 1.14 (1.06 to 1.22)<br>Population studied: n = 13,113<br>Associated with an increased risk | ⊕⊕⊕⊕<br>High |
| Obesity or overweight outcomes |  |  |  |  |  |  |  |  |
| Children with wealthier parents: Obesity or overweight outcomes (follow-up: range 36 months to 60 months; assessed with: BMI or BMI-related measure [primary outcome]) |  |  |  |  |  |  |  |  |
| 1 Kwansa et al. (2022) | non-randomised studies | serious<sup>f</sup> | not serious | serious<sup>g</sup> | serious<sup>c</sup> | none | Wolde (2014): OR: 3.51 (1.30 to 9.50)<br>Population studied: n = 358<br>Associated with an increased risk | ⊕○○○<br>Very low |
| Larger household size: Obesity or overweight (follow-up: range 36 months to 60 months; assessed with: BMI or BMI-related measure [primary outcome]) |  |  |  |  |  |  |  |  |
| 1 Kwansa et al. (2022) | non-randomised studies | serious<sup>f</sup> | not serious | serious<sup>g</sup> | not serious | none | Gewa (2010): 7% reduction in odds of overweight/obesity with each additional person in the household (p < 0.05)<br>Population studied: n = 1,495<br>Associated with a decreased risk | ⊕⊕○○<br>Low |

\*see Table 1 for definitions of GRADE certainty BMI: body mass index; CI: confidence interval; n: number of study participants; OR: odds ratio c. Wide confidence intervals around the point estimate of effect. d. Review authors assessed studies as moderate and/or serious risk of bias f. Cross-sectional study design used. g. Downgraded for indirectness because studies did not include countries/setting(s)/population(s) likely to be directly relevant to the UK, or countries were not reported in the studies

##### 2.2.20.1 Mothers who work

One study from a review by Kwansa et al. (2022) reported an association between children with mothers who work and increased risk of childhood overweight (Mamabolo 2005) (very low-certainty evidence). Another study from a review by Lou et al. (2022) found an association between increasing maternal working hours (measured in increments of 10 hours per week) and increased risk of childhood obesity (Hawkins 2008) (high-certainty evidence).

##### 2.2.20.2 Wealthier parents

One study by Wolde (2014) from a review by Kwansa et al. (2022) reported an increased risk of childhood obesity or overweight in children with wealthier parents (very-low certainty evidence).

##### 2.2.20.3 Larger household size

One study by Gewa (2010) from a review by Kwansa et al. (2022) reported an association between larger household sizes and decreased risk of childhood obesity or overweight (low-certainty evidence).

#### 2.2.21 Education (quantitative evidence)

One review by Kwansa et al. (2022) examined the effect of the education status of a caregiver as a factor for obesity or overweight in children less than five years old. A summary of the design and characteristics of the studies in the reviews can be found in Section 6.2, Table 19. Comprehensive details of the results for this section are available in Table 14.

**Table 14:**
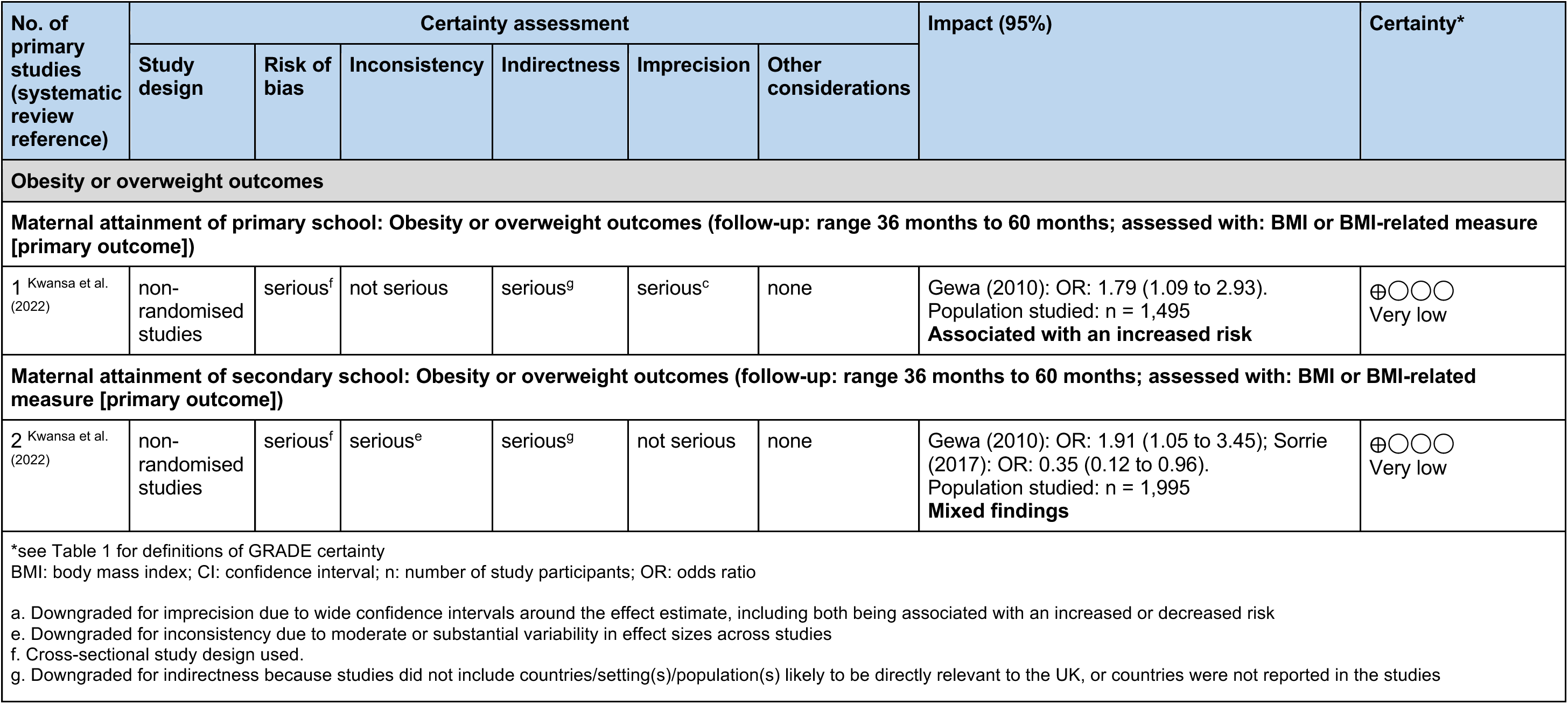
Summary of reviews reporting education as a factor associated with childhood overweight or obesity: adapted from GRADE.

##### 2.2.21.1 Maternal attainment of primary school

A study by Gewa (2010) in the review by Kwansa et al. (2022) reported an association between maternal attainment of primary school and childhood obesity or overweight outcomes (very low-certainty evidence).

##### 2.2.21.2 Maternal attainment of secondary school

Two studies by Gewa (2010) and Sorrie (2017) in the review by Kwansa et al. (2022) reported mixed findings between maternal attainment of secondary school and childhood obesity or overweight (very low-certainty evidence).

##### 2.2.21.3 COVID-19 related school closures

No relevant reviews were identified that reported COVID-19 related school closures and any association with obesity or overweight in children less than five years old.

#### 2.2.22 Food insecurity

No relevant reviews were identified that reported food insecurity and any association with obesity or overweight in children less than five years old.

#### 2.2.23 Food prices

No relevant reviews were identified that reported food prices and any association with obesity or overweight in children less than five years old.

#### 2.2.24 Built environment

No relevant reviews were identified that reported built environment-related factors (for example, the distribution of office, residential, retail, entertainment, sporting infrastructure and education within a given area) and any association with obesity or overweight in children less than five years old.

#### 2.2.25 Environmental exposures (quantitative evidence)

Seven reviews examined the effect of environmental exposures as a factor for obesity or overweight in children less than five years old (Baron et al. 2020, Gutierrez-Torres et al. 2018, Miller et al. 2018b, Riedel et al. 2014, Solans et al. 2022, Srivastava et al. 2020, Weng et al. 2012). A summary of the design and characteristics of the studies in the reviews can be found in Section 6.2, Table 19. Comprehensive details of the results for this section are available in Table 15.

**Table 15:** Summary of reviews reporting environmental exposures as a factor associated with childhood overweight or obesity: adapted from GRADE.

| No. of primary studies (systematic review reference) | Certainty assessment |  |  |  |  |  | Impact (95% CI) | Certainty* |
| --- | --- | --- | --- | --- | --- | --- | --- | --- |
|  | Study design | Risk of bias | Inconsistency | Indirectness | Imprecision | Other considerations |  |  |
| Obesity outcomes |  |  |  |  |  |  |  |  |
| Prenatal antibiotic exposure (overall - 1st, 2nd and 3rd trimester): Obesity outcomes (follow-up: 48 months; assessed with: BMI or BMI-related measure [primary outcome]) |  |  |  |  |  |  |  |  |
| 1 Baron et al. (2020), Solans et al. (2022) | non-randomised studies | not serious | not serious | not serious | serious<sup>a</sup> | publication bias strongly suspected<sup>b</sup> | Wang (2018): RR: 0.98 (0.90 to 1.06)<br>Population studied: n = 33,228<br>**No statistically significant relationship** | ⊕⊕○○<br>Low |
| Prenatal exposure to hexachlorobenzene (HCB): Obesity outcomes (follow-up: 48 months; assessed with: BMI or BMI-related measure [primary outcome]) |  |  |  |  |  |  |  |  |
| 1 Gutierrez-Torres et al. (2018) | non-randomised studies | not serious | not serious | not serious | serious<sup>c</sup> | none | Vafeiadi (2016): RR: 8.14 (1.85 to 35.81)<br>Population studied: n = 689<br>**Associated with an increased risk** | ⊕⊕⊕○<br>Moderate |
| Prenatal exposure to dichlorodiphenyl dichloroethene (DDE): Obesity outcomes (follow-up: 48 months; assessed with: BMI or BMI-related measure [primary outcome]) |  |  |  |  |  |  |  |  |
| 1 Gutierrez-Torres et al. (2018) | non-randomised studies | not serious | not serious | not serious | serious<sup>c</sup> | none | Vafeiadi (2016): RR: 3.80 (1.19 to 12.14)<br>Population studied: n = 689<br>**Associated with an increased risk** | ⊕⊕⊕○<br>Moderate |
| Postnatal (first 2 years of life) antibiotic exposure: Obesity outcomes (follow-up: range 24 months to 59 months; assessed with: BMI or BMI-related measure [primary outcome]) |  |  |  |  |  |  |  |  |
| 4 Srivastava et al. (2020) | non-randomised studies | serious<sup>d</sup> | serious<sup>e</sup> | not serious | serious<sup>a</sup> | none | Pooled analysis of Bailey (2014), Scott (2016), Trasande (2013), Ville (2017) (n = 116,021):<br>OR: 1.14 (0.97 to 1.35).<br>**No statistically significant relationship** | ⊕○○○<br>Very low |

| Overweight outcomes |  |  |  |  |  |  |  |  |
| --- | --- | --- | --- | --- | --- | --- | --- | --- |
| Prenatal (1st, 2nd and 3rd trimester) or postnatal (first 2 years of life) antibiotic exposure: Overweight outcomes (follow-up: range 12 months to 38 months; assessed with: Multiple primary outcome measures) |  |  |  |  |  |  |  |  |
| 4 Srivastava et al. (2020) | non-randomised studies | serious<sup>d</sup> | not serious | not serious | not serious | none | Pooled analysis of Bridgman (2018), Cassidy-Bushrow (2018), Saari (2015), Trasande (2013) (n = 16,411):<br>OR 1.22 (1.05 to 1.42).<br>**Associated with an increased risk** | ⊕⊕⊕○<br>Moderate |
| Prenatal (1st, 2nd and 3rd trimester) antibiotic exposure: Overweight outcomes (follow-up: 48 months; assessed with: BMI or BMI-related measure [primary outcome]) |  |  |  |  |  |  |  |  |
| 1 Baron et al. (2020) | non-randomised studies | not serious | not serious | not serious | serious<sup>a</sup> | none | Wang (2018): OR: 1.01 (0.97 to 1.06)<br>Population studied: n = 33,228<br>**No statistically significant relationship** | ⊕⊕⊕○<br>Moderate |
| Prenatal Bisphenol A (BPA) exposure: Overweight outcomes (follow-up: range 24 months to 60 months; assessed with: BMI or BMI-related measure [primary outcome]) |  |  |  |  |  |  |  |  |
| 2 Gutierrez-Torres et al. (2018) | non-randomised studies | not serious | not serious | not serious | serious<sup>a</sup> | none | Braun (2014): OR: 0.65 (0.19 to 2.18);<br>Valvi (2015): RR: 1.38 (0.72 to 2.67)<br>Population studied: n = 641<br>**No statistically significant relationship** | ⊕⊕⊕○<br>Moderate |
| Postnatal (first 2 years of life) antibiotic exposure: Overweight outcomes (follow-up: 24 months; assessed with: BMI or BMI-related measure [primary outcome]) |  |  |  |  |  |  |  |  |
| 1 Srivastava et al. (2020) | non-randomised studies | not serious | not serious | not serious | serious<sup>a</sup> | none | Saari (2015): OR: 1.25 (0.96 to 1.61)<br>Total population studied: (n = 14,764)<br>**No statistically significant relationship** | ⊕⊕⊕○<br>Moderate |
| Maternal smoking during pregnancy versus no maternal smoking during pregnancy: Overweight outcomes (follow-up: 36 months; assessed with: BMI or BMI-related measure [primary outcome]) |  |  |  |  |  |  |  |  |
| 1 Weng et al. (2012) | non-randomised studies | serious<sup>d</sup> | not serious | not serious | not serious | publication bias strongly suspected<sup>b</sup> | Hawkins (2009): OR: 1.34 (1.05 to 1.70)<br>Population studied: n = 13,188<br>**Associated with an increased risk** | ⊕⊕○○<br>Low |

| **Maternal smoking during pregnancy versus no maternal smoking during pregnancy: Overweight outcomes (follow-up: 48 months; assessed with: BMI or BMI-related measure [primary outcome])** |  |  |  |  |  |  |  |  |
| --- | --- | --- | --- | --- | --- | --- | --- | --- |
| 1 Riedel et al. (2014) | non-randomised studies | not serious | not serious | not serious | serious<sup>a</sup> | none | Heppe (2013): OR: 1.33 (0.96 to 1.84)<br>Population studied: n = 3,610<br>**No statistically significant relationship** | ⊕⊕⊕○<br>Moderate |
| 1 Weng et al. (2012) | non-randomised studies | serious<sup>d</sup> | not serious | not serious | serious<sup>c</sup> | publication bias strongly suspected<sup>b</sup> | Fasting (2009): OR: 2.83 (1.13 to 7.10)<br>Population studied: n = 711<br>**Associated with an increased risk** | ⊕○○○<br>Very low |
| **Maternal smoking during pregnancy versus no maternal smoking during pregnancy: Overweight outcomes (follow-up: 54 months; assessed with: BMI or BMI-related measure [primary outcome])** |  |  |  |  |  |  |  |  |
| 1 Weng et al. (2012) | non-randomised studies | serious<sup>d</sup> | not serious | not serious | not serious | publication bias strongly suspected<sup>b</sup> | Dubois (2006): OR: 1.80 (1.20 to 2.80)<br>Population studied: n = 1,550<br>**Associated with an increased risk** | ⊕⊕○○<br>Low |
| **Paternal smoking during pregnancy versus no paternal smoking during pregnancy: Overweight outcomes (follow-up: 48 months; assessed with: BMI or BMI-related measure [primary outcome])** |  |  |  |  |  |  |  |  |
| 1 Riedel et al. (2014) | non-randomised studies | not serious | not serious | not serious | serious<sup>a</sup> | none | Heppe (2013): OR: 1.03 (0.77 to 1.38)<br>Population studied: n = 3,610<br>**No statistically significant relationship** | ⊕⊕⊕○<br>Moderate |
| **Obesity or overweight outcomes** |  |  |  |  |  |  |  |  |
| **Prenatal (1st, 2nd and 3rd trimester) antibiotic exposure: Obesity or overweight outcomes (follow-up: range 24 months to 48 months; assessed with: BMI or BMI-related measure [primary outcome])** |  |  |  |  |  |  |  |  |
| 2 Solans et al. (2022) | non-randomised studies | not serious | not serious | not serious | serious<sup>a</sup> | none | Cassidy-Bushrow (2018): RR: 1.04 (0.62 to 1.74); Wang (2018): RR: 1.01 (0.97 to 1.06)<br>Population studied: n = 33,755<br>**No statistically significant relationship** | ⊕⊕⊕○<br>Moderate |

| Postnatal (first 2 years of life) antibiotic exposure: Obesity or overweight outcomes (follow-up: range 24 months to 60 months; assessed with: BMI or BMI-related measure [primary outcome]) |  |  |  |  |  |  |  |  |
| --- | --- | --- | --- | --- | --- | --- | --- | --- |
| 4 Miller et al. (2018b) | non-randomised studies | not serious | serious<sup>e</sup> | not serious | not serious | publication bias strongly suspected<sup>b</sup> | Pooled analysis of Bailey (2014), Poulsen (2017), Saari (2015), Scott (2016) (n = 107,049):<br>OR: 1.06 (1.01 to 1.12).<br>**Associated with an increased risk** | ⊕⊕○○<br>Low |
| *see Table 1 for definitions of GRADE certainty<br>BMI: body mass index; CI: confidence interval; n: number of study participants; OR: odds ratio; RR: risk ratio<br>a. Downgraded for imprecision due to wide confidence intervals around the effect estimate, including both being associated with an increased or decreased risk<br>b. Review authors report that publication bias was detected in the review<br>c. Wide confidence intervals around the point estimate of effect.<br>d. Review authors assessed studies as moderate and/or serious risk of bias<br>e. Downgraded for inconsistency due to moderate or substantial variability in effect sizes across studies |  |  |  |  |  |  |  |  |

##### 2.2.25.1 Prenatal antibiotic exposure

Two studies by Wang (2018) and Cassidy-Bushrow (2018), from two reviews by Baron et al. (2020) and Solans et al. (2022), reported no statistically significant relationship between prenatal (1^st^, 2^nd^ and 3^rd^ trimester) antibiotic exposure and childhood obesity and/or overweight outcomes (low- to moderate-certainty evidence).

##### 2.2.25.2 Prenatal chemical exposure

Three studies in one review by Gutierrez-Torres et al. (2018) reported mixed findings for the association between prenatal chemical exposure and childhood obesity and overweight outcomes. A study by Vafeiadi (2016) reported an association between prenatal exposure to hexachlorobenzene (HCB) and increased risk of childhood obesity, and prenatal exposure to dichlorodiphenyl dichloroethene (DDE) and childhood obesity (moderate-certainty evidence). Two studies found no statistically significant relationship between prenatal bisphenol A (BPA) exposure and childhood overweight outcomes (Braun 2014, Valvi 2015) (moderate-certainty evidence).

##### 2.2.25.3 Prenatal or postnatal antibiotic exposure

Pooled analysis of four studies in a review by Srivastava et al. (2020) reported an association between prenatal (1^st^, 2^nd^ or 3^rd^ trimester) or postnatal (first two years of life) antibiotic exposure and increased risk of childhood overweight outcomes (moderate-certainty evidence).

##### 2.2.25.4 Postnatal antibiotic exposure

Two reviews by Miller et al. (2018b) and Srivastava et al. (2020) of six studies reported mixed findings for the association of postnatal (first two years of life) antibiotic exposure and childhood obesity and/or overweight. A pooled analysis of four studies in the review by

Srivastava et al. (2020) (very low-certainty evidence) reported no effect between postnatal antibiotic exposure and childhood obesity, and an additional study by Saari (2015) (moderate-certainty evidence) reported no statistically significant relationship between postnatal antibiotic exposure and childhood overweight. A pooled analysis of four studies in the review by Miller et al. (2018b) reported an association between postnatal antibiotic exposure and increased risk of childhood obesity or overweight (low-certainty evidence).

##### 2.2.25.5 Maternal smoking during pregnancy

Four studies in two reviews by Riedel et al. (2014) and Weng et al. (2012) reported mixed findings for the association of maternal smoking during pregnancy and childhood overweight, compared to no maternal smoking during pregnancy. Three studies reported an association between maternal smoking during pregnancy and childhood overweight outcomes (Dubois 2006, Fasting 2009, Hawkins 2009) (very low- to low-certainty evidence). One study reported no statistically significant relationship between this factor and childhood overweight (Heppe 2013) (moderate-certainty evidence).

##### 2.2.25.6 Paternal smoking during pregnancy

One study in the review by (Riedel et al. 2014) reported no statistically significant relationship between paternal smoking during pregnancy and childhood overweight, compared to no paternal smoking during pregnancy (Heppe 2013) (moderate-certainty evidence).

#### 2.2.26 Marketing or advertising

No relevant reviews were identified that reported marketing or advertising and any association with obesity or overweight in children less than five years old.

#### 2.2.27 Child’s screentime (quantitative evidence)

One review by Kwansa et al. (2022) examined the effect of child’s screentime as a factor for obesity or overweight in children less than five years old. A summary of the design and characteristics of the studies in the reviews can be found in Section 6.2, Table 19.

Comprehensive details of the results for this section are available in Table 16.

**Table 16:** Summary of reviews reporting child’s screentime as a factor associated with childhood overweight or obesity: adapted from GRADE.

| No. of primary studies (systematic review reference) | Certainty assessment |  |  |  |  |  | Impact (95% CI) | Certainty* |
| --- | --- | --- | --- | --- | --- | --- | --- | --- |
|  | Study design | Risk of bias | Inconsistency | Indirectness | Imprecision | Other considerations |  |  |
| Obesity or overweight outcomes |  |  |  |  |  |  |  |  |
| Watching TV > 2 hours per day: Obesity or overweight outcomes (follow-up: range 36 months to 60 months; assessed with: BMI or BMI-related measure [primary outcome]) |  |  |  |  |  |  |  |  |
| 1 Kwansa et al. (2022) | non-randomised studies | serious<sup>f</sup> | not serious | serious<sup>g</sup> | serious<sup>c</sup> | none | Sorrie (2017): OR: 4.01 (2.22 to 7.28)<br>Population studied: n = 500<br>**Associated with an increased risk** | ⊕○○○<br>Very low |
| *see Table 1 for definitions of GRADE certainty<br>BMI: body mass index; CI: confidence interval; n: number of study participants; OR: odds ratio<br>c. Wide confidence intervals around the point estimate of effect.<br>f. Cross-sectional study design used.<br>g. Downgraded for indirectness because studies did not include countries/setting(s)/population(s) likely to be directly relevant to the UK, or countries were not reported in the studies |  |  |  |  |  |  |  |  |

##### 2.2.27.1 Duration of television viewing

The study by Sorrie (2017) in the review by Kwansa et al. (2022) reported an association between watching more than two hours of television per day and childhood obesity or overweight outcomes (very low-certainty evidence).

##### 2.2.27.2 Eating whilst watching television

No relevant reviews were identified that reported eating whilst watching television and any association with obesity or overweight in children less than five years old.

##### 2.2.27.3 Computer game use

No relevant reviews were identified that reported computer game use and any association with obesity or overweight in children less than five years old.

#### 2.2.28 Childcare (quantitative evidence)

Two reviews examined the effect of childcare as a factor for obesity or overweight in children less than five years old (Alberdi et al. 2016, Black et al. 2017). A summary of the design and characteristics of the studies in the reviews can be found in Section 6.2, Table 19.

Comprehensive details of the results for this section are available in Table 17.

##### 2.2.28.1 Formal childcare

Four studies in one review by Black et al. (2017) reported mixed findings for the association between receiving formal childcare (at a childcare centre) and obesity and/or overweight.

One study reported a decreased risk of obesity in children receiving formal childcare and obesity (Koleilat 2012) (low-certainty evidence), two studies reported no statistically significant relationship between receiving formal childcare and overweight (Gubbels 2010) (very low-certainty evidence) or obesity/overweight (Tanskanen 2013) (very low-certainty evidence), and one study reported an association between receiving formal childcare and increased risk of overweight (Sata 2015) (very low-certainty evidence).

##### 2.2.28.2 Informal childcare

Three studies in two reviews by Alberdi et al. (2016) and Black et al. (2017) reported mixed findings for the association between receiving informal childcare and obesity and/or overweight. Two studies reported an association between receiving informal childcare (from grandparents) and overweight outcomes (Sata 2015) (very low-certainty evidence) or obesity/overweight outcomes (Tanskanen 2013) (low-certainty evidence).One study reported no statistically significant relationship between receiving informal childcare and obesity or overweight (Lehto 2016) (very low-certainty evidence), and one study reported no statistically significant relationship between receiving informal childcare (other than grandparents) and obesity or overweight (Tanskanen 2013) (very low-certainty evidence).

##### 2.2.28.3 Formal or informal childcare

A study by Zahir (2013), in the review by Alberdi et al. (2016), reported no statistically significant relationship between receiving informal or formal childcare and obesity or overweight (very low-certainty evidence).

##### 2.2.28.4 Time spent in childcare

A study by Pearce (2010b), in the review by Black et al. (2017), reported mixed findings for time spent in childcare and obesity or overweight. There was an association between full-time hours at informal childcare (grandparents and non-grandparents) and increased risk of obesity or overweight (moderate certainty-evidence), but no statistically significant relationship between full-time hours in formal childcare (low certainty-evidence). Part-time hours at informal (grandparent) childcare was associated with increased risk of obesity or overweight (moderate-certainty evidence), but there was no statistically significant relationship between part-time hours at informal (non-grandparents) and formal childcare (low-certainty evidence).

**Table 17:** Summary of reviews reporting childcare as a factor associated with childhood overweight or obesity: adapted from GRADE.

| No. of primary studies (systematic review reference) | Certainty assessment |  |  |  |  |  | Impact (95% CI) | Certainty |
| --- | --- | --- | --- | --- | --- | --- | --- | --- |
|  | Study design | Risk of bias | Inconsistency | Indirectness | Imprecision | Other considerations |  |  |
| Obesity outcomes |  |  |  |  |  |  |  |  |
| Formal childcare (childcare centre) (amount of time in childcare not reported): Obesity outcomes (follow-up: range 36 months to 48 months; assessed with: BMI or BMI-related measure [primary outcome]) |  |  |  |  |  |  |  |  |
| 1 Black et al. (2017) | non-randomised studies | serious<sup>d</sup> | not serious | not serious | not serious | publication bias strongly suspected<sup>b</sup> | Koleilat (2012): OR: 0.61 (0.41 to 0.90)<br>Population studied: n = 566<br>**Associated with a decreased risk** | ⊕⊕○○<br>Low |
| Overweight outcomes |  |  |  |  |  |  |  |  |
| Formal childcare (childcare centre) (amount of time in childcare not reported): Overweight outcomes (follow-up: 24 months; assessed with: BMI or BMI-related measure [primary outcome]) |  |  |  |  |  |  |  |  |
| 1 Black et al. (2017) | non-randomised studies | serious<sup>d</sup> | not serious | not serious | serious<sup>a</sup> | publication bias strongly suspected<sup>b</sup> | Gubbels (2010): OR: 1.19 (0.93 to 1.53)<br>Population studied: n = 2,396<br>**No statistically significant relationship** | ⊕○○○<br>Very low |
| Informal (grandparent) childcare versus parental childcare (amount of time in childcare not reported): Overweight outcomes (follow-up: 36 months; assessed with: BMI or BMI-related measure [primary outcome]) |  |  |  |  |  |  |  |  |
| 1 Black et al. (2017) | non-randomised studies | serious<sup>d</sup> | not serious | serious<sup>g</sup> | not serious | publication bias strongly suspected<sup>b</sup> | Sata (2015): OR: 2.1 (1.4 to 3.3)<br>Population studied: n = 4,281<br>**Associated with an increased risk** | ⊕○○○<br>Very low |
| Formal (nursery/kindergarten) childcare versus parental childcare (amount of time in childcare not reported): Overweight outcomes (follow-up: 36 months; assessed with: BMI or BMI-related measure [primary outcome]) |  |  |  |  |  |  |  |  |
| 1 Black et al. (2017) | non-randomised studies | serious<sup>d</sup> | not serious | serious<sup>g</sup> | not serious | publication bias strongly suspected<sup>b</sup> | Sata (2015): OR: 1.8 (1.2 to 2.7)<br>Population studied: n = 4,281<br>**Associated with an increased risk** | ⊕○○○<br>Very low |

| Obesity or overweight outcomes |  |  |  |  |  |  |  |  |
| --- | --- | --- | --- | --- | --- | --- | --- | --- |
| Full-time hours at informal (grandparent) childcare versus parental childcare: Obesity or overweight outcomes (follow-up: 36 months; assessed with: BMI or BMI-related measure [primary outcome]) |  |  |  |  |  |  |  |  |
| 1 Black et al. (2017) | non-randomised studies | not serious | not serious | not serious | not serious | publication bias strongly suspected<sup>b</sup> | Pearce (2010b): RR: 1.34 (1.12 to 1.60)<br>Population studied: n = 12,354<br>**Associated with an increased risk** | ⊕⊕⊕○<br>Moderate |
| Full-time hours at informal (other than grandparents) childcare versus parental childcare: Obesity or overweight outcomes (follow-up: 36 months; assessed with: BMI or BMI-related measure [primary outcome]) |  |  |  |  |  |  |  |  |
| 1 Black et al. (2017) | non-randomised studies | not serious | not serious | not serious | not serious | publication bias strongly suspected<sup>b</sup> | Pearce (2010b): RR: 1.40 (1.06 to 1.86)<br>Population studied: n = 12,354<br>**Associated with an increased risk** | ⊕⊕⊕○<br>Moderate |
| Full-time hours at formal childcare versus parental childcare: Obesity or overweight outcomes (follow-up: 36 months; assessed with: BMI or BMI-related measure [primary outcome]) |  |  |  |  |  |  |  |  |
| 1 Black et al. (2017) | non-randomised studies | not serious | not serious | not serious | serious<sup>a</sup> | publication bias strongly suspected<sup>b</sup> | Pearce (2010b): RR: 1.13 (0.94 to 1.34)<br>Population studied: n = 12,354<br>**No statistically significant relationship** | ⊕⊕○○<br>Low |
| Part-time hours at informal (grandparent) childcare versus parental childcare: Obesity or overweight outcomes (follow-up: 36 months; assessed with: BMI or BMI-related measure [primary outcome]) |  |  |  |  |  |  |  |  |
| 1 Black et al. (2017) | non-randomised studies | not serious | not serious | not serious | not serious | publication bias strongly suspected<sup>b</sup> | Pearce (2010b): RR: 1.15 (1.01 to 1.30)<br>Population studied: n = 12,354<br>**Associated with an increased risk** | ⊕⊕⊕○<br>Moderate |
| Part-time hours at informal (other than grandparent) childcare versus parental childcare: Obesity or overweight outcomes (follow-up: 36 months; assessed with: BMI or BMI-related measure [primary outcome]) |  |  |  |  |  |  |  |  |
| 1 Black et al. (2017) | non-randomised studies | not serious | not serious | not serious | serious<sup>a</sup> | publication bias strongly suspected<sup>b</sup> | Pearce (2010b): RR: 1.07 (0.87 to 1.32)<br>Population studied: n = 12,354<br>**No statistically significant relationship** | ⊕⊕○○<br>Low |
| Part-time hours at formal childcare versus parental childcare: Obesity or overweight outcomes (follow-up: 36 months; assessed with: BMI or BMI-related measure [primary outcome]) |  |  |  |  |  |  |  |  |
| 1 Black et al. (2017) | non-randomised studies | not serious | not serious | not serious | serious<sup>a</sup> | publication bias strongly suspected<sup>b</sup> | Pearce (2010b): RR: 1.09 (0.96 to 1.23)<br>Population studied: n = 12,354<br>**No statistically significant relationship** | ⊕⊕○○<br>Low |
| **Informal childcare versus other informal (parental) childcare (amount of time in childcare not reported): Obesity or overweight outcomes (follow-up: 36 months; assessed with: BMI or BMI-related measure [primary outcome])** |  |  |  |  |  |  |  |  |
| 1 Black et al. (2017) | non-randomised studies | very serious<sup>d,f</sup> | not serious | not serious | serious<sup>a</sup> | publication bias strongly suspected<sup>b</sup> | Lehto (2016): OR: 1.71 (0.41 to 7.19)<br>Population studied: n = 1,683<br>**No statistically significant relationship** | ⊕○○○<br>Very low |
| **Informal childcare (grandparent) versus parental childcare (amount of time in childcare not reported): Obesity or overweight outcomes (follow-up: 36 months; assessed with: BMI or BMI-related measure [primary outcome])** |  |  |  |  |  |  |  |  |
| 1 Alberdi et al. (2016), Black et al. (2017) | non-randomised studies | serious<sup>f</sup> | not serious | not serious | not serious | publication bias strongly suspected<sup>b</sup> | Tanskanen (2013): OR: 1.20 (1.03 to 1.40)<br>Population studied: n = 9,000<br>**Associated with an increased risk** | ⊕⊕○○<br>Low |
| **Informal childcare (other than grandparent) versus parental childcare (amount of time in childcare not reported): Obesity or overweight outcomes (follow-up: 36 months; assessed with: BMI or BMI-related measure [primary outcome])** |  |  |  |  |  |  |  |  |
| 1 Black et al. (2017) | non-randomised studies | serious<sup>f</sup> | not serious | not serious | serious<sup>a</sup> | publication bias strongly suspected<sup>b</sup> | Tanskanen (2013): OR: 1.09 (0.81 to 1.46)<br>Population studied: n = 9,000<br>**No statistically significant relationship** | ⊕○○○<br>Very low |
| **Formal childcare (childcare centre) versus parental childcare (amount of time in childcare not reported): Obesity or overweight outcomes (follow-up: 36 months; assessed with: BMI or BMI-related measure [primary outcome])** |  |  |  |  |  |  |  |  |
| 2 Black et al. (2017) | non-randomised studies | serious<sup>d,f</sup> | not serious | not serious | serious<sup>a</sup> | publication bias strongly suspected<sup>b</sup> | Tanskanen (2013): OR: 1.02 (0.87 to 1.19) ; Lehto (2016): OR: 1.27 (0.67 to 2.43)<br>Population studied: n = 10,683<br>**No statistically significant relationship** | ⊕○○○<br>Very low |
| **Informal or formal childcare (amount of time in childcare not reported): Obesity or overweight outcomes (follow-up: 48 months; assessed with: BMI or BMI-related measure [primary outcome])** |  |  |  |  |  |  |  |  |
| 1 Alberdi et al. (2016) | non-randomised studies | serious<sup>d</sup> | not serious | not serious | serious<sup>a</sup> | publication bias strongly suspected<sup>b</sup> | Zahir (2013): Obesity/overweight in childcare (47.7%); Normal weight in childcare (52.3%) (p = 0.56 for obesity and 0.98 for overweight)<br>Population studied: n = 201<br>**No statistically significant relationship** | ⊕○○○<br>Very low |
| *see Table 1 for definitions of GRADE certainty<br>BMI: body mass index; CI: confidence interval; n: number of study participants; OR: odds ratio; RR: risk ratio<br>a. Downgraded for imprecision due to insignificant p-values<br>b. Review authors report that publication bias was detected in the review<br>d. Review authors assessed studies as moderate and/or serious risk of bias<br>f. Cross-sectional study design used.<br>g. Downgraded for indirectness because studies did not include countries/setting(s)/population(s) likely to be directly relevant to the UK, or countries were not reported in the studies |  |  |  |  |  |  |  |  |

#### 2.2.29 Bottom line results for quantitative evidence

A wide range of biological, psychological, environmental and societal factors were identified as being associated with childhood obesity or overweight. Factors were associated with an increased risk, decreased risk, variable risk or no statistically significant effect with childhood obesity or overweight. Table 3 in Section 2.2 summarises the findings and certainty of the quantitative evidence.

#### 2.2.30 Bottom line results for qualitative evidence

Both systematic reviews referenced in our rapid qualitative evidence synthesis highlight the impact of cultural and familial influences on infant feeding practices across different demographics. Notably, this primary theme is closely related to the quantitative findings on feeding in infancy (Section 2.2.11), diet in childhood (Section 2.2.13) and caregiver behaviour (Section 2.2.18). A second main theme, parenting styles, was less represented or related to any of the quantitative evidence on factors associated with obesity/overweight. A key limitation of the evidence is that the review by Cartagena et al. (2014) focussed on the Hispanic population, and only three-out-of-10 studies included in Fraser et al. (2011) were UK studies, which may limit applicability of the overall findings and recommendations to Wales.

## 3. DISCUSSION

### 3.1 Summary of the findings

#### 3.1.1 Quantitative evidence

Twenty-eight systematic reviews reported the association between factors and obesity and/or overweight in children under five years. Most of these reviews included studies from Organization for Economic Cooperation and Development countries, and/or focused on predominantly Caucasian race/ethnicity, and so are likely to be of at least partial relevance to the UK, although generalisability will vary according to the specific country and setting included and the type of obesity/overweight-associated factors that were studied. Twelve reviews included studies conducted in the UK, from which the results are more likely to be transferable.

Most factors identified in the systematic reviews were consistently found to be associated with an increased risk of childhood obesity or overweight. The broad categories of those factors associated with an increased risk of childhood obesity and/or overweight include maternal weight, type of delivery, birthweight, growth rate, childhood diet, sleep duration, caregiver behaviour, socioeconomic status, education of parent, environmental exposure, screentime, and childcare. However, there was variability in the findings across sub-group factors within some of the broad categories (see Table 3). Four factors included evidence assessed as being of high certainty of being associated with an increased risk of childhood overweight using GRADE (see Table 1 for definitions of GRADE certainty). These are pre-pregnancy maternal overweight, maternal working hours (with an increment of 10 hours per week), rapid weight gain from zero to 12 months, and consumption of 1% milk compared to higher-fat milk. Most of the high-certainty evidence was on the association between factors and childhood overweight, and we did not identify any high-certainty evidence solely reporting childhood obesity.

There were only two factors in which all statistically significant studies in the review reported a decreased risk of the association with childhood obesity or overweight (breastfeeding and larger household size).

A similar number of factors were reported as having mixed findings or no statistically significant relationship with childhood obesity or overweight.

Table 3, in Section 2.2, summarises these findings and certainty.

The findings in this report may differ from other published reviews due to differences in the scope of what was studied. Of particular note to UK policy making in this area are two recent reports from the Scientific Advisory Committee on Nutrition (SACN), on feeding in the first year of life (The Scientific Advisory Committee on Nutrition 2018), and on feeding young children aged one to five years old (The Scientific Advisory Committee on Nutrition 2023).

The findings in our review specifically focused on factors associated with obesity and/or overweight when both the factor and its impact on weight status were studied in children under five years of age. Consequently, we did not include evidence where weight outcomes were only reported in older children (age five years and above) or where the age of the subjects was unclear. In contrast, the SACN reviews have a broader scope, examining the influence of factors on weight outcomes at any age, not just in those under five years. For example, some of the studies investigating protein intake measured the odds of being overweight at seven years old (The Scientific Advisory Committee on Nutrition 2023).

The outcomes studied in our review differ from those in the SACN reviews. Our review focused on evidence that specifically classified children as overweight or obese using BMI (or other well-accepted measures for children under two years), and did not include any studies that used surrogate measures for obesity or overweight, or that reported increases in BMI without clarifying the impact on the overall risk or incidence of overweight or obesity. In contrast, the SACN reviews cover a broad spectrum of weight-related outcomes. For example, some of the studies investigating milk consumption as a factor reported the difference in BMI z-score after a certain follow-up time, but with no specific mention of the risk or incidence of obesity/overweight (The Scientific Advisory Committee on Nutrition 2023).

#### 3.1.2 Qualitative evidence

Cartagena et al. (2014) found that differing infant feeding practices can have a large impact on a child’s risk of being overweight or obese. The study also highlights the complex interplay between ethnicity, acculturation, breastfeeding practices, and infant health outcomes. Rooted cultural perceptions can also heavily shape maternal attitudes towards infant weight status. This perception often leads to overfeeding and early introduction of solid foods, contributing to an increased risk of childhood overweight and obesity. The findings also highlight the importance of culturally-sensitive interventions that address familial influences and maternal perceptions, to promote healthy feeding practices and prevent childhood obesity within the Hispanic community.

Fraser et al. (2011) explored factors associated with the increase in childhood obesity, including paternal influences, parenting styles, eating and physical activity habits, and knowledge about nutrition and mealtime behaviours. Overall, the findings of this systematic review emphasise the importance of fathers in child development and weight management. Studies within Fraser et al. (2011) show that paternal parenting styles, characterised by both firmness and supportiveness, can significantly impact children’s weight outcomes, with greater paternal acceptance correlating with reduced childhood obesity. Controlling behaviours during feeding may also contribute to higher body fat percentages in children.

However, the relationship between the controlling behaviour during feeding such as restriction and children’s weight was still unclear.

### 3.2 Strengths and limitations of the available evidence and this rapid review

#### 3.2.1 Strengths

A large number of quantitative reviews were identified, with all studies including a large number of participants. Of the 24 classes of factors we identified as potentially associated with childhood obesity or overweight, we were able to identify evidence that met our inclusion criteria for the majority (14/24). The interactive evidence maps in Figure 2 highlight the breadth and complexity of the evidence identified, and show gaps in the evidence.

Most of the studies were conducted in countries deemed relevant to the UK, as they were either Organization for Economic Cooperation and Development countries, and/or focused on predominantly Caucasian race/ethnicity. Only two reviews were identified as likely to be not relevant to the UK. Thirteen out of 30 reviews included studies conducted in the UK, but none of the studies specifically mentioned Wales. Three out of the ten studies included in the one qualitative study that included UK studies were conducted in the UK. The majority of the reviews also measured childhood obesity or overweight using measures defined in our primary outcomes (see Methods section for details), which we deemed to be the most reliable and direct measures of obesity or overweight, rather than reliance on secondary ‘surrogate’ outcomes.

#### 3.2.2 Limitations

As this is a review of existing systematic reviews, we relied largely on the reporting and conclusions of the authors of the included reviews. To ensure our findings were meaningful and robust, we only included reviews that met suitable reporting standards for systematic reviews, and that included all information required for our review. We judged many potentially useful reviews as incomplete or not robust enough to use (for example, because of poor reporting of outcome data, or lack of suitable assessment of risk of bias). Whilst many of these reviews may contain potentially useful evidence, extraction of this evidence would require a more thorough and resource-intensive approach that is beyond the scope of this report.

The majority of reviews included cohort studies. Four of the reviews included cross-sectional studies, which cannot establish cause-and-effect. Beyond classification of study design, this rapid review relied on judgements about risk of bias of included studies made by the authors of the reviews themselves. Although we only used reviews that used a recognised critical-appraisal tool and reported their judgements in full, this still results in some likely variability in how risk of bias was assessed between reviews. It also limits our ability to comment on factors of particular importance regarding certainty. For example, controlling for confounding variables in the included primary studies would increase the certainty of the evidence they report, but it is not clear to what extent this was done in each study; some, but not all critical-appraisal tools used by review authors will account for this. Because a more thorough analysis of this issue would require detailed scrutiny of individual studies at a level that is beyond the scope of a review of systematic reviews, a causal relationship between the factors and childhood obesity or overweight cannot be established.

Although the rapid approach of this review precluded any rigorous assessment of overlap between the included systematic reviews, initial observations suggest that there was not as much overlap as we expected. The reasons why there may not be as much overlap between systematic reviews as we anticipated remain unclear.

We found no suitable reviews reporting the relationship between obesity/overweight in children under five and biological factors, paternal weight status, maternal diet or nutrition, maternal mental health, maternal physical activity, gestational age at birth, childhood physical activity, COVID-19 related school closures, neglect or abuse, caregiver involvement in children’s healthy eating or physical activity interventions, dietary inflammatory potential, food insecurity, food prices, eating whilst watching television, computer game use, built environment, and marketing or advertising, associated with childhood obesity or overweight. While such reviews likely exist, they were not considered suitable for inclusion in this analysis. Similarly, there could be primary studies on these topics that have not yet been examined by any comprehensive, well-conducted systematic review.

### 3.3 Evidence into practice opportunities

The evidence identified suggests that it is important to consider a wide range of biological, psychological, environmental and societal factors when developing interventions or actions to address the reasons for childhood obesity or overweight in children younger than five years.

The high-certainty evidence supports:

- Helping overweight women, who are thinking about having a baby or trying to conceive, to lose weight. Note that there was also low-certainty evidence for this factor reported by some studies.
- Reducing rapid weight gain during the first 12 months of life. Note that there was also moderate- and low-certainty evidence for this factor reported by some studies.
- Providing opportunities for children of working mothers to access healthier foods and be more physically active, particularly if the mothers work long hours.

The moderate-certainty supports:

- Promoting breastfeeding.
- Reducing rapid weight gain during the first 13 months of life.
- Monitoring the child’s growth rate during the first two years of life, particularly of babies with catch-up growth
- Promoting baby-led weaning (spoon-feeding and purees for 10% or less of the time).
- Focus on reducing the regular consumption of sugar-containing beverages during childhood.
- Educating and supporting grandparents and other informal caregivers to provide healthier foods and opportunities for play or physical activity for children.

Other factors with lower-certainty evidence may be useful to consider or acknowledge when developing interventions to address childhood obesity or overweight, including: babies born large for their age and sex; babies born by caesarean delivery; consumption of sweet food, fried or fatty food, or fast food during childhood; high dietary diversity during childhood; maternal under-evaluation of a child’s weight; children with wealthier parents; children watching television for more than two hours per day; household size; maternal pre-pregnancy underweight; and timing of weaning.

The qualitative evidence identified synthesis highlight the impact of cultural and familial influences on infant feeding practices across different demographics. The evidence suggests that interventions addressing breastfeeding support and healthy dietary practices may be beneficial. The findings also highlight the importance of culturally-sensitive interventions that

address familial influences and maternal perceptions, to promote healthy feeding practices and prevent childhood obesity within the Hispanic community. Interventions could focus on educating both mothers and extended family members on the importance of responsive feeding and appropriate feeding cues, although the sole focus of the evidence was on the Hispanic community and interventions would likely need to be altered to best fit the Welsh population and cultural settings.

The evidence also highlights the need for a comprehensive understanding of familial influences on children’s eating behaviours and confirms the importance of involving both parents in interventions aimed at promoting healthy eating habits and preventing childhood obesity. These findings imply the need for comprehensive strategies to promote healthy eating habits and overall well-being in children, including setting limits, establishing consistent routines, anticipating the child’s needs, interpreting nonverbal cues, maintaining physical and emotional closeness, and modelling desirable behaviours.

### 3.4 Implications for future research

- Most of the high-certainty evidence related to childhood overweight. There was some high-certainty evidence related to childhood obesity or overweight, but no high-certainty evidence was identified on the association between factors and childhood obesity. More robust primary studies, and/or reviews of these studies, clearly reporting methods, outcomes and risk of bias, are needed investigating factors associated with childhood obesity.
- Whilst we reported outcomes for children younger than five years, all of the selected reviews also included studies with children older than five years. More reviews solely focusing on children younger than five years would help provide more informative evidence on factors associated with childhood obesity or overweight in this age group.
- We found limited evidence regarding obesity or overweight outcomes in children under two years old (6/30 studies included children under two years). Conducting studies focused on measuring obesity or overweight in this age group could help address this gap, offering valuable insights into potential risk factors identifiable from a very young age.
- Limited evidence was identified on the role of fathers in the development of childhood obesity or overweight.
- Limited qualitative evidence was identified, including limited qualitative evidence from the UK. However, the systematic reviews of qualitative evidence we did identify underscore the influence of cultural and familial factors on infant feeding practices across various demographics. This central theme aligns closely with quantitative data on feeding in infancy, childhood diet, and caregiver behaviour. These insights stress the necessity of fully understanding how family influences children’s eating habits. Given that little of this evidence was UK-focussed, there is potential for this to be explored further in a UK setting.
- Many of the studies identified explored associations between factor(s) of interest and childhood obesity or overweight, but given that many factors are interrelated and given the study design of many of the studies, it is difficult to conclude whether these are causative relationships. More experimental studies which can better establish cause-and-effect relationships between factors and childhood obesity or overweight are needed.
- Our examination of current systematic reviews revealed inconsistent evidence regarding the association of certain factors with obesity/overweight, and a complete lack of evidence for other factors. As mentioned in section 3.2.2, our methodology probably means that some primary evidence capable of addressing or clarifying these discrepancies was not included in our review. Conducting more focussed systematic reviews, containing detailed analysis of individual factors or a limited number of closely related factors, could address this.

### 3.5 Economic considerations*

- Experience of various measures of childhood poverty have been identified as a key factor explaining variation in prevalence of obesity and overweight in children of reception age (four to five years) (Cheung et al. 2022). Food costs are a considerable factor in purchasing decisions, especially for families in which children experience poverty. To comply with NHS Eatwell guidance, the most deprived household quintile would have to spend 50% of their disposable income on food (Goudie 2023).
- In the UK, local authorities with the greatest proportion of children aged under-five years living in households in receipt of out-of-work benefits have on average 3.5% more children aged three to five years who are obese or overweight compared to local authorities with the lowest proportion (Cheung et al. 2022).
- Childhood obesity or overweight is positively associated with increased lifetime healthcare resource utilisation, resulting in increased healthcare associated costs (Hasan et al. 2020, Okunogbe et al. 2021). Systematic review evidence suggests medication and healthcare costs are greater in adolescents (12 to 18 years) with obesity or overweight than young children (zero to five years). However, nonhospital healthcare costs (such as out-of-pocket expenses) were reported to be significantly higher in the zero to five year’s cohort (Ling et al. 2023).

*\*This section has been completed by the Centre for Health Economics & Medicines Evaluation (CHEME), Bangor University*

## Data Availability

All data produced in the present study are available upon reasonable request to the authors

## Glossary

**Bias**: Systematic (as opposed to random) deviation of the results of a study from the ’true’ results, which is caused by the way the study is designed or conducted.

**Catch-up growth**: in the first two years after life, a rapid rate of growth after being born small for gestational age or experiencing slow growth after birth. Definitions vary, but two systematic reviews in this rapid review define it as an increase in weight-for-age z-score greater than 0.67 standard deviation units during the first two years of life (from birth to two years).

**Cohort study**: an observational study with two or more groups (cohorts) of people with similar characteristics. One group is exposed to a risk factor and the other group is not. The study records their progress over time and records what happens.

**Cross-sectional study**: A ’snapshot’ observation of a set of people at one time.

**Confidence interval**: A way of expressing how certain we are about the findings from a study, using statistics. It gives a range of results that is likely to include the ’true’ value for the population. A wide confidence interval indicates a lack of certainty about the true effect of the test or treatment.

**Dietary inflammatory potential**: refers to the effect of diet on the levels of inflammatory factors in the body.

**Grading of Recommendations, Assessment, Development and Evaluation (GRADE)**: a common approach to grading the certainty of evidence. Definitions of the certainty of the evidence can be found in Table 1. See www.gradeworkinggroup.org for more information.

**High-certainty evidence**: We are very confident that the true effect lies close to that of the estimate of the effect.

**Imprecision**: issues such as measuring a treatment effect based on very few patients or effects.

**Inconsistency of results**: unexplained heterogeneity across studies.

**Indirectness**: how applicable the evidence is to the research question and population of interest.

**Low-certainty evidence**: Our confidence in the effect estimate is limited: The true effect may be substantially different from the estimate of the effect.

**Moderate-certainty evidence**: We are moderately confident in the effect estimate: The true effect is likely to be close to the estimate of the effect, but there is a possibility that it is substantially different.

**Organization for Economic Cooperation and Development countries:** A unique forum where the governments of 37 democracies with market-based economies collaborate to develop policy standards to promote sustainable economic growth.

**P value**: a statistical measure that indicates whether or not an effect is statistically significant. By convention, if the p value is below 0.05 (that is, there is less than a 5% probability that the results occurred by chance), it is considered that there probably is a real difference between treatments or other measured effects.

**Publication bias**: Publication bias occurs when researchers publish the results of studies showing that a treatment works well and do not publish those results showing it did not have any effect.

**Qualitative evidence**: explores people’s beliefs, experiences, attitudes, behaviour and interactions. It asks questions about how and why.

**Quantitative evidence**: Research that generates numerical data or data that can be converted into numbers.

**Rapid review**: a systematic review accelerated through streamlining or omitting specific methods.

**Risk of bias**: also called study limitations. Limitations in the study design and execution may bias the estimates of the treatment effect.

**Statistical significance**: A statistically significant result is one that is assessed as being due to a true effect rather than random chance.

**Study design:** whether evidence is from randomised controlled trials, observational studies, or other sources

**Systematic review**: a review that summarises the evidence on a clearly formulated review question according to a predefined protocol, using systematic and explicit methods to identify, select and appraise relevant studies, and to extract, analyse, collate and report their findings.

**Very-low certainty evidence**: We have very little confidence in the effect estimate: The true effect is likely to be substantially different from the estimate of effect.

## Acknowledgements

The authors would like to thank Emily Finney, Alex Hicks, Nathan Davies and Ed Wilson for their time, their expertise and contributions during stakeholder meetings, and in guiding the focus of the review.

## 5. RAPID REVIEW METHODS

### 5.1 Initial exploration of the evidence

Prior to planning this rapid review, a preliminary scoping search for existing evidence was undertaken using TRIP, Medline, and the Cochrane Database for Systematic Reviews (CDSR), using the keywords obesity, overweight, child/childhood/infant/preschool, risk factors, determinants, causative factors, treatment, management, interventions. This aimed to identify evidence on factors associated with increased levels of childhood obesity/overweight in children up to 5 years old and interventions to address these. The findings were presented to the Stakeholders and used to decide on the scope of the rapid review, and to inform the methods. It was decided that the rapid review should provide an overview of all potential causes of childhood obesity and indicate where there is evidence to support their association with obesity. This would take the form of a ‘review of existing reviews’, mapping possible causes to the evidence available, its certainty, and applicability to Wales.

A preliminary literature search for the rapid review indicated that there would likely be a large volume of potentially relevant evidence. We therefore undertook an interim analysis (or evidence map) of the reviews identified during the preliminary search, to inform a more substantive focus for the rapid review. As part of the preliminary literature search, we included all relevant quantitative and qualitative secondary evidence. We included any factors associated with childhood obesity or overweight found in the reviews, or reported them as a possible association, and used this information to determine as many potentially relevant factors to include in our own review as possible, and to develop our own classification system for these. At this stage we included any reviews that included data, or at least some data, on preschool-age children. We ranked evidence according to age of the participants, and their likely relevance in descending order:

1. Category 1: Reviews that only study children < 5 years
2. Category 2: Reviews that include older ages, but report results and findings for children < 5 years separately
3. Category 3: Reviews that include older ages, but where a majority of the overall population (we suggested more than 75%) are children < 5 years
4. Category 4: Reviews that include older ages, but the age range included may allow relevant conclusions to be drawn on the under-5 age group. This includes scenarios such as where studies included covered an age-range outside but only slightly above our age range of interest (typically, we used this classification if all children were aged up to 5 or up to 6 years); or where results and findings for children < 5 years were reported separately in a review that included other ages, but a very small or unclear proportion of the evidence was in our age group of interest

We also classified evidence according to the relevance of the study population/settings/countries to Wales: a review was classified as relevant if the majority of the studies in the review include Organization for Economic Cooperation and Development countries, and/or focus on predominantly Caucasian race/ethnicity.

At this stage, we extracted other basic information about the design of the review and the studies it included, broadly reflecting the data presented here in Section 6.2, but we did not extract outcomes or findings of the reviews.

After discussion of these findings with stakeholders, it was concluded that we should, as part of the proposed rapid review, only include reviews that report data separately for children less than five years old (i.e. categories one and two within our framework, described above), but that we should include all settings/populations/countries. Due to the large volume of secondary evidence identified, it was agreed that primary evidence would not be included.

### 5.2 Eligibility criteria

**Table 18:**
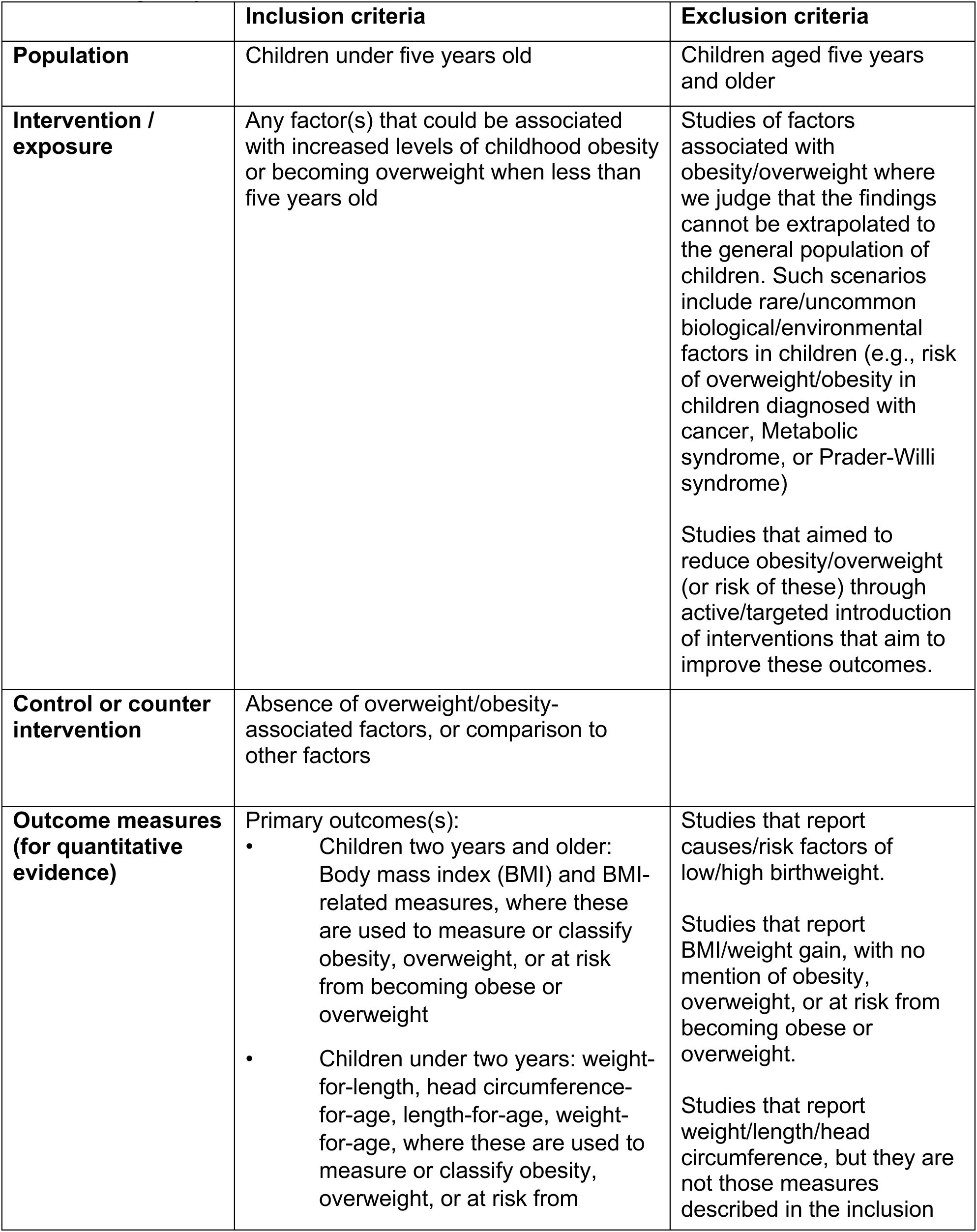
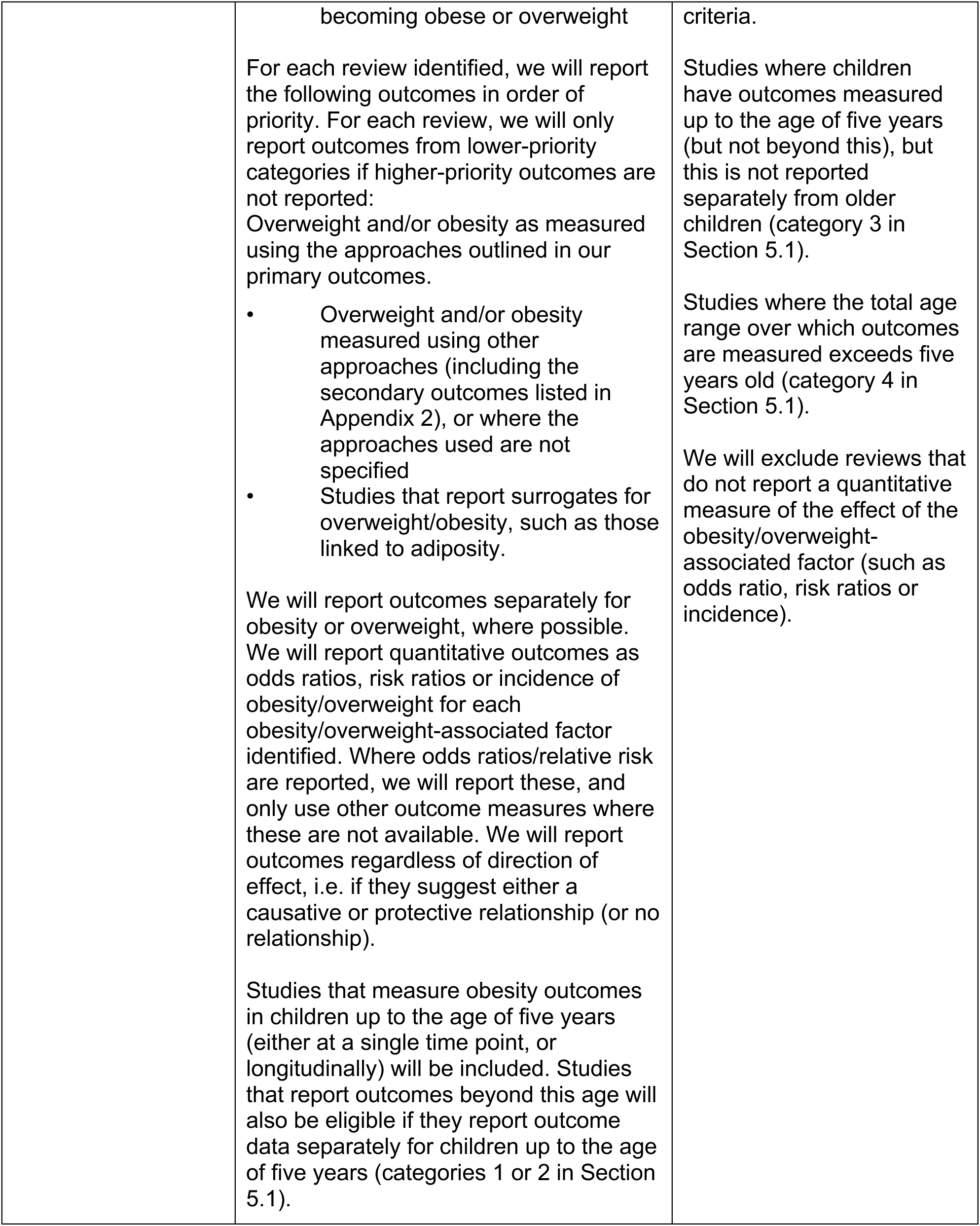
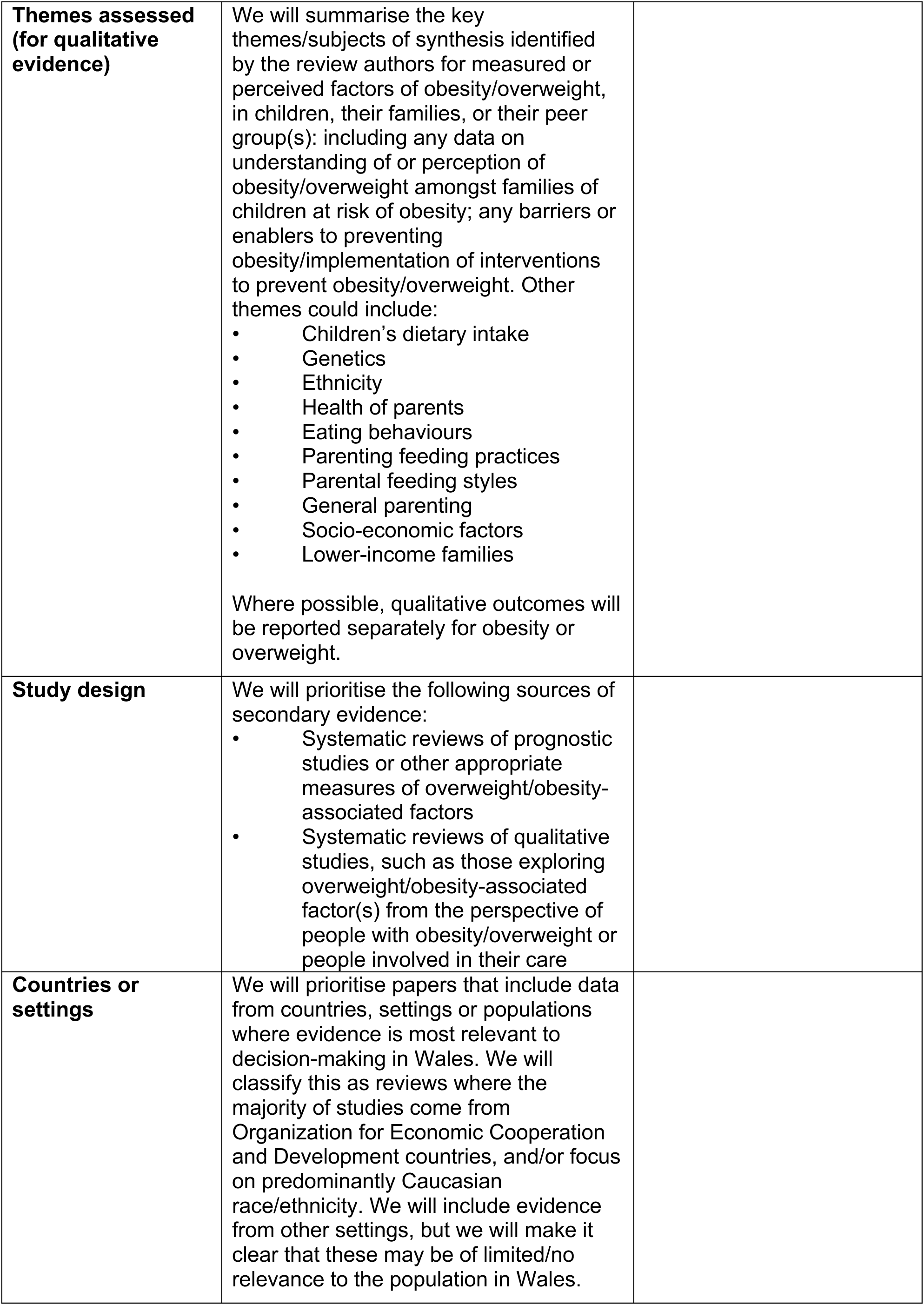
Eligibility criteria.

|  | **Inclusion criteria** | **Exclusion criteria** |
| --- | --- | --- |
| **Population** | Children under five years old | Children aged five years and older |
| **Intervention / exposure** | Any factor(s) that could be associated with increased levels of childhood obesity or becoming overweight when less than five years old | Studies of factors associated with obesity/overweight where we judge that the findings cannot be extrapolated to the general population of children. Such scenarios include rare/uncommon biological/environmental factors in children (e.g., risk of overweight/obesity in children diagnosed with cancer, Metabolic syndrome, or Prader-Willi syndrome)<br>Studies that aimed to reduce obesity/overweight (or risk of these) through active/targeted introduction of interventions that aim to improve these outcomes. |
| **Control or counter intervention** | Absence of overweight/obesity-associated factors, or comparison to other factors |  |
| **Outcome measures (for quantitative evidence)** | Primary outcomes(s):<br>Children two years and older: Body mass index (BMI) and BMI-related measures, where these are used to measure or classify obesity, overweight, or at risk from becoming obese or overweight<br>Children under two years: weight-for-length, head circumference-for-age, length-for-age, weight-for-age, where these are used to measure or classify obesity, overweight, or at risk from | Studies that report causes/risk factors of low/high birthweight.<br>Studies that report BMI/weight gain, with no mention of obesity, overweight, or at risk from becoming obese or overweight.<br>Studies that report weight/length/head circumference, but they are not those measures described in the inclusion |
|  | becoming obese or overweight<br>For each review identified, we will report the following outcomes in order of priority. For each review, we will only report outcomes from lower-priority categories if higher-priority outcomes are not reported:<br>Overweight and/or obesity as measured using the approaches outlined in our primary outcomes.<br>• Overweight and/or obesity measured using other approaches (including the secondary outcomes listed in Appendix 2), or where the approaches used are not specified<br>• Studies that report surrogates for overweight/obesity, such as those linked to adiposity.<br>We will report outcomes separately for obesity or overweight, where possible. We will report quantitative outcomes as odds ratios, risk ratios or incidence of obesity/overweight for each obesity/overweight-associated factor identified. Where odds ratios/relative risk are reported, we will report these, and only use other outcome measures where these are not available. We will report outcomes regardless of direction of effect, i.e. if they suggest either a causative or protective relationship (or no relationship).<br>Studies that measure obesity outcomes in children up to the age of five years (either at a single time point, or longitudinally) will be included. Studies that report outcomes beyond this age will also be eligible if they report outcome data separately for children up to the age of five years (categories 1 or 2 in Section 5.1). | criteria.<br>Studies where children have outcomes measured up to the age of five years (but not beyond this), but this is not reported separately from older children (category 3 in Section 5.1).<br>Studies where the total age range over which outcomes are measured exceeds five years old (category 4 in Section 5.1).<br>We will exclude reviews that do not report a quantitative measure of the effect of the obesity/overweight-associated factor (such as odds ratio, risk ratios or incidence). |
| **Themes assessed (for qualitative evidence)** | We will summarise the key themes/subjects of synthesis identified by the review authors for measured or perceived factors of obesity/overweight, in children, their families, or their peer group(s): including any data on understanding of or perception of obesity/overweight amongst families of children at risk of obesity; any barriers or enablers to preventing obesity/implementation of interventions to prevent obesity/overweight. Other themes could include:<br>• Children's dietary intake<br>• Genetics<br>• Ethnicity<br>• Health of parents<br>• Eating behaviours<br>• Parenting feeding practices<br>• Parental feeding styles<br>• General parenting<br>• Socio-economic factors<br>• Lower-income families<br>Where possible, qualitative outcomes will be reported separately for obesity or overweight. |  |
| **Study design** | We will prioritise the following sources of secondary evidence:<br>• Systematic reviews of prognostic studies or other appropriate measures of overweight/obesity-associated factors<br>• Systematic reviews of qualitative studies, such as those exploring overweight/obesity-associated factor(s) from the perspective of people with obesity/overweight or people involved in their care |  |
| **Countries or settings** | We will prioritise papers that include data from countries, settings or populations where evidence is most relevant to decision-making in Wales. We will classify this as reviews where the majority of studies come from Organization for Economic Cooperation and Development countries, and/or focus on predominantly Caucasian race/ethnicity. We will include evidence from other settings, but we will make it clear that these may be of limited/no relevance to the population in Wales. |  |
| **Language of publication** | English |  |
| **Publication date** | None | None |
| **Publication type** | Published and preprint |  |
| **Other factors** | Where possible we will report evidence separately for the following subgroups:<br>• Specific age ranges within preschool children<br>• Preconception/pregnancy |  |

### 5.3 Literature search

A comprehensive search was conducted in December 2023 to identify any additional English-language evidence (not identified in the preliminary searches), focussing on identifying secondary evidence for the reasons described above. There was no literature search start date. The databases searched were: Medline, American Psychological Association PsychInfo, Embase, Cumulated Index to Nursing and Allied Health Literature (CINAHL), Cochrane, Epistemonikos.

The Medline search for English-language reviews can be found in Appendix 1.

### 5.4 Reference management

All citations retrieved from the database searches were imported or entered manually into EndNote^TM^ (Thomson Reuters, CA, USA) and duplicates removed by a single reviewer. The citations that remained were exported as an TXT file and then imported to Rayyan^TM^ for study selection.

### 5.5 Study selection process

Two reviewers screened 20% of titles and abstracts independently. After this, the level of agreement was assessed with disagreements settled by discussion and consensus. Both reviewers had to achieve at least 80% agreement on screened records before progressing to the next stage. The remaining titles and abstracts were screened by the primary reviewer alone. 20% of full texts were screened by both reviewers, with the same agreement threshold (80%) as before necessary before the remaining records could be screened by the primary reviewer alone. During independent screening, the primary reviewer consulted with the secondary reviewer in the case of any uncertainties.

### 5.6 Data extraction

Data extraction took place in two parts. For the first part, data were extracted and added to an Excel spreadsheet, where it was coded according to obesity/overweight-associated factor, age and relevance to the UK (as described in Section 5.1). Data extracted in this first part of the process included: study author, year, DOI/link, obesity/overweight-associated factor and coding, age range at which outcome measured and coding of relevance, countries/settings of studies and coding of relevance, and number of studies in the review.

For the second part of the data extraction process, data extraction was based on the final outlined eligibility criteria. A standardised data extraction form was created to capture key information about the study and findings relevant to the review question.

The following data were extracted where available:

- Study information (author, year, country)
- Population characteristics (age, gender, ethnicity, socioeconomic status, specific vulnerabilities)
- Measurement of weight and/or obesity. e.g., BMI and percentile charts.
- Qualitative data regarding obesity/overweight-associated factor(s), e.g., parental feeding practices and diet
- Other methods used for the evaluation of outcomes.

Data were extracted by a single reviewer and quality assured by a second reviewer.

### 5.7 Study design classification

Because we only included systematic reviews, we utilised and reported review authors’ own classifications of study design. In cases of any ambiguity around the design of a primary study that was included in a systematic review, we retrieved the full text of the primary study and classified the study based on the primary study authors’ description of the study and their methods.

### 5.8 Critical appraisal of the evidence

We did not formally assess the risk of bias of the included systematic reviews, but we ensured that they met a certain minimum standard of conduct. We only included reviews that undertook critical appraisal using a suitable tool and report the findings in a usable format.

We also only included reviews if they systematically searched more than one database for evidence. We reported the findings of the systematic review authors’ own conclusions on rigour and quality assessment of the studies they included, based on the use of a suitable critical-appraisal tool.

### 5.9 Synthesis

We undertook narrative synthesis of the evidence identified based on the selection criteria outlined above, and also utilised the meta-analyses in some included reviews. Summary tables were provided where possible (e.g., types of factors or for age groups). EPPI-Mapper^TM^ was used to present data more visually in the form of an evidence map.

Rapid qualitative evidence synthesis was done based on the recommendations of Booth (2024), focussing on identifying themes found in one or more review and synthesising findings on each theme.

### 5.10 Assessment of body of evidence

We assessed certainty in the body of evidence of outcomes derived from the quantitative evidence using a modified version of GRADE (The Grading of Recommendations, Assessment, Development, and Evaluations) (Higgins JPT 2023). The GRADE approach provides a formal system to categorise the certainty of the evidence for each outcome or effect into one of four levels: high, moderate, low and very low. For the purpose of this review, we assessed each individual study for how certain the findings are, rather than assess the overall body of evidence for each factor, which is how GRADE is generally applied. The definitions of these GRADE ratings are provided in Table 1.

The system for assessing the certainty of evidence includes five domains relating to: risk of bias across the studies, inconsistency (heterogeneity) of the results between studies, indirectness (including subgroup analyses and applicability of the outcome measure), imprecision of the result (number of events, and width of the confidence intervals), and publication bias (Higgins JPT 2023). At the start it is assumed that (multiple) cohort prognostic studies provide high certainty level evidence and cross-sectional studies low-certainty evidence. This evidence rating is then downgraded based on the assessment of the different domains. The evidence rating can also be upgraded in the presence of: large magnitude of effect, dose response, or when plausible confounding factors are unlikely to increase the effect (Higgins JPT 2023).

We used systematic review authors’ own risk-of-bias assessments to inform this and carried out our own assessment of the other domains that influence certainty. A summary of the criteria that we used to assess the certainty of the evidence from the systematic reviews are:

- Study design: We assigned cohort studies a high-certainty status for this domain, as very few randomised controlled trials were identified. The evidence rating was downgraded where a cross-sectional study design was used.
- Risk of bias: The evidence rating was downgraded if the review authors assessed studies as moderate and/or serious risk of bias.
- Inconsistency: The evidence rating was downgraded where there was moderate or substantial variability in effect sizes across studies.
- Indirectness: The evidence rating was downgraded where studies did not include countries/setting(s)/population(s) likely to be directly relevant to the UK, or countries were not reported in the studies.
- Imprecision: The evidence rating was downgraded where wide confidence intervals were reported around the effect estimate, relating to either an increased or decreased risk of childhood obesity/overweight; the rating was also downgraded for imprecision due to insignificant p-values.
- Publication bias: The evidence rating was downgraded where the review authors report that publication bias was detected in the review.

The initial confidence rating was downgraded one GRADE category each time the available evidence (for each outcome) was assessed as having one of these criteria.

## 6. EVIDENCE

### 6.1 Search results and study selection

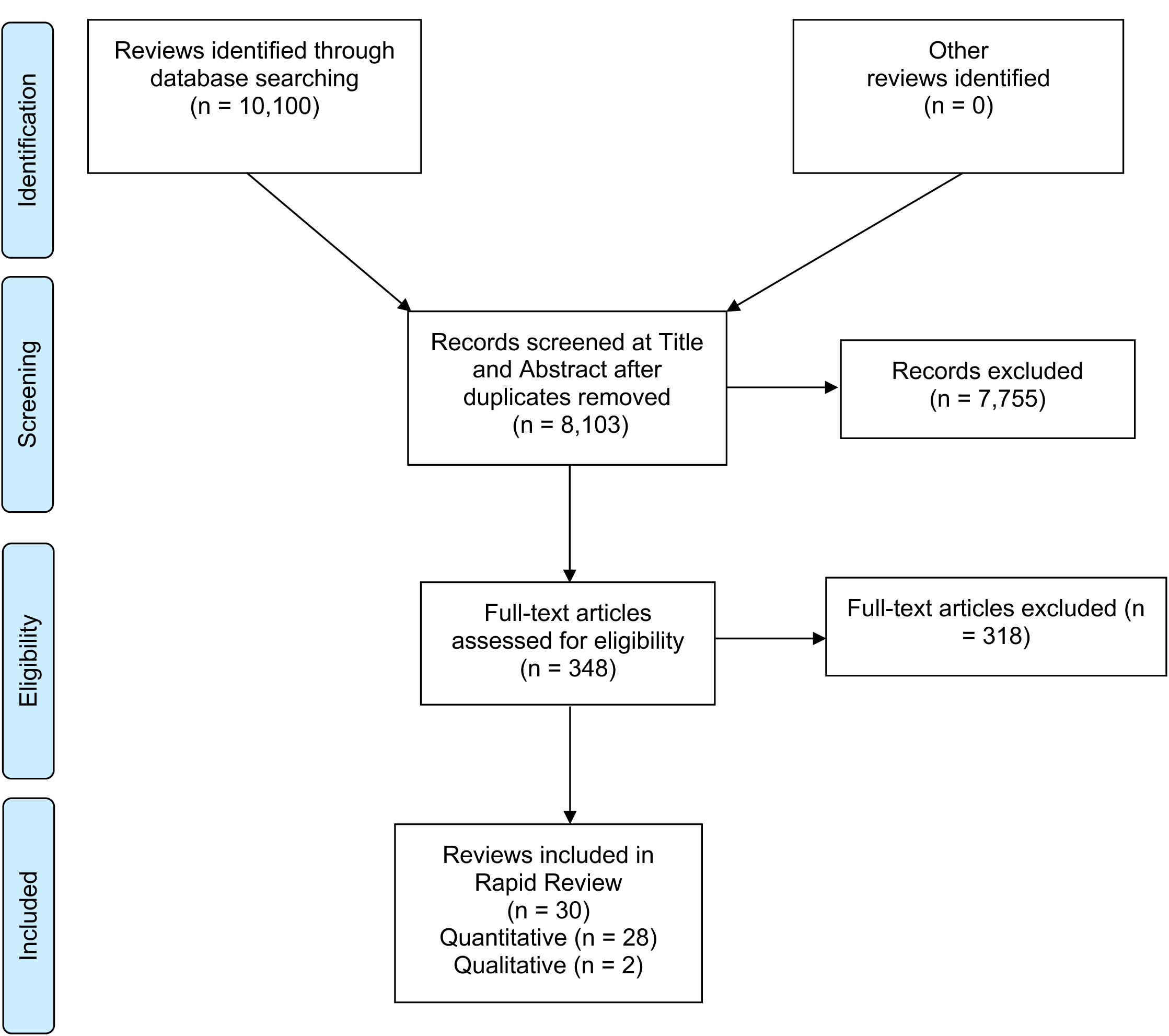

### 6.2 Data extraction

All systematic reviews included in the rapid review are listed in Table 19 (quantitative) and Table 20 (qualitative)

**Table 19:**
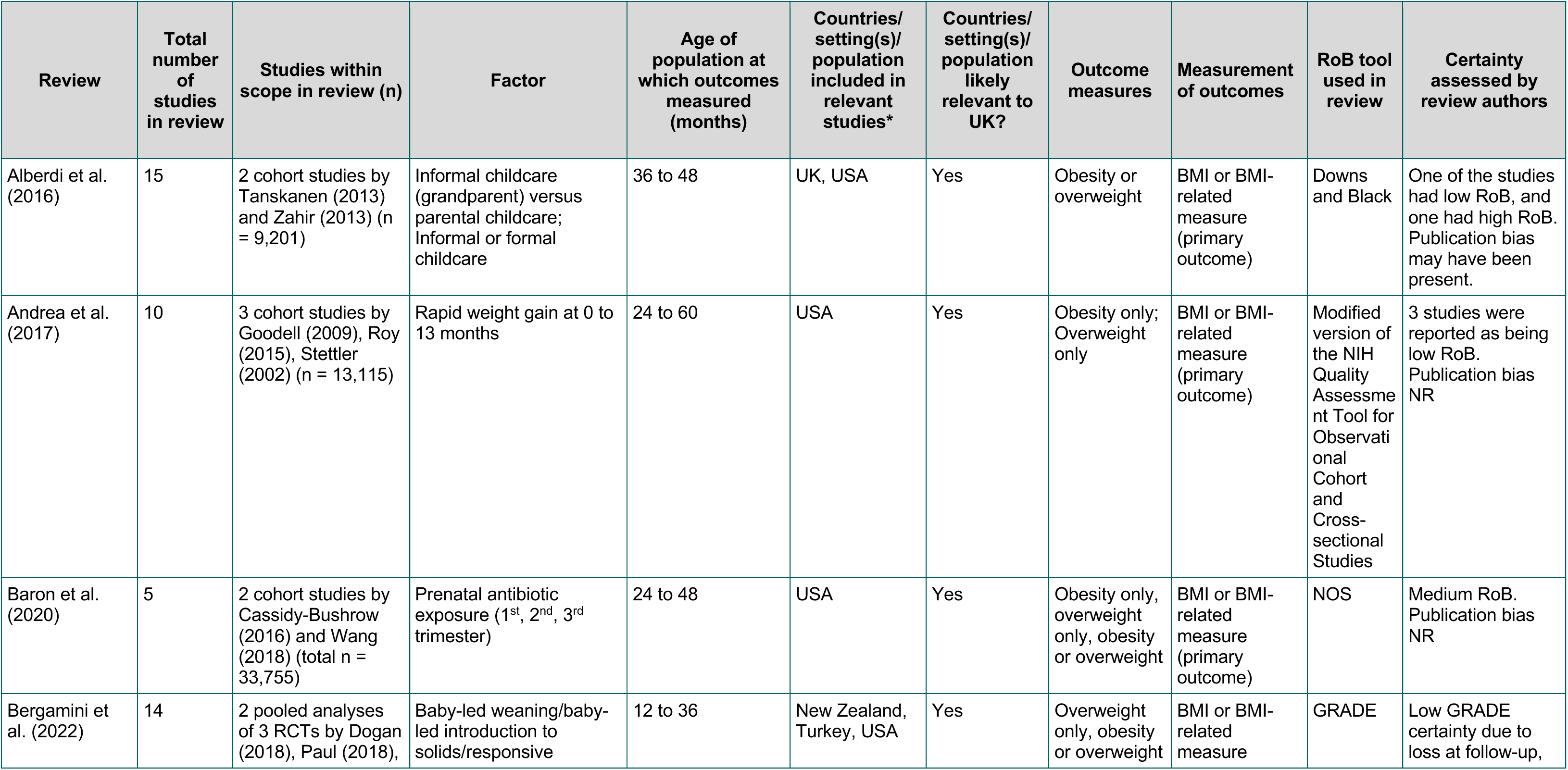
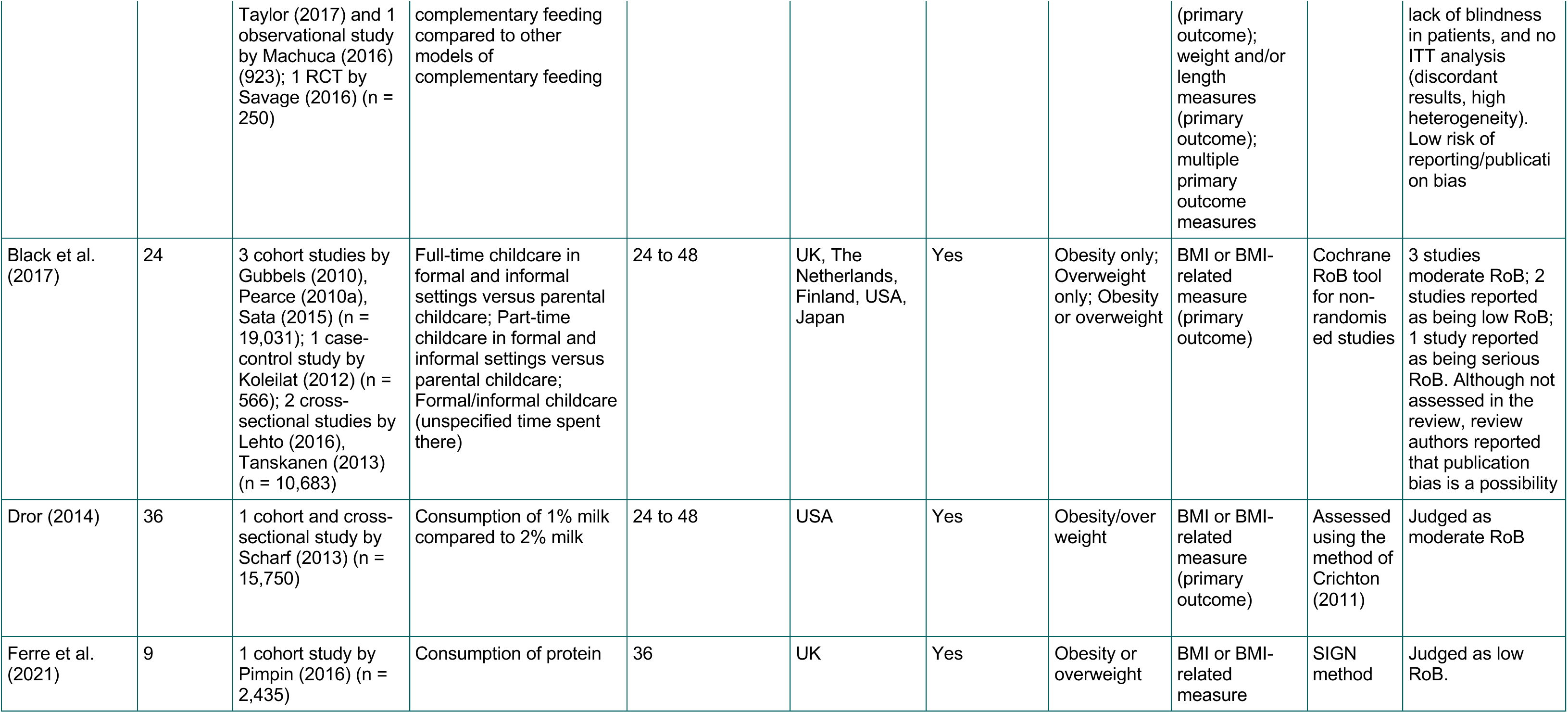
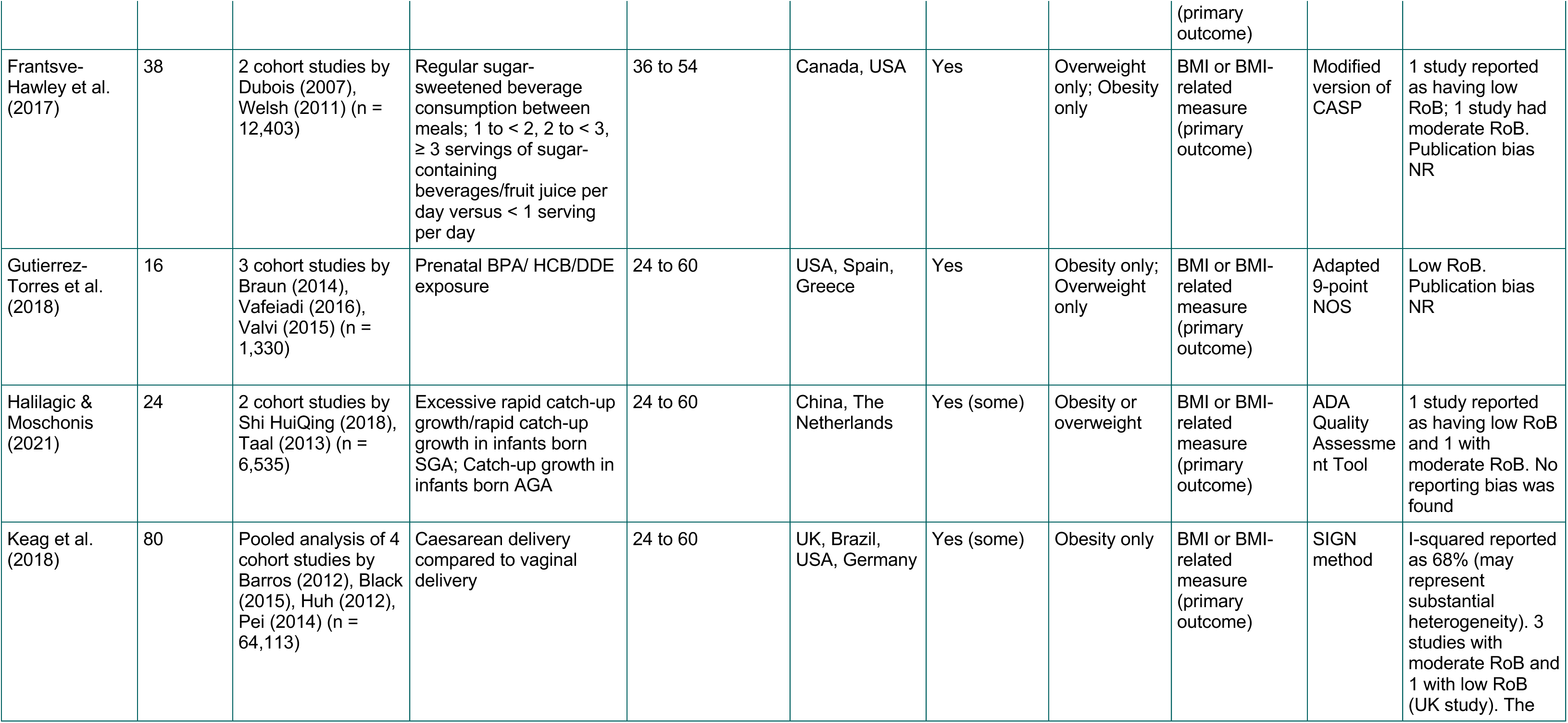
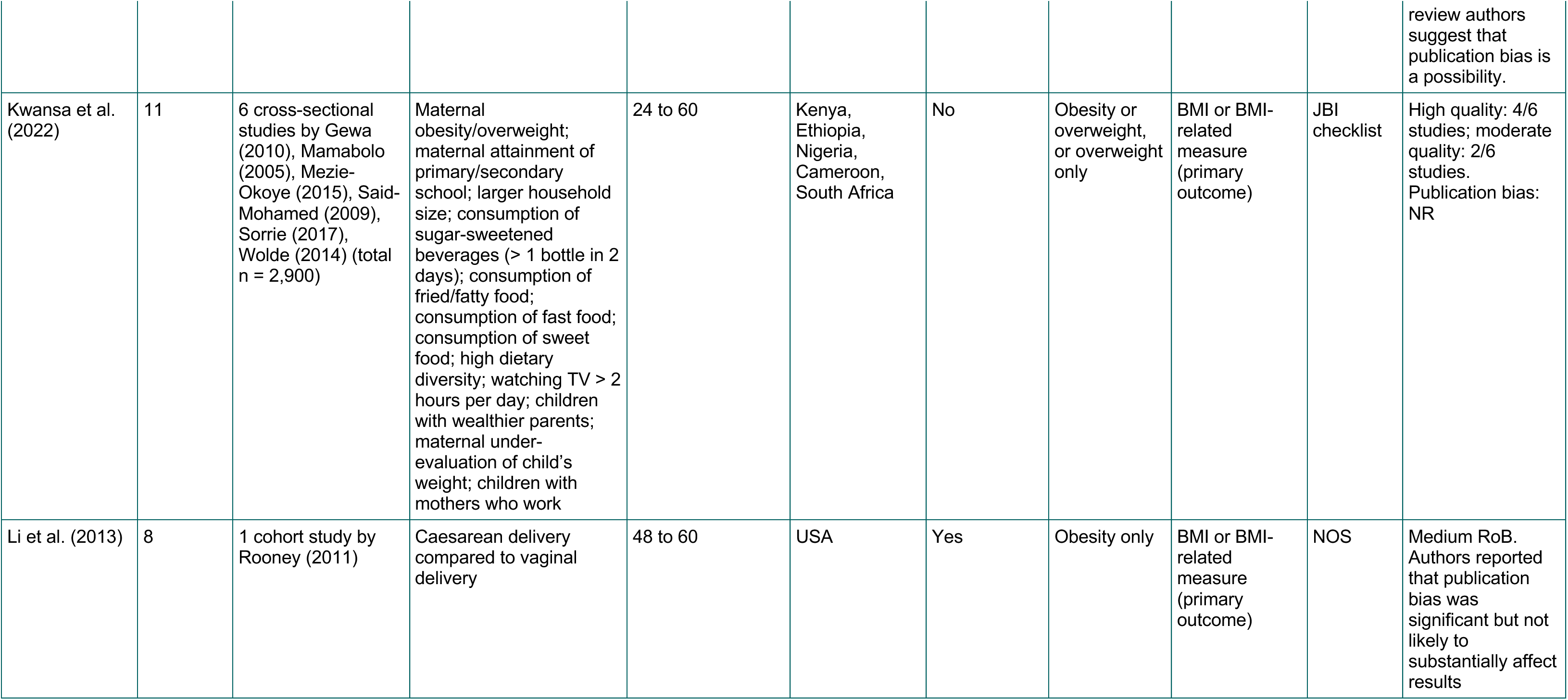
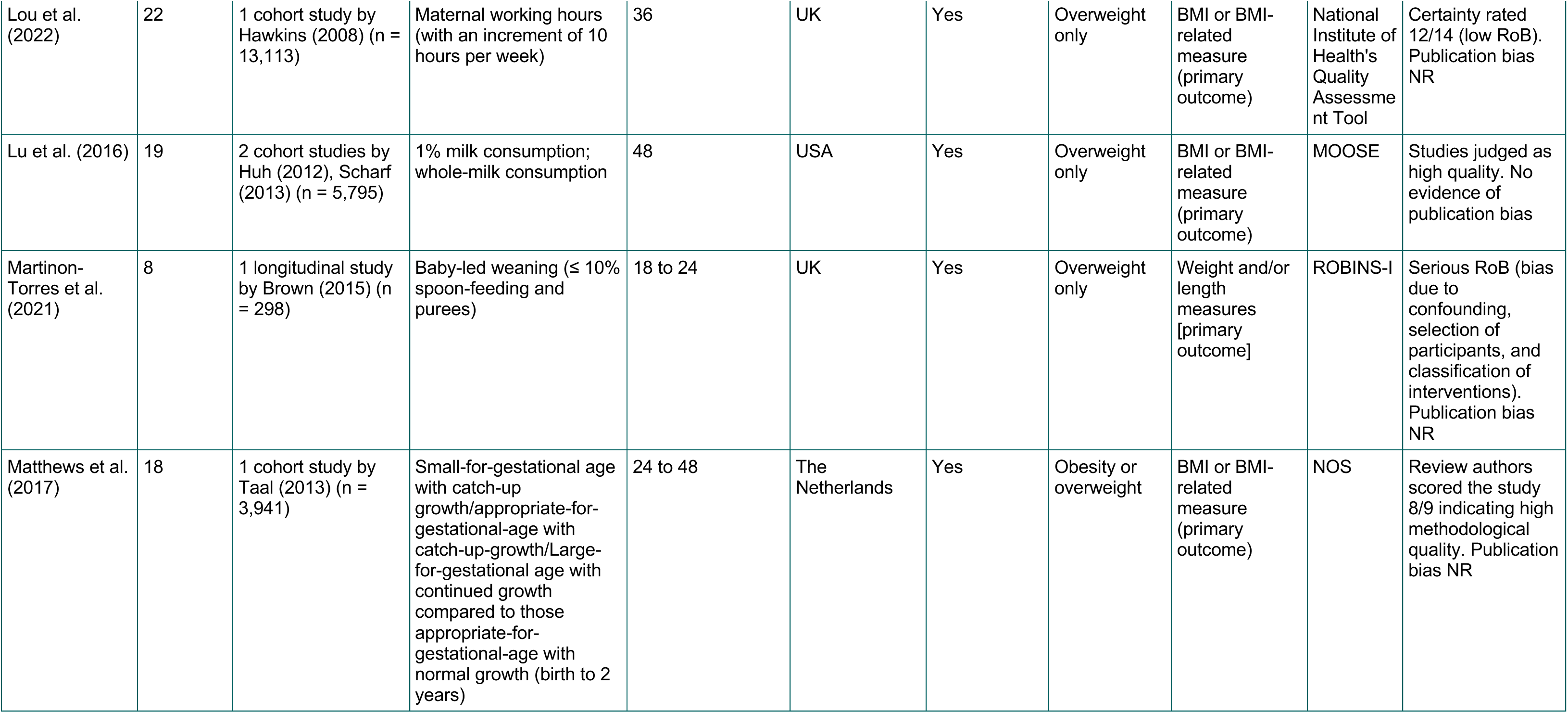
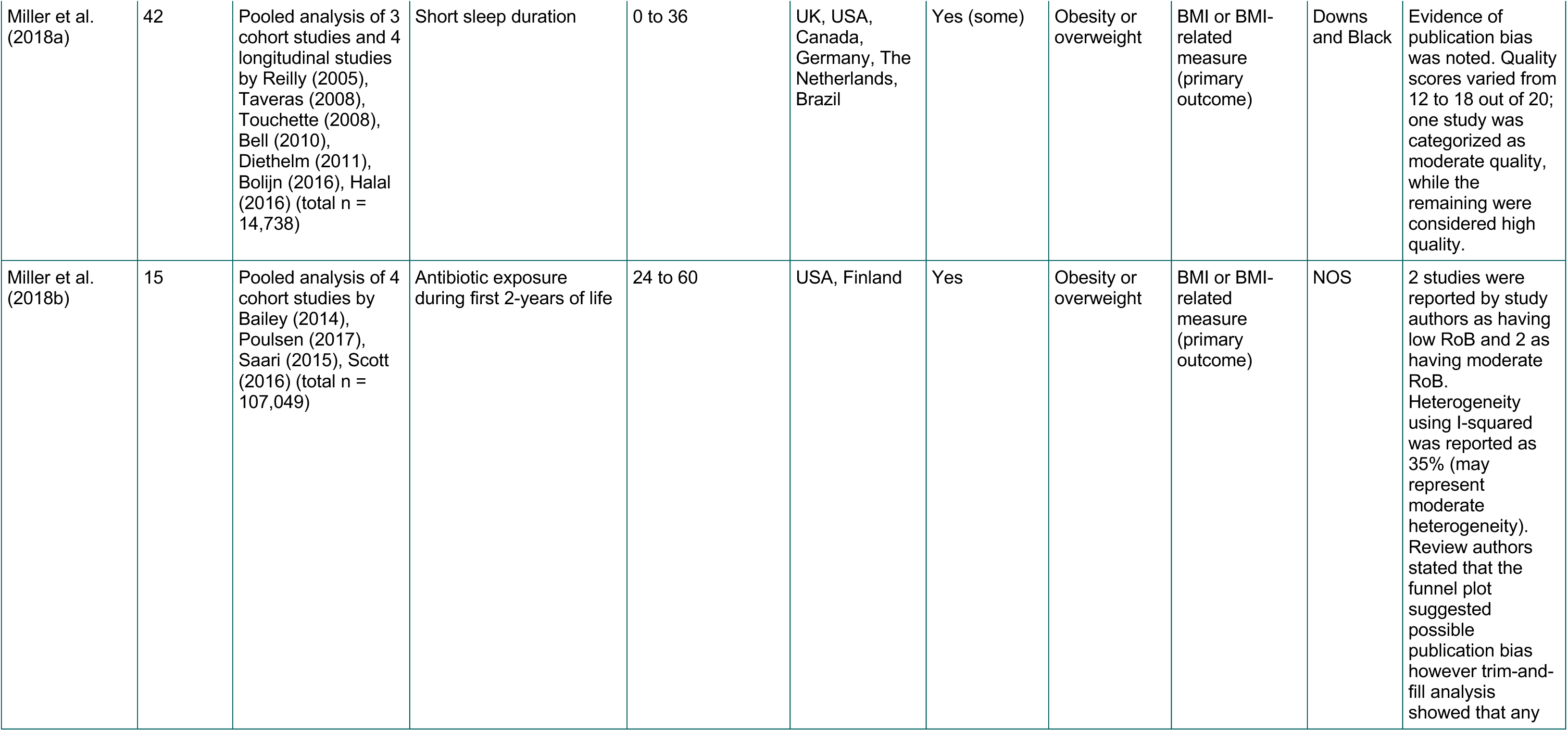
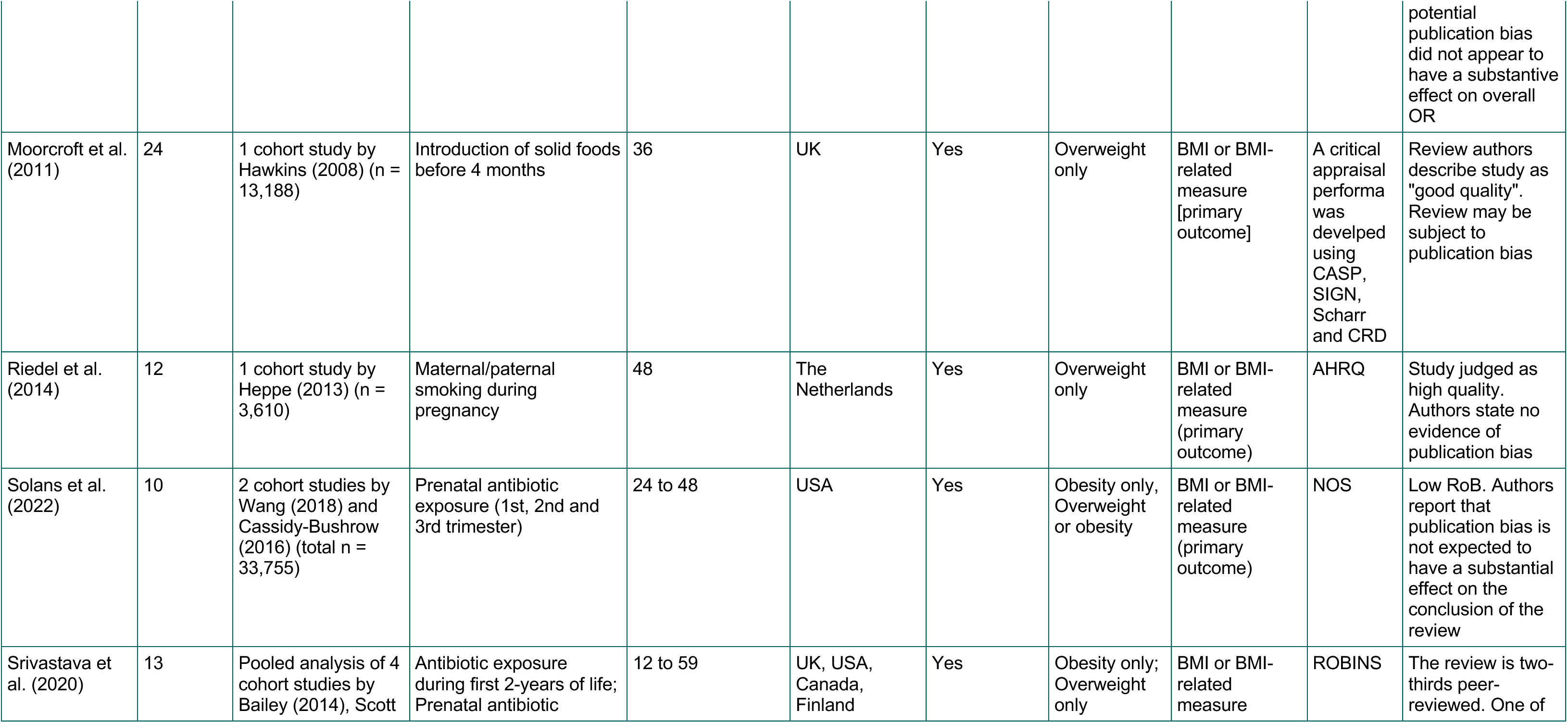
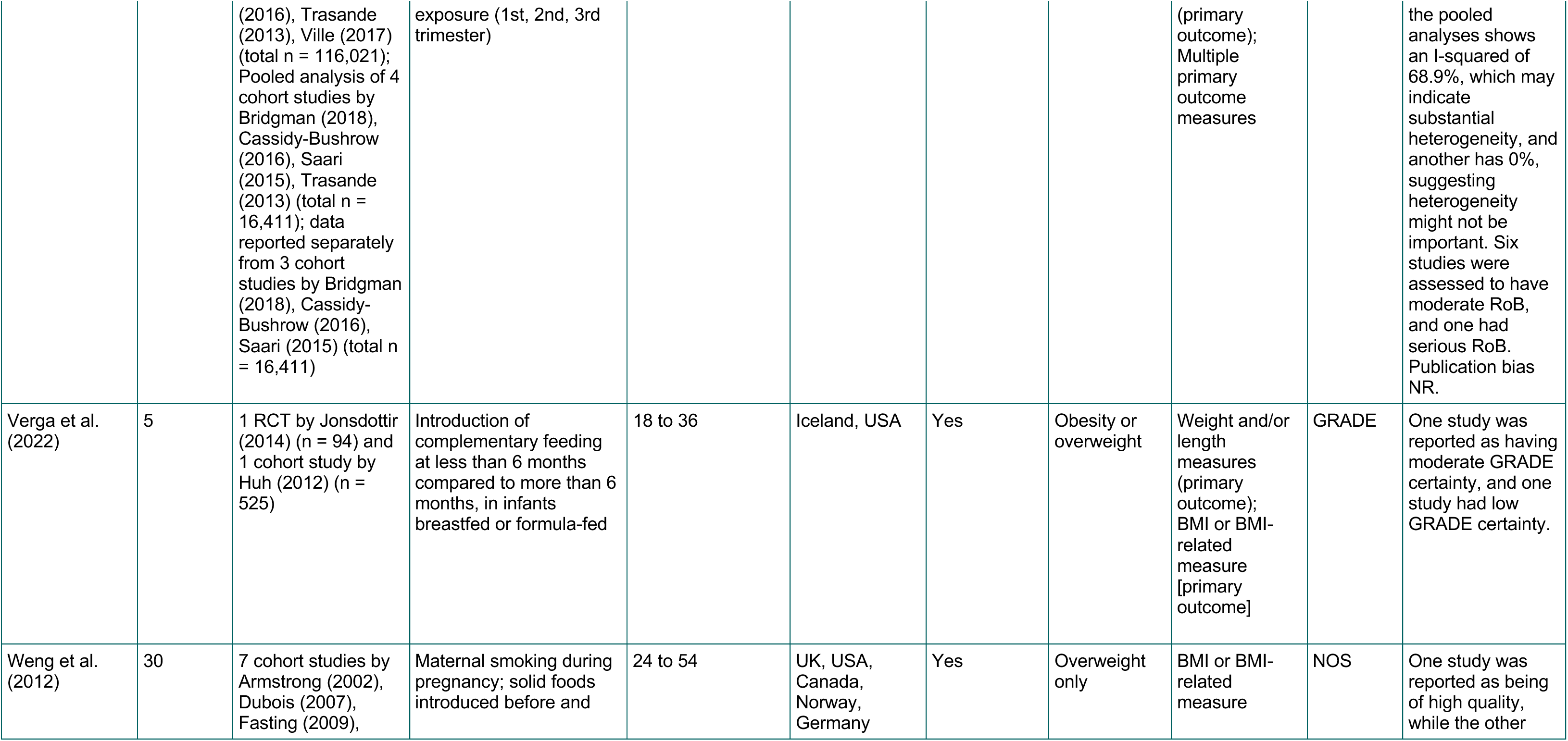
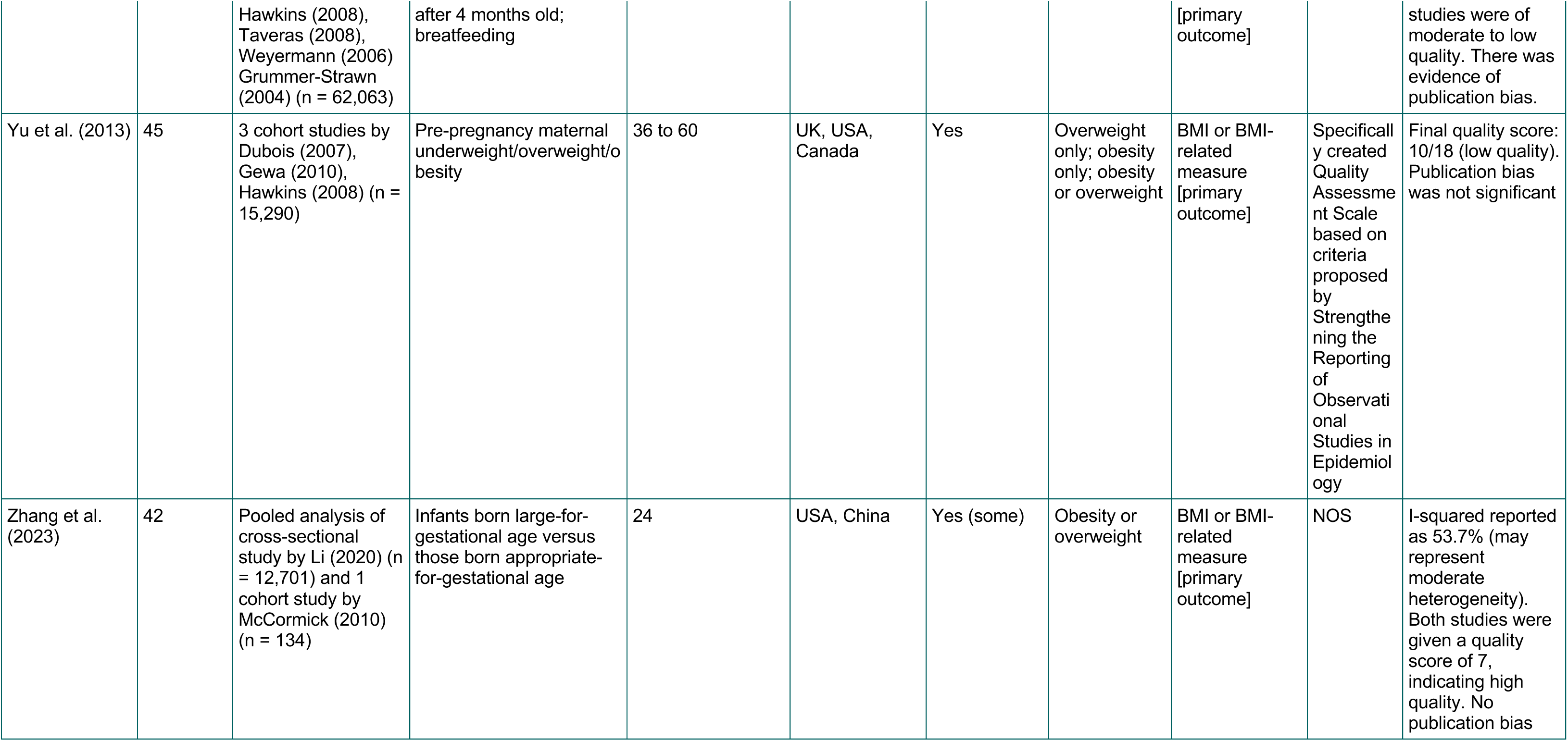
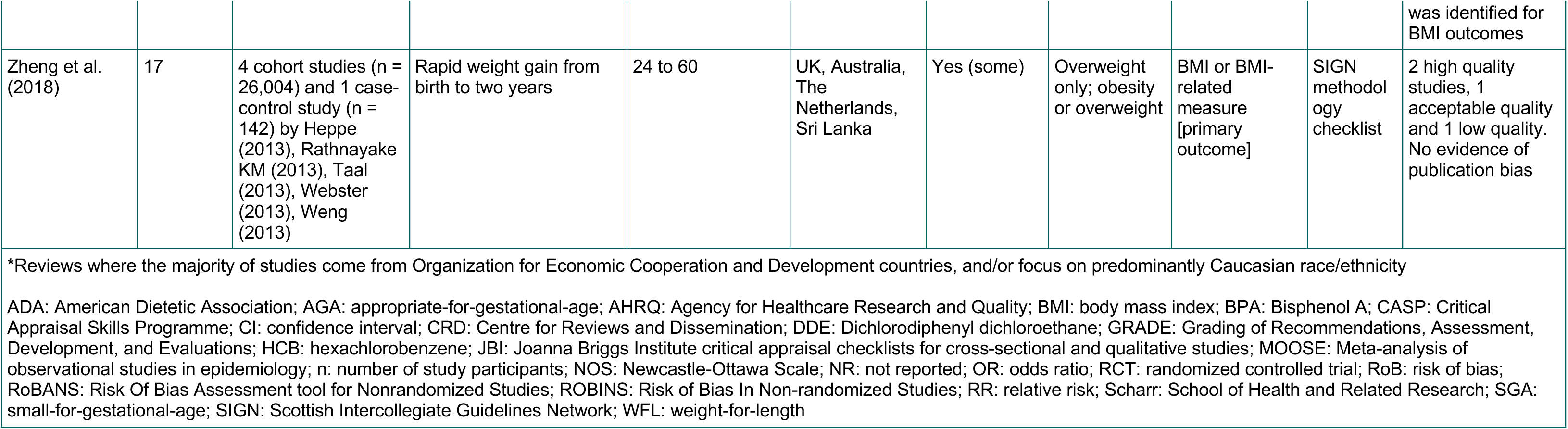
Summary of quantitative reviews included in this review.

**Table 20:** Summary of qualitative reviews included in this review.

| Review | Total number of studies in review | Types of studies | Risk Factor | Age of population at which outcomes measured (months) | Countries/ setting(s)/ population included in relevant studies* | Countries/ setting(s)/ population likely relevant to UK? | Themes | RoB tool used in review | Certainty assessed by review authors |
| --- | --- | --- | --- | --- | --- | --- | --- | --- | --- |
| Cartagena et al. (2014) | 35 | 9 qualitative, 15 cross-sectional, 9 cohort and 2 longitudinal studies. | Infant feeding practices: Factors Contributing to Infant Overfeeding with Hispanic Mothers. | 0 to 24 | Hispanic-only or multiethnic Mothers<br>USA | Unlikely | Breastfeeding and Formula Feeding Practices, Family and Cultural Influences on Maternal Feeding Beliefs and Practices, | N/A | N/A |
| Fraser et al. (2011) | 10 | 8 cross-sectional, 1 prospective longitudinal, 1 qualitative study. | Caregiver behaviour: Paternal influences on children's weight gain. | 6 to 67 | UK, USA, France, Australia. | Yes | Parenting styles, paternal perceptions of infant feeding practices and weight gain, and parental knowledge. | N/A | N/A |
| *Reviews where the majority of studies come from Organization for Economic Cooperation and Development countries, and/or focus on predominantly Caucasian race/ethnicity<br>N/A: not applicable; RoB: risk of bias |  |  |  |  |  |  |  |  |  |

### 6.3 Information available on request

The protocol, search strategies, and excluded studies for this rapid review are available on request.

## 8. APPENDIX

### APPENDIX 1: Medline search strategy

#### Ovid MEDLINE(R) ALL <1946 to December 07, 2023>

| # | Query | Results from 15 Dec 2023 |
| --- | --- | --- |
| 1 | exp Overweight/ | 277,244 |
| 2 | exp Weight Gain/ | 37,163 |
| 3 | exp Adiposity/ | 15,687 |
| 4 | exp Body Mass Index/ | 150,667 |
| 5 | obes*.tw. | 383,422 |
| 6 | (weight gain or overweight or over-weight or overeat* or over eat* or adipos* or macrosomia).tw. | 288,316 |
| 7 | ((bmi or body mass index) adj5 (gain* or increase* or change*)).tw. | 31,340 |
| 8 | (growth adj2 (measure* or development)).tw. | 60,421 |
| 9 | 1 or 2 or 3 or 4 or 5 or 6 or 7 or 8 | 739,043 |
| 10 | exp Child, Preschool/ | 994,333 |
| 11 | *Prenatal Exposure Delayed Effects/ | 25,574 |
| 12 | (baby or babies or neonat* or neo-nat* or f?etus or f?etal or newborn or infant* or infancy or toddler*).tw. | 1,098,507 |
| 13 | (child* adj2 (pre-school or preschool or nursery or young*)).tw. | 123,617 |
| 14 | (nursery adj2 school*).tw. | 1,275 |
| 15 | (early adj (years or childhood)).tw. | 38,577 |
| 16 | "first year* of life".tw. | 14,987 |
| 17 | 10 or 11 or 12 or 13 or 14 or 15 or 16 | 2,062,086 |
| 18 | 9 and 17 | 72,658 |
| 19 | exp fetal macrosomia/ | 3,315 |
| 20 | exp fetal weight/ | 2,255 |
| 21 | 19 or 20 | 5,410 |
| 22 | 18 or 21 | 75,620 |
| 23 | exp Risk Factors/ | 971,270 |
| 24 | exp Prognosis/ | 1,944,812 |
| 25 | (cause* or causative or causal* or determinant* or predict* or antecedent or prognos*).tw. | 5,651,757 |
| m26 | (risk adj2 factor*).tw. | 782,393 |
| 27 | 23 or 24 or 25 or 26 | 7,669,784 |
| 28 | exp "Systematic Review"/ | 247,209 |
| 29 | exp Meta-Analysis/ | 191,513 |
| 30 | exp Systematic Reviews as Topic/ | 12,234 |
| 31 | exp Meta-Analysis as Topic/ | 28,774 |
| 32 | (systematic review* or meta-analysis or review*).pt. | 3,425,548 |
| 33 | (systematic adj3 (review* or overview*)).ti,ab,kf. | 329,477 |
| 34 | (meta-analy* or metaanaly* or meta-synthes#s or metasynthes#s).tw. | 290,991 |
| 35 | (rapid review* or umbrella review*).ti,ab,kf. | 3,863 |
| 36 | 28 or 29 or 30 or 31 or 32 or 33 or 34 or 35 | 3,527,828 |
| 37 | 22 and 27 and 36 | **4,945** |

### APPENDIX 2. Measures of obesity other than BMI (possible secondary outcomes)

- Waist circumference
- Neck circumference
- Rohrer’s Ponderal Index (Rohrer’s Index, Ponderal Index or Corpulence Index)
- Benn’s Index
- Waist-to-height ratio (WHtR)
- Percentage body fat
- Waist-to-hip ratio (WHR)
- Body adiposity index (BAI)
- Skinfold thickness (SFT)
- Fat mass index (FMI)
- Near-infrared interactance (NIR)
- Densitometry (hydrostatic weighing, underwater [hydrostatic] weighting)
- Densitometry (air displacement plethysmography [ADP])
- Multicomponent models
- Dual-energy X-ray absorptiometry (DEXA)
- Deuterium dilution method (D2O)

